# Immune organization defines adaptive immune competence and clinical outcome in breast cancer

**DOI:** 10.64898/2026.07.17.26358324

**Authors:** Esther Sanfeliu, Elia Seguí, Anabel Martínez-Romero, Victor Albarrán-Fernández, Tomás Pascual, Mercedes Marín, Olga Martínez-Sáez, Raquel Gómez-Bravo, Isabel García-Fructuoso, Adela Rodríguez-Hernández, Benjamín Walbaum, Patricia Galván, Laura Angelats, Carlota Rubio-Pérez, Cristina Saura, Mafalda Oliveira, Eva Ciruelos, Luís Manso, Sònia Pernas, Maria Vidal, Adrienne G. Waks, Sara M. Tolaney, Laia Paré, Joel S. Parker, Patricia Villagrasa, Juan Manuel Ferrero-Cafiero, Charles M. Perou, Elias Campo, Josep Tabernero, Fara Brasó-Maristany, Aleix Prat

## Abstract

Tumor-infiltrating lymphocytes (TILs) are widely used to assess antitumor immunity in breast cancer but may not reflect the functional competence of adaptive immune responses. We show that immune organization, reflected by tertiary lymphoid structures (TLS) and coordinated humoral and cellular immune programs, represents a distinct dimension of tumor immunity beyond lymphocyte abundance. By integrating histologic, transcriptomic, spatial, and immune receptor profiling analyses across multiple breast cancer cohorts, we show that immune organization is associated with greater immune repertoire diversity, evidence of therapy-induced clonal selection, and improved clinical outcomes, independent of immune infiltration. Transcriptomic measures of immune organization retained independent prognostic value across external cohorts, whereas measures of immune infiltration did not. Furthermore, treatment-induced increases in immune organization, but not immune infiltration, were associated with therapeutic response. These findings identify immune organization as a dynamic and clinically measurable state of adaptive antitumor immunity with implications for prognosis, treatment monitoring, and therapeutic development in breast cancer.

**One Sentence Summary:** Spatially organized immune responses, rather than lymphocyte abundance alone, define clinically relevant antitumor immunity in breast cancer.

## INTRODUCTION

Immune profiling plays a central role in predicting clinical outcome and therapeutic response in breast cancer(*1*). Current approaches primarily quantify lymphocyte abundance, most commonly through the assessment of stromal tumor-infiltrating lymphocytes (TILs) on hematoxylin and eosin (H&E)–stained sections(*2–4*). Although TILs provide robust prognostic and predictive information, particularly in triple-negative breast cancer(*5, 6*), lymphocyte abundance alone may not fully capture the functional competence of adaptive antitumor immunity. Effective immunity depends not only on immune cell abundance but also on their spatial organization into coordinated cellular and humoral immune networks.

Emerging evidence suggests that organized adaptive immune responses represent a distinct dimension of tumor immunity. Tertiary lymphoid structures (TLS) are ectopic lymphoid aggregates containing spatially coordinated B-cell and T-cell regions and, in mature forms, germinal centers that support affinity maturation and humoral immunity(*7, 8*). Across multiple solid tumors, TLS have been associated with favorable prognosis and improved response to immunotherapy(*9–17*), suggesting that organized immune states may represent a functionally competent form of adaptive antitumor immunity beyond lymphocyte abundance alone(*12, 18–20*). However, the biological and clinical significance of immune organization in breast cancer remains incompletely understood.

Independent observations further support a role for immune organization in adaptive immunity. B-cell– and immunoglobulin-related gene expression signatures have been associated with treatment response and favorable clinical outcomes in breast cancer(*21–31*), indicating that coordinated B-cell and plasma-cell activity reflects effective adaptive immune responses. Together, these findings raise the possibility that immune organization integrates humoral and cellular immunity in ways not captured by lymphocyte abundance alone.

Together, these observations suggest that immune infiltration and immune organization represent related but distinct dimensions of the tumor immune microenvironment. However, whether immune organization captures biologically meaningful features of adaptive immune competence beyond lymphocyte abundance, provides independent clinical information, and represents a dynamic property that can be remodeled by therapy remains unknown.

Here, we define immune organization as a distinct dimension of tumor immunity characterized histologically by TLS and molecularly by coordinated humoral and cellular immune programs. By integrating histopathology, transcriptomics, spatial profiling, and immune repertoire analyses across multiple breast cancer cohorts, we demonstrate that immune organization captures adaptive immune competence beyond immune cell abundance and represents a dynamic and therapeutically remodelable component of antitumor immunity.

## RESULTS

### Immune organization represents a histologically distinct dimension of tumor immunity

To evaluate immune organization in breast cancer, we developed a standardized H&E-based framework based on two features: stromal lymphoid aggregate abundance and the presence of germinal centers (GCs), a hallmark of coordinated humoral immune responses and adaptive immune activation (**Fig. 1A,B**, **Fig. S1**). Tumors were classified into five categories reflecting increasing degrees of immune organization, ranging from absence of TLS to abundant TLS with GCs (**Fig. 1B**).

**Fig. 1.**
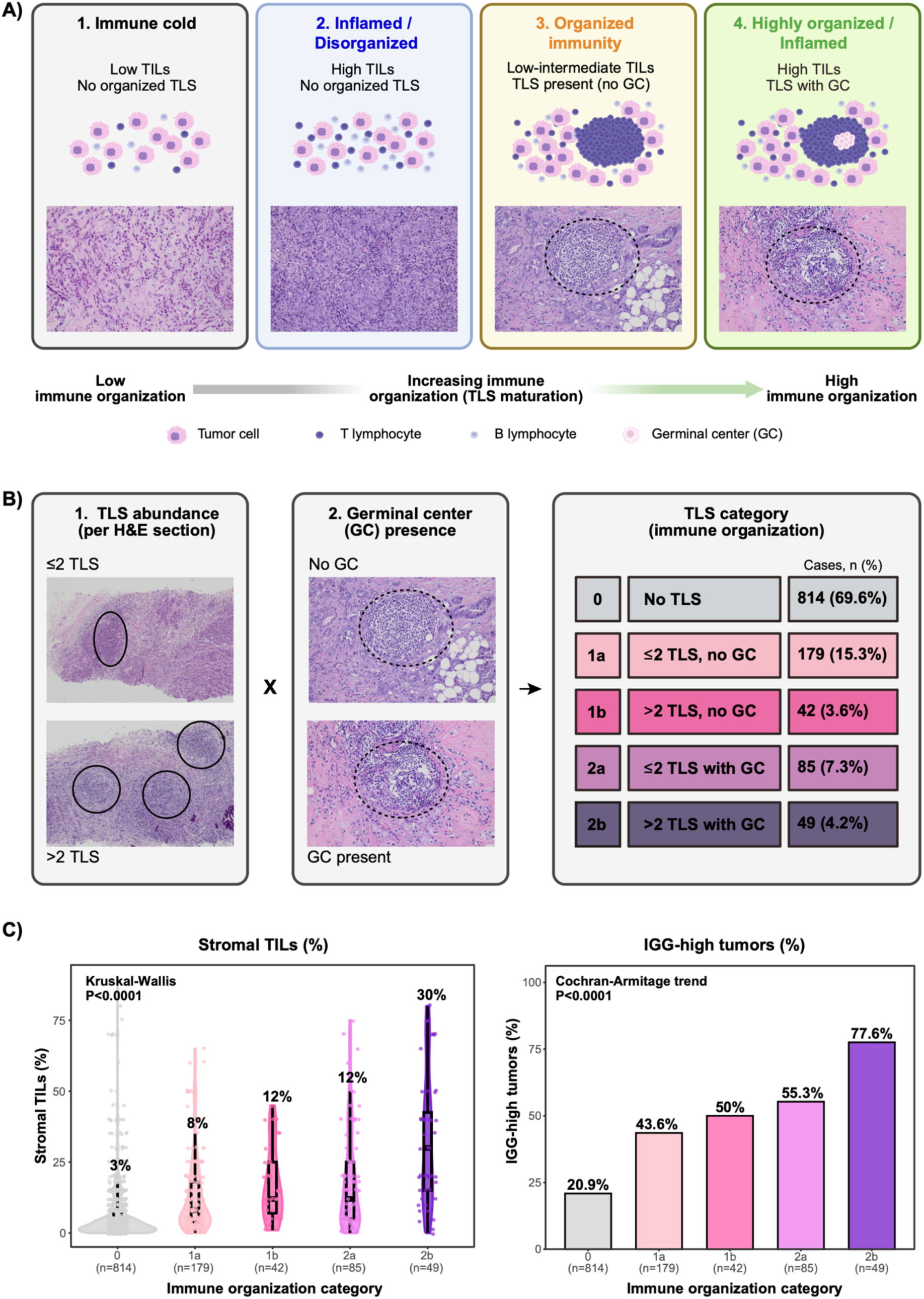
Immune organization is distinct from lymphocyte abundance. **(A)** Representative immune states illustrating distinct combinations of lymphocyte abundance and immune organization across the spectrum of breast tumor immune microenvironments. Tumors may exhibit low lymphocyte infiltration without TLS (“immune cold”), high TILs without evidence of immune organization (“inflamed/disorganized”), organized adaptive immune states with TLS despite only low-to-intermediate TIL levels, or highly organized and inflamed immune states characterized by mature germinal center (GC)-containing TLS. Representative H&E sections illustrate the corresponding histologic patterns. The horizontal gradient indicates increasing adaptive immune organization and TLS maturation from left to right. **(B)** H&E-based framework for histologic assessment of immune organization. TLS abundance was evaluated at low magnification and classified as ≤2 or >2 lymphoid aggregates per section. Germinal center (GC) presence was confirmed at higher magnification (20–40×). Combining TLS abundance and GC status defined five categories reflecting increasing degrees of immune organization: TLS 0 (no TLS), TLS 1a (≤2 TLS without GC), TLS 1b (>2 TLS without GC), TLS 2a (≤2 TLS with GC), and TLS 2b (>2 TLS with GC). **(C)** Association of immune organization with lymphocyte abundance and humoral immune activation. Increasing TLS categories were associated with progressively higher stromal TIL levels (left) and increasing prevalence of IGG-high tumors (right). Violin plots show the distribution of stromal TILs across TLS categories. Bar plots indicate the proportion of IGG-high tumors in each category. P values were calculated using Kruskal-Wallis (TILs) and Cochran-Armitage trend tests (IGG-high tumors). **Abbreviations:** TLS, tertiary lymphoid structures; GC, germinal center; TILs, tumor-infiltrating lymphocytes; H&E, hematoxylin and eosin; IGG, immunoglobulin G gene expression signature.

This framework identified four biologically distinct immune states defined by different combinations of lymphocyte abundance and immune organization (**Fig. 1A**). Notably, highly infiltrated tumors frequently lacked organized immune structures, whereas organized immune niches were observed despite only low-to-intermediate TIL levels, indicating that lymphocyte abundance alone does not fully capture the structural organization of adaptive immune responses. Interobserver reproducibility was high, with 92% concordance for TLS classification and strong correlation for TIL quantification (Spearman ρ = 0.95; **Fig. S2**).

TLS were identified across breast cancer subtypes and disease stages. Overall, 30.4% of tumors contained TLS, including 11.5% with mature GC-containing TLS (**Fig. 1B**, **Table S1**). Increasing immune organization was associated with progressively higher stromal TIL levels and increasing prevalence of tumors with high expression of an immunoglobulin G gene expression signature(*23, 27, 28*) (IGG-high tumors) (**Fig. 1C**, **Table S1**), supporting a coordinated relationship between lymphocyte infiltration, humoral immune activation, and adaptive immune organization.

### Immune organization is preserved across breast cancer disease contexts

Immune organization was observed across breast cancer disease stages and molecular subtypes, indicating that organized adaptive immune states are a conserved feature of breast cancer immunity. TLS were detected in 37.3% of primary tumors and 19.5% of metastatic lesions, with mature GC-containing TLS observed less frequently in metastatic samples (14.8% versus 6.4%; **Fig. S3**). Nevertheless, organized immune structures remained detectable across metastatic sites (**Fig. S4**), indicating that immune organization can persist beyond the primary tumor.

Similarly, immune organization was also evident across molecular subtypes, including HR+/HER2-, HER2-positive, and triple-negative breast cancers (**Table S1**, **Fig. S3**). HER2-positive tumors exhibited the highest overall TLS prevalence (36.0%), whereas triple-negative tumors showed the greatest enrichment of mature GC-containing TLS (16.0%). Together, these findings indicate that immune organization is broadly conserved across breast cancer disease contexts, supporting its role as a fundamental feature of adaptive antitumor immunity rather than a subtype- or stage-specific phenomenon.

### Immune infiltration and immune organization are related but non-equivalent

Among tumors with evaluable TLS, stromal TIL levels increased progressively across TLS categories (**Fig. 2A**), indicating an overall association between lymphocyte abundance and immune organization. Consistently, higher categorical TIL levels were enriched in TLS-positive tumors (**Fig. 2B**), and continuous TIL levels discriminated TLS presence and maturation (**Fig. 2C**).

**Fig. 2.**
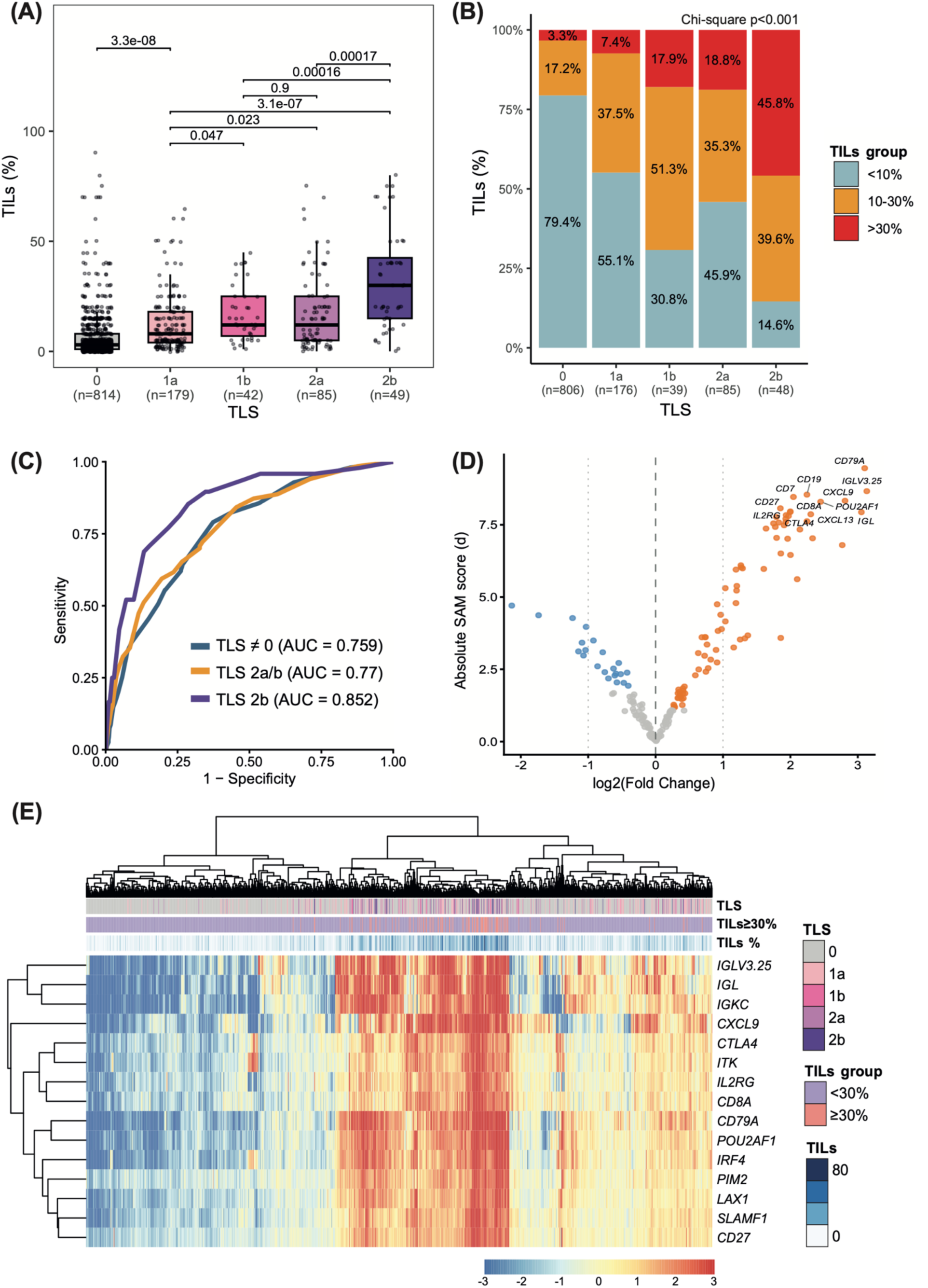
Lymphocyte abundance incompletely captures immune organization and primarily reflects broad immune activation. **(A)** Distribution of stromal TILs across TLS categories, showing progressively increasing lymphocyte abundance with increasing degrees of immune organization. Pairwise comparisons were performed using unpaired two-sided t-tests; unadjusted P values are shown. **(B)** Distribution of categorical TIL groups (<10%, 10–30%, >30%) across TLS categories (chi-square test), demonstrating enrichment of higher TIL levels in TLS-positive tumors. **(C)** Receiver operating characteristic (ROC) curves assessing the ability of continuous TIL levels to discriminate TLS presence and maturation, with corresponding area under the curve (AUC) values. **(D)** Volcano plot of differentially expressed genes comparing TIL-high tumors (≥30%) with TIL-low/intermediate tumors (<30%), highlighting broad activation of immune-related programs, including B-cell, T-cell, cytotoxic, and interferon-associated genes. **(E)** Heatmap of selected top differentially expressed genes associated with TIL-high tumors, illustrating the broad inflammatory transcriptional programs associated with high lymphocyte abundance across samples. **Abbreviations**: TLS, tertiary lymphoid structures; TILs, tumor-infiltrating lymphocytes; ROC, receiver operating characteristic; AUC, area under the curve.

Despite this overall association, lymphocyte abundance incompletely captured immune organization. High-TIL tumors frequently lacked TLS, whereas organized immune structures were also detected in tumors with only low-to-intermediate lymphocyte infiltration (**Fig. 2A,B**). Moreover, tumors with and without GC-containing TLS showed substantial overlap in TIL levels, indicating that lymphocyte abundance more closely reflects the abundance of lymphoid aggregates than the structural maturation of adaptive immune responses. Together, these findings demonstrate that immune infiltration and immune organization represent related but distinct dimensions of the tumor immune microenvironment.

To further define the biology captured by lymphocyte abundance, we compared transcriptomic profiles of TIL-high (≥30%) and TIL-low/intermediate tumors (<30%). TIL-high tumors showed broad upregulation of immune-related transcriptional programs, including B-cell, T-cell, cytotoxic, and interferon-associated genes (**Fig. 2D,E** and **Table S2**), consistent with a globally inflamed tumor microenvironment. These findings indicate that high lymphocyte abundance is associated with activation of multiple immune programs, including humoral immunity. However, these transcriptional programs were observed across tumors with both organized and disorganized immune states and therefore did not specifically capture immune organization. Together, these results suggest that lymphocyte abundance primarily reflects the overall magnitude of the immune response, whereas immune organization represents a distinct dimension of adaptive immunity that cannot be inferred from immune cell abundance alone.

### Immune organization is characterized by coordinated adaptive immune programs

To define transcriptomic programs associated with immune organization, we next examined humoral immune signatures. The IGG gene expression signature(*23, 27, 28*), a measure of humoral immune activation, was strongly associated with TLS presence and maturation. IGG levels increased progressively across TLS categories (**Fig. 3A**), with marked enrichment of IGG-high tumors in GC-containing TLS categories (**Fig. 3B**), supporting a close relationship between humoral immune activation and histologic immune organization.

**Fig. 3.**
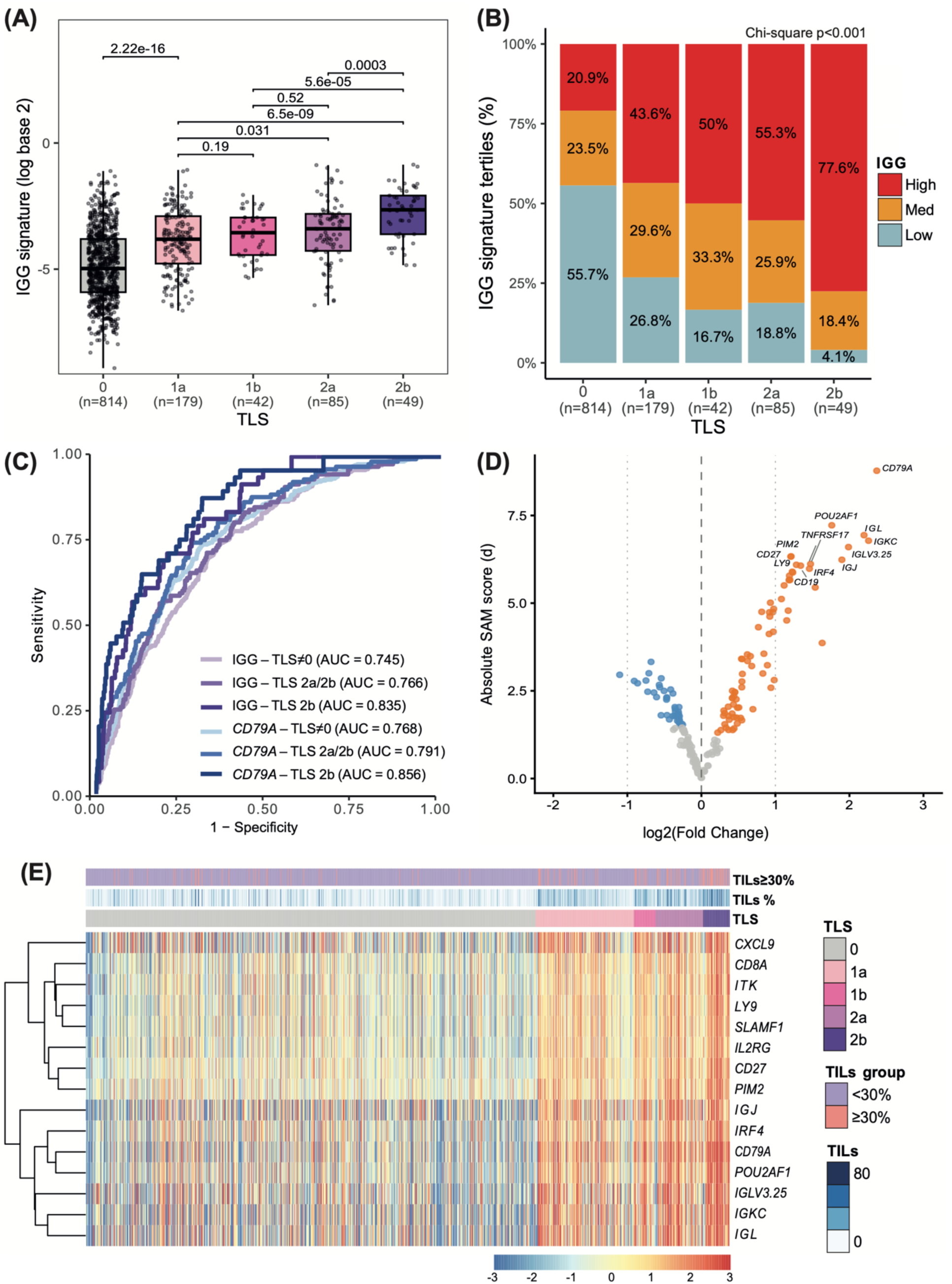
Humoral immune programs define transcriptomic states of immune organization. **(A)** Distribution of IGG signature scores across TLS categories, showing progressive increases with TLS presence and maturation. Pairwise comparisons were performed using unpaired two-sided t-tests; unadjusted P values are shown. No adjustment for multiple comparisons was applied. **(B)** Distribution of IGG signature tertiles (low, medium, high) across TLS categories. **(C)** Receiver operating characteristic (ROC) curves evaluating the ability of the IGG signature and CD79A expression to predict TLS presence and maturation; AUC values are shown. **(D)** Volcano plot of differentially expressed genes comparing tumors with mature TLS (TLS 2a/2b) versus tumors without mature TLS (TLS 0/1a/1b), highlighting coordinated upregulation of B-cell, plasma-cell, germinal center, T-cell, and antigen-presentation programs associated with organized immune states. **(E)** Heatmap of selected top differentially expressed genes associated with TLS, illustrating coordinated activation of humoral and adaptive immune pathways. **Abbreviations:** TLS, tertiary lymphoid structures; IGG, immunoglobulin G gene expression signature; ROC, receiver operating characteristic; AUC, area under the curve.

Similarly, expression of *CD79A*, a marker of B-cell receptor signaling, closely tracked with increasing TLS abundance and maturation (**Fig. 3C**), further supporting humoral immune markers as robust transcriptomic surrogates of immune organization.

To further define the transcriptional architecture of immune organization, we compared tumors with mature GC-containing TLS (TLS 2a/2b) with tumors lacking mature TLS (TLS 0/1a/1b). Differential expression analyses identified coordinated upregulation of B-cell, plasma-cell, germinal center, antigen-presentation, cytotoxic, and T-cell programs, consistent with integrated adaptive immune responses associated with organized immune states (**Fig. 3D,E** and **Table S3**). Importantly, immune organization was not defined by humoral immunity alone but by the coordinated activation of both humoral and cellular immune pathways. Together, these findings establish humoral immune signatures as transcriptomic surrogates of immune organization and demonstrate that immune organization reflects coordinated adaptive immune programs beyond those captured by lymphocyte abundance alone.

### Humoral immune programs complement lymphocyte abundance in identifying immune organization

To determine whether humoral immune programs capture features of immune organization beyond lymphocyte abundance, we evaluated the independent relationship of TILs and the IGG signature with mature GC-containing TLS. Both stromal TIL levels and the IGG signature remained independently associated with mature TLS after adjustment for subtype and tumor site (**Table S4**), indicating that lymphocyte abundance and humoral immune activation contribute complementary and non-redundant information to the identification of organized adaptive immune states.

Notably, the association between the IGG signature and mature TLS was strongest in tumors with low lymphocyte infiltration (IGG × TIL interaction, P = 0.003), suggesting that transcriptomic measures of humoral immune activation are particularly informative when immune cell abundance is limited and histologic evidence of immune organization is less apparent.

Consistent with this model, combined analyses incorporating TILs, IGG, and TLS abundance demonstrated that each component contributed independently to the characterization of organized immune states (**Table S5**). Together, these findings demonstrate that immune infiltration and humoral immune programs capture complementary biological dimensions of the tumor immune microenvironment, with humoral immune signatures providing additional information on immune organization beyond lymphocyte abundance alone.

### Immune organization independently predicts clinical outcome

We next investigated the prognostic relevance of immune organization across independent breast cancer cohorts. To enable scalable transcriptomic assessment of these complementary immune dimensions, we derived gene expression profiles capturing immune infiltration (TIL-GEP) and immune organization (TLS-GEP) from histologically annotated tumors and evaluated their performance across independent cohorts (**Fig. 4A** and **Table S6**).

**Fig. 4.**
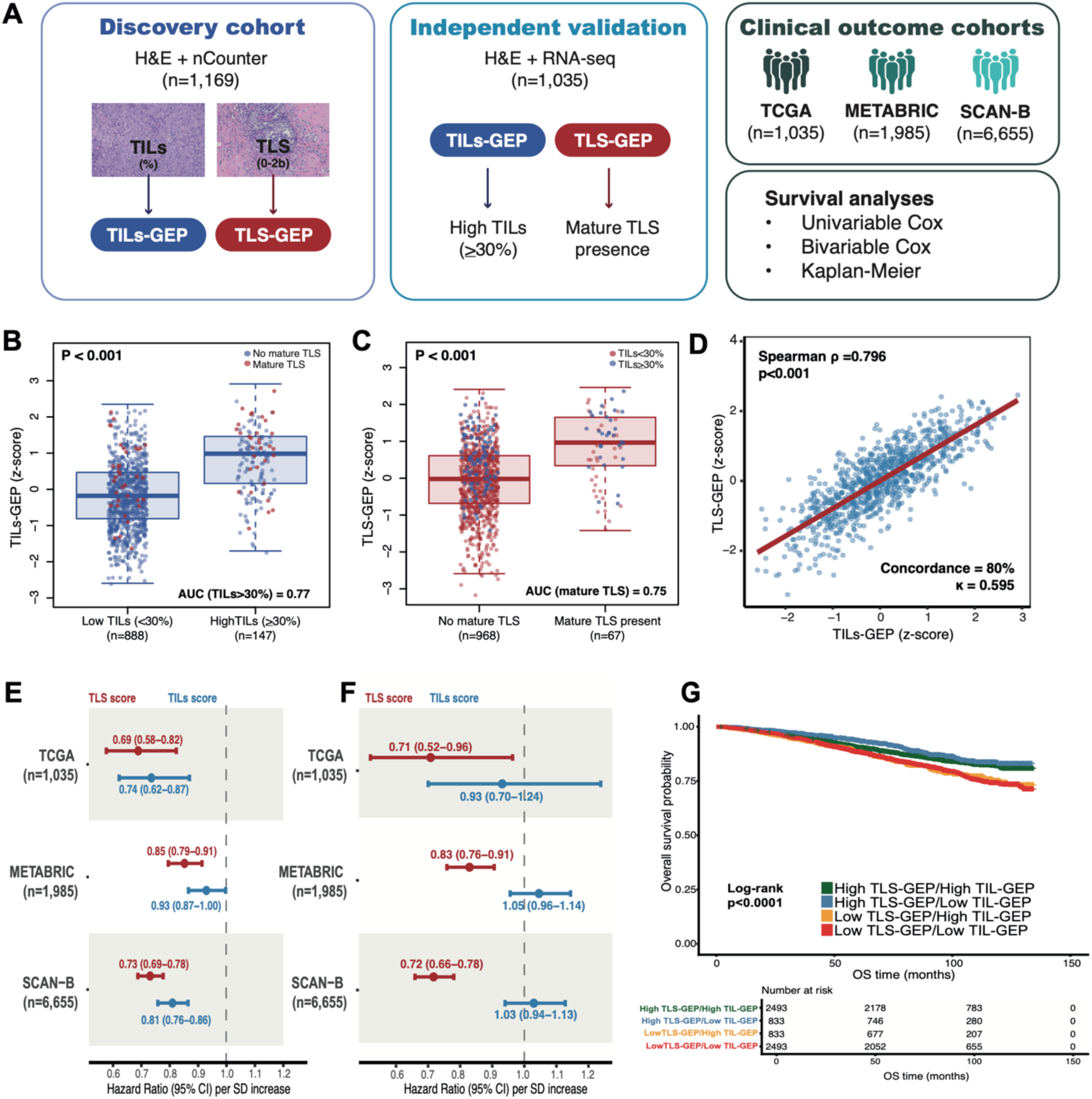
TLS-GEP captures immune organization and provides independent prognostic information beyond TIL-GEP. **(A)** Study design. TIL-GEP and TLS-GEP were derived from a discovery cohort of H&E-annotated tumors with gene expression data (n=1,169) and validated in an independent histologically annotated cohort (TCGA, n=1,035). Survival analyses were performed across independent cohorts (TCGA, METABRIC, and SCAN-B) using subtype-adjusted Cox models. **(B)** TIL-GEP scores according to TIL group (<30% versus ≥30%) in TCGA (n=1,035). Comparisons were performed using unpaired two-sided Wilcoxon rank-sum tests. AUC values from ROC analyses are shown. **(C)** TLS-GEP scores according to mature TLS status (absence versus presence) in TCGA (n=1,035). Comparisons were performed using unpaired two-sided Wilcoxon rank-sum tests. AUC values from ROC analyses are shown. **(D)** Correlation between TIL-GEP and TLS-GEP scores in TCGA (n=1,035), assessed using Spearman correlation (ρ and P values shown). Concordance and Cohen’s κ based on median dichotomization are also indicated, highlighting overlapping but non-equivalent dimensions of the tumor immune microenvironment. **(E)** Subtype-adjusted Cox models showing associations of TIL-GEP and TLS-GEP with overall survival across cohorts; hazard ratios (HRs) and 95% confidence intervals (CIs) per standard deviation (SD) increase are shown. **(F)** Multivariable Cox models including both signatures and adjusted for tumor subtype, showing independent prognostic effects of TLS-GEP after accounting for TIL-GEP; HRs and 95% CIs per SD increase are shown. **(G)** Kaplan-Meier curves in SCAN-B (n=6,655), stratified by combined TIL-GEP and TLS-GEP groups using median-based dichotomization. Tumors with high TLS-GEP showed favorable outcomes irrespective of TIL-GEP levels, whereas high TIL-GEP in the absence of elevated TLS-GEP was not associated with similarly improved survival. Statistical significance was assessed using the log-rank test. **Abbreviations**: TLS, tertiary lymphoid structures; TILs, tumor-infiltrating lymphocytes; GEP, gene expression profile; H&E, hematoxylin and eosin; ROC, receiver operating characteristic; AUC, area under the curve; HR, hazard ratio; CI, confidence interval; SD, standard deviation.

In an independent validation cohort (TCGA), both signatures aligned with their corresponding histologic features (**Fig. 4B,C** and **Fig. S5**). Although TIL-GEP and TLS-GEP were strongly correlated (Spearman ρ = 0.80, P < 0.001; **Fig. 4D** and **Fig. S6**), this correlation was incomplete, indicating that immune infiltration and immune organization capture overlapping but distinct features of tumor immunity. Consistent with the underlying biology, TLS-GEP preferentially reflected humoral and germinal center–associated immune programs, whereas TIL-GEP captured broader immune activation (**Table S6**).

We then evaluated the prognostic relevance of these immune dimensions across TCGA, METABRIC, and SCAN-B cohorts. Both TIL-GEP and TLS-GEP were associated with improved overall survival in subtype-adjusted models (**Fig. 4E**). However, when both signatures were included in the same model, TLS-GEP retained independent prognostic significance across all cohorts, whereas the association of TIL-GEP was attenuated and no longer significant (**Fig. 4F**). Kaplan-Meier analyses further demonstrated that tumors with high TLS-GEP showed favorable outcomes irrespective of TIL-GEP levels, whereas high TIL-GEP in the absence of elevated TLS-GEP was not associated with similarly improved survival (**Fig. 4G** and **Fig. S7**). These findings indicate that immune infiltration alone does not identify the tumors with the most favorable clinical outcomes.

Together, these findings indicate that immune organization provides prognostic information beyond immune infiltration. The consistent independent prognostic value of TLS-GEP across three independent cohorts supports immune organization as an independent and clinically relevant dimension of adaptive antitumor immunity.

### Immune organization complements intrinsic molecular risk

To determine whether immune organization captures clinically relevant information beyond intrinsic tumor biology, we evaluated the prognostic relevance of TLS-GEP and the IGG signature in HR+/HER2- tumors after adjustment for PAM50 Risk of Recurrence (ROR-P), a clinically validated molecular prognostic model. In multivariable Cox models, both TLS-GEP and the IGG signature remained independently associated with improved overall survival after adjustment for ROR-P across the TCGA, METABRIC, and SCAN-B cohorts (**Table S7**), indicating that immune organization captures prognostic information independent of intrinsic molecular subtype and proliferation-associated risk.

Consistent with these findings, tumors with high immune organization showed favorable outcomes across ROR categories, whereas tumors with low immune organization exhibited poorer outcomes despite similar intrinsic risk classification (**Fig. S8**). These results indicate that patients with comparable tumor-intrinsic biology can be further stratified according to the degree of immune organization.

Together, these findings indicate that immune organization represents a clinically relevant dimension of tumor biology that complements intrinsic molecular risk. The ability of TLS-GEP and IGG-based measures to provide prognostic information independently of ROR-P suggests that organized immune states capture aspects of host adaptive antitumor immunity not represented by current genomic classifiers. These findings support integrating immune organization with existing molecular prognostic frameworks to improve risk stratification in HR+/HER2- breast cancer.

### Immune organization defines adaptive immune competence

To determine whether immune organization reflects functional features of adaptive immune competence, we characterized T-cell receptor (TCR) repertoires in paired tumor samples collected before and after neoadjuvant therapy (**Fig. 5A-H** and **Table S8**).

**Fig. 5.**
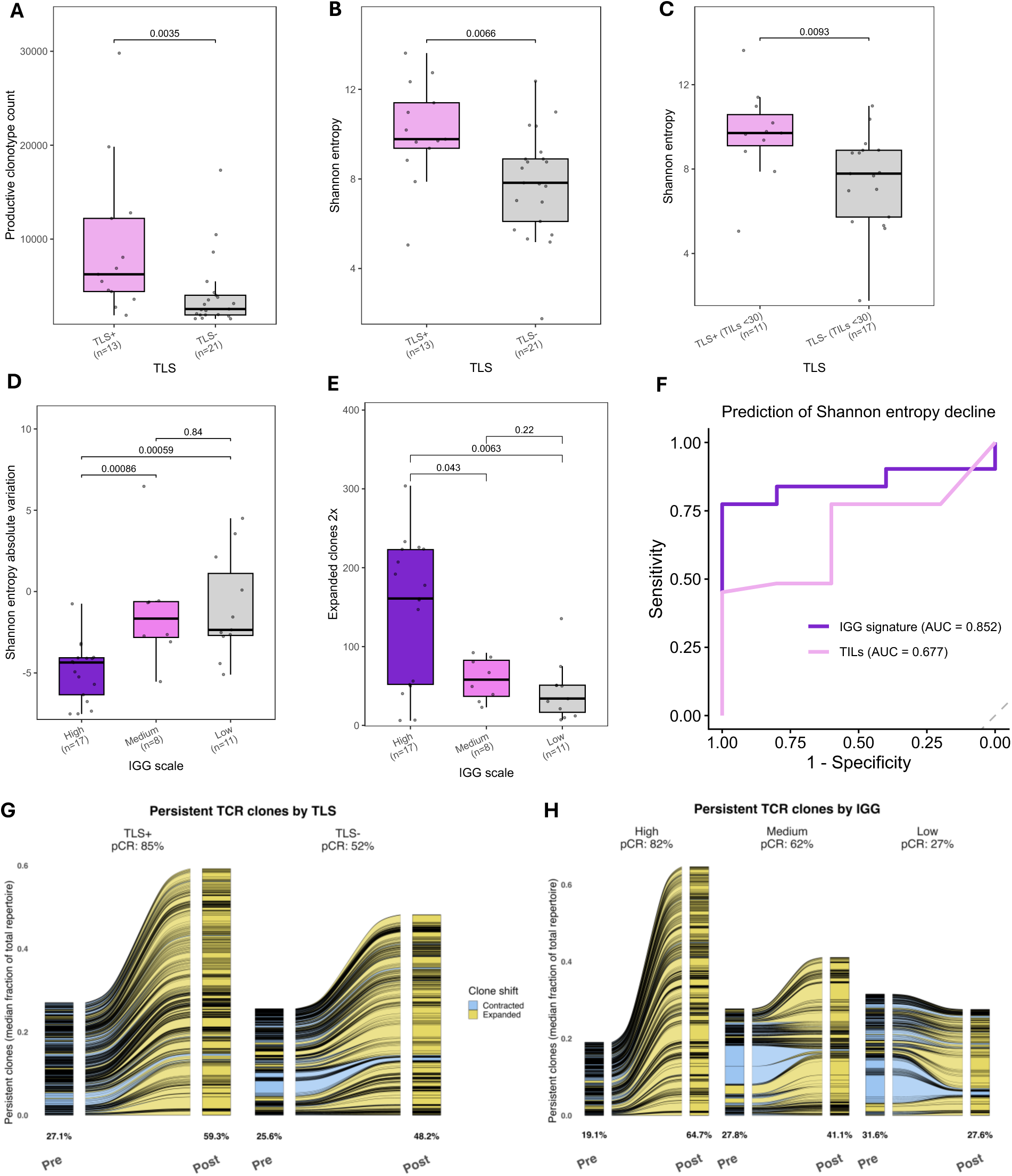
Immune organization defines adaptive immune competence through repertoire diversity and clonal selection. **(A–B)** Baseline T-cell receptor (TCR) repertoire characteristics according to TLS status, showing increased productive clonotype counts and higher clonal diversity (Shannon entropy) in TLS-positive tumors. **(C)** Baseline TCR diversity in tumors with low lymphocyte infiltration (TILs <30%), demonstrating higher repertoire diversity in TLS-positive tumors independent of lymphocyte abundance. **(D–E)** TCR clonal dynamics during therapy according to IGG signature expression, showing greater reductions in repertoire diversity and increased expansion of clonotypes in tumors with higher IGG levels, consistent with selective amplification of tumor-reactive T-cell populations. **(F)** Receiver operating characteristic (ROC) analysis demonstrating that IGG signature expression, but not baseline TIL levels (%), predicts entropy decline during treatment. Area under the curve (AUC) values are shown. **(G–H)** Clonotype-level analyses of persistent TCRs during therapy according to TLS status and IGG signature expression. Tumors with organized immune states showed preferential expansion of pre-existing tumor-resident clonotypes, whereas tumors lacking immune organization exhibited proportionally greater emergence of newly detected clonotypes. Similar patterns were observed in tumors achieving pathologic complete response (Fig. S10). **Abbreviations**: TLS, tertiary lymphoid structures; TILs, tumor-infiltrating lymphocytes; TCR, T-cell receptor; IGG, immunoglobulin G gene expression signature; ROC, receiver operating characteristic; AUC, area under the curve.

At baseline, tumors with organized immune states exhibited broader and more diverse adaptive immune repertoires. TLS-positive tumors showed increased numbers of productive clonotypes, and higher TCR diversity compared with TLS-negative tumors (**Fig. 5A,B**). Similar associations were observed with higher IGG signature expression, whereas tumors lacking immune organization displayed more oligoclonal repertoires, consistent with reduced adaptive repertoire breadth. Comparable findings were observed in B-cell receptor analyses (**Fig. S9**).

Importantly, these associations persisted in tumors with low lymphocyte infiltration (TILs <30%) (**Fig. 5C**), indicating that immune organization captures features of adaptive immune competence beyond immune cell abundance alone. In multivariable analyses, the association between TLS and baseline repertoire diversity remained significant after accounting for TIL levels (**Fig. S9**), demonstrating that immune organization, rather than immune cell abundance alone, is associated with repertoire diversity.

We next examined therapy-induced clonal dynamics. Tumors with high IGG expression exhibited greater reductions in repertoire diversity and increased expansion of clonotypes during treatment (**Fig. 5D,E**), consistent with selective expansion of tumor-reactive T-cell clones. Notably, IGG expression, but not baseline TIL levels, predicted entropy decline during therapy (**Fig. 5F**), linking immune organization to functional clonal remodeling rather than simply immune cell abundance.

At the clonotype level, organized immune states were associated with preferential expansion of persistent tumor-resident T-cell clones. TLS-positive and IGG-high tumors showed greater expansion of pre-existing clonotypes during therapy, whereas tumors lacking immune organization exhibited proportionally greater emergence of newly detected clonotypes (**Fig. 5G,H** and **Fig. S10**). Similar patterns were observed in tumors achieving pathologic complete response (**Fig. S10**). These findings suggest that organized immune states preferentially amplify pre-existing tumor-reactive T-cell populations rather than relying predominantly on de novo clonal recruitment.

Together, these findings provide functional evidence that immune organization reflects adaptive immune competence. Organized immune states are characterized by broader baseline repertoire diversity, therapy-induced clonal selection, and preferential expansion of persistent tumor-resident T-cell populations, defining a functional state of coordinated adaptive antitumor immunity.

### Spatial transcriptomics reveals a continuum of immune organization

To define the spatial architecture underlying immune organization, we performed spatial single-cell transcriptomic profiling using the 10x Genomics Xenium platform across breast tumors representing diverse immune contexts and TLS states (**Fig. 6A** and **Tables S9-S10**). Regions of interest (ROIs) included TLS-containing regions, non-TLS tumor microenvironment (TME) regions, and tumor-cell regions.

**Fig. 6.**
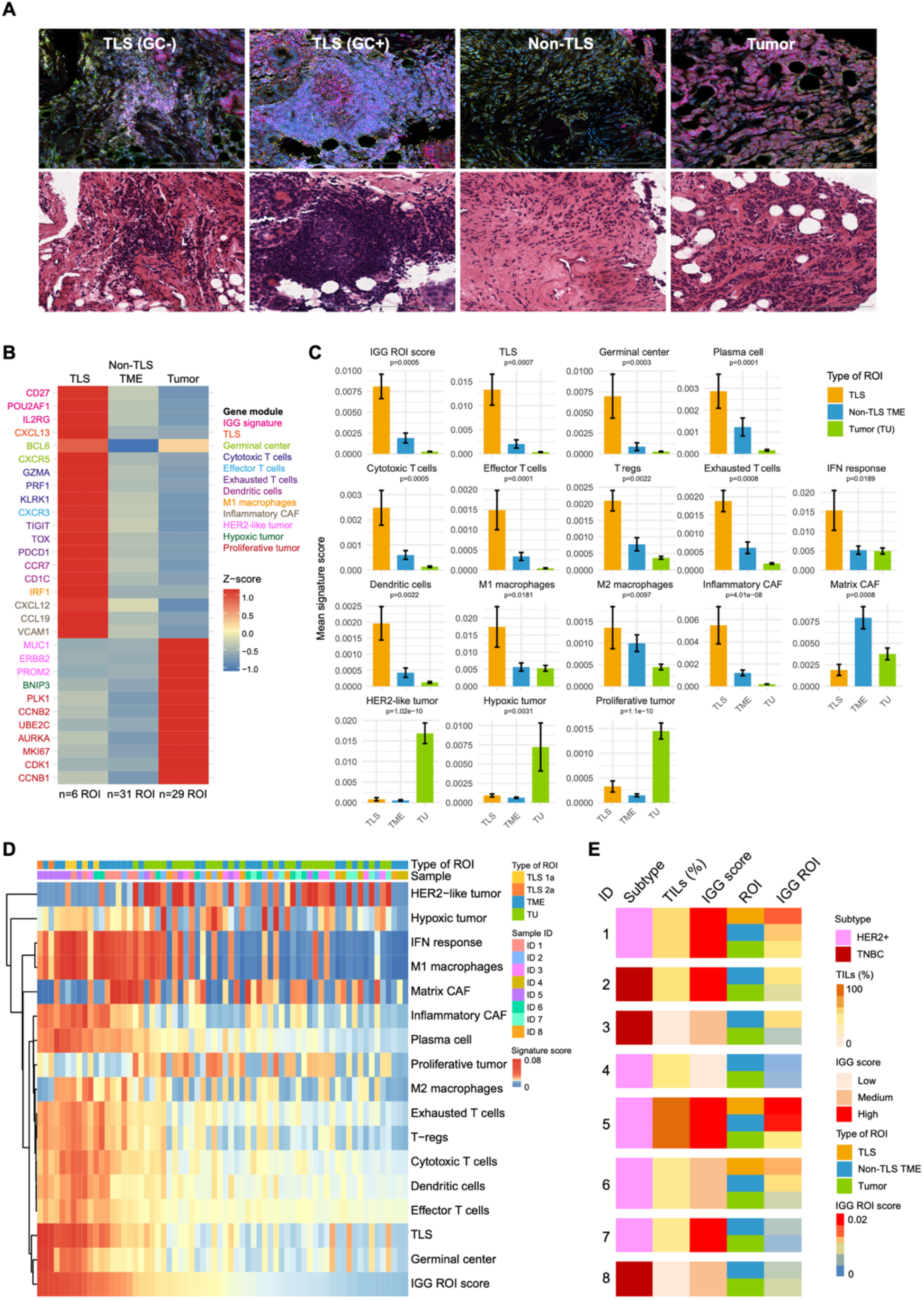
Spatial transcriptomics reveals coordinated immune niches and a continuum of immune organization. **(A)** Representative spatial images illustrating distinct immune architectures, including TLS without germinal centers (GCs), TLS with GCs, non-TLS tumor microenvironment (TME), and tumor-cell regions. TLS regions display densely organized immune aggregates, whereas non-TLS TME regions show more diffuse immune infiltration. **(B)** Heatmap of top differentially expressed immune-related genes across spatial compartments (TLS, non-TLS TME, and tumor-cell regions), illustrating compartment-specific enrichment. **(C)** Quantification of immune-related gene-expression signatures across spatial compartments, showing enrichment of plasma-cell, dendritic-cell, cytotoxic T-cell, germinal center, and IGG-related programs in TLS regions compared with non-TLS TME and tumor-cell regions. **(D)** Heatmap of all regions of interest (ROIs) based on gene-expression signatures, revealing coordinated immune programs and clustering of regions with high IGG-related expression and humoral immune activity. IGG-related expression was calculated as the median expression of IGG signature genes represented in the Xenium panel. **(E)** Summary of spatial heterogeneity across samples, highlighting that humoral and germinal center-associated programs are detectable in a subset of non-TLS TME regions despite the absence of morphologically identifiable TLS, consistent with a continuum of immune organization. **Abbreviations:** TLS, tertiary lymphoid structures; TME, tumor microenvironment; ROI, region of interest; GC, germinal center.

Spatial profiling demonstrated a progressive organization of adaptive immune programs across these compartments rather than discrete immune states. TLS regions displayed densely organized immune aggregates, including structures with and without germinal centers (GCs), whereas non-TLS TME regions exhibited more diffuse immune infiltration. Tumor-cell regions showed relative immune exclusion (**Fig. 6A**).

Transcriptomic analyses demonstrated graded enrichment of adaptive immune programs (**Fig. 6B,C**). TLS regions were enriched for humoral immunity, antigen presentation, plasma-cell, germinal center, and cytotoxic T-cell programs, whereas tumor-cell regions showed minimal immune activity. Non-TLS TME regions displayed intermediate immune programs, supporting a continuum of immune organization extending from immune-excluded tumor regions to highly organized adaptive immune niches.

Across ROIs, coordinated immune programs characteristic of organized adaptive immune responses clustered together spatially (**Fig. 6D**). Importantly, regions with high IGG-related expression exhibited coordinated activation of humoral, germinal center, and cytotoxic immune programs, indicating that immune organization reflects an integrated adaptive immune ecosystem rather than isolated B-cell responses.

Notably, a subset of non-TLS TME regions, particularly in tumors with high bulk IGG expression and TLS elsewhere in the tissue, exhibited enrichment of humoral and TLS-associated transcriptional programs despite lacking identifiable TLS or GCs (**Fig. 6E**). These observations indicate that histologically recognizable TLS represent the most organized end of a broader spatial continuum of adaptive immune organization rather than a discrete biological entity.

### Immune organization is dynamically remodeled by therapy

To determine whether immune organization represents a dynamic and therapeutically remodelable component of antitumor immunity, we analyzed paired tumor samples collected before and after treatment across multiple window-of-opportunity studies evaluating mechanistically distinct therapeutic approaches.

In three studies with centralized histopathologic assessment of TLS and TILs (PROMETEO-I, PROMETEO-II, and short preoperative endocrine therapy), treatment induced substantial remodeling of TLS architecture, characterized by transitions from TLS-negative tumors to tumors containing immature or mature TLS (**Fig. 7A**). In contrast, changes in stromal TIL categories were generally more modest (**Fig. S11**), and tumors exhibiting TLS induction frequently remained within the same TIL category over time (**Fig. 7E**). Representative cases illustrated that TLS induction could occur either together with increased lymphocyte infiltration or despite little or no change in stromal TIL levels (**Fig. 7B**). These findings indicate that therapeutic immune remodeling preferentially involves structural reorganization of the immune microenvironment rather than simply increasing immune cell abundance.

**Fig. 7.**
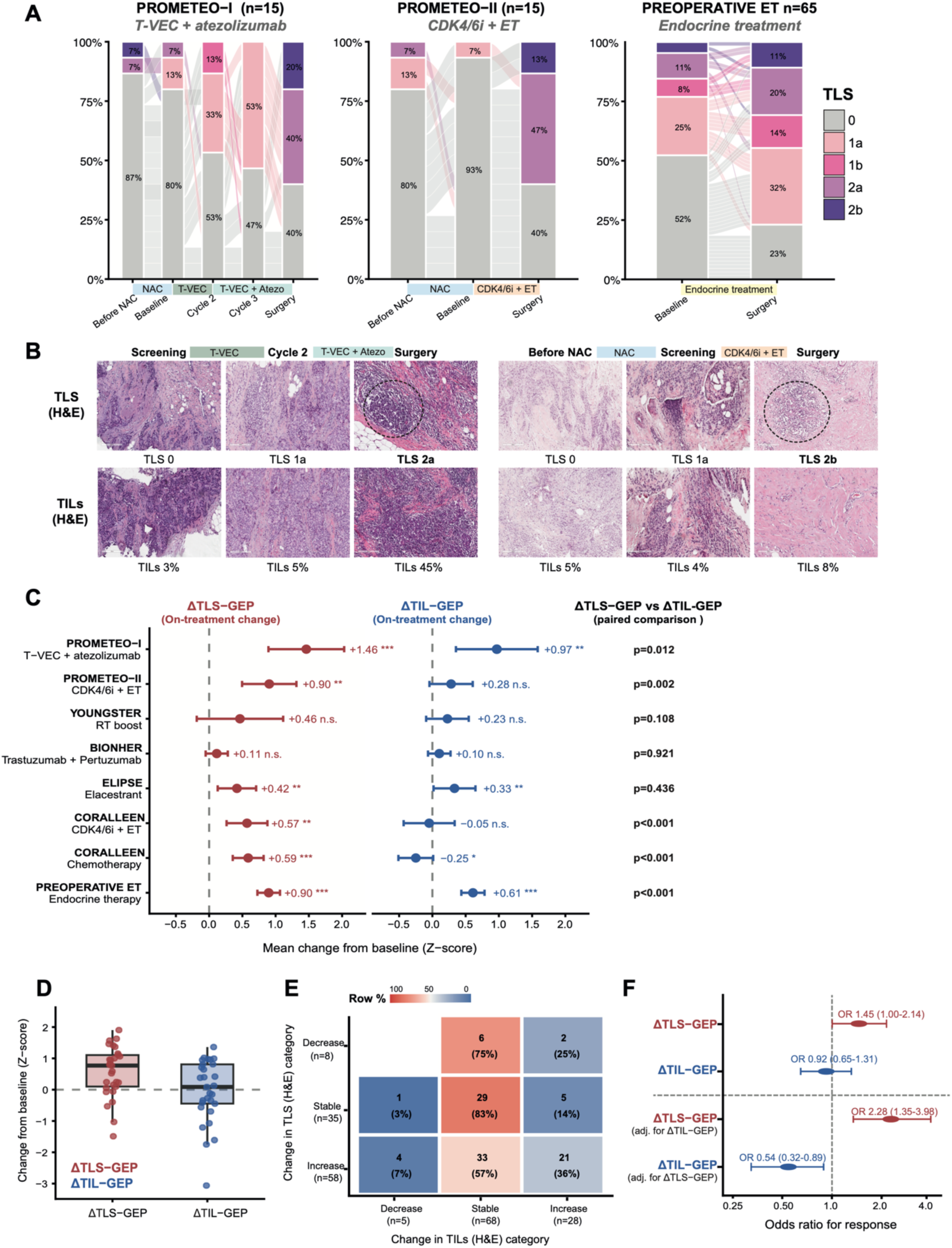
Therapeutic immune remodeling preferentially induces immune organization and is associated with treatment response. **(A)** Changes in histologic TLS categories across paired tumor samples from PROMETEO-I (T-VEC plus atezolizumab, n=15), PROMETEO-II (CDK4/6 inhibitor plus endocrine therapy, n=15), and short preoperative endocrine therapy (n=65). Alluvial plots show transitions between TLS categories (0, 1a, 1b, 2a, and 2b) from baseline to on-treatment and/or surgery samples. **(B)** Representative H&E images illustrating treatment-induced immune remodeling. Left panels show a case with coordinated increases in TLS category and stromal TILs following T-VEC plus atezolizumab. Right panels show a case with TLS induction following CDK4/6 inhibitor plus endocrine therapy despite minimal changes in stromal TIL levels. Dashed circles indicate TLS structures. **(C)** Mean on-treatment changes (Δ) in TLS-GEP and TIL-GEP across studies evaluating distinct therapeutic modalities. Points represent mean changes from baseline (Z-score units), and error bars indicate 95% confidence intervals. Asterisks indicate significance of on-treatment changes relative to baseline (*P<0.05, **P<0.01, ***P<0.001; n.s., not significant). P values shown correspond to paired comparisons between ΔTLS-GEP and ΔTIL-GEP within each study. **(E)** Representative distribution of treatment-induced changes in TLS-GEP and TIL-GEP in the CORALLEEN CDK4/6 inhibitor plus endocrine therapy cohort (n=29). Boxplots display median, interquartile range, and individual observations. **(F)** Relationship between changes in histologic TLS category and stromal TIL category in paired samples from PROMETEO-I, PROMETEO-II, and preoperative endocrine therapy cohorts. Values indicate the number and percentage of tumors within each category. **(G)** Logistic regression models evaluating associations between treatment-induced changes in TLS-GEP and TIL-GEP and treatment response. Odds ratios (ORs) and 95% confidence intervals (CIs) are shown for models adjusted for study and for models including both signatures simultaneously. Increases in TLS-GEP, but not TIL-GEP, were associated with improved response. **Abbreviations**: TLS, tertiary lymphoid structures; TILs, tumor-infiltrating lymphocytes; GEP, gene expression profile; H&E, hematoxylin and eosin; ET, endocrine therapy; CDK4/6i, cyclin-dependent kinase 4/6 inhibitor; OR, odds ratio; CI, confidence interval.

Consistent with these histologic observations, treatment-induced changes in organization-associated transcriptional programs were generally greater than changes in infiltration-associated programs across studies. In several therapeutic contexts, TLS-GEP increased despite limited changes in TIL-GEP, indicating that therapeutic remodeling preferentially enhanced immune organization rather than simply increasing immune infiltration (**Fig. 7C,D** and **Table S11**). Furthermore, increases in TLS-GEP were strongly correlated with increases in the IGG signature (**Fig. S12**), supporting coordinated activation of humoral immune programs during therapy-induced immune remodeling.

Finally, in models adjusted for study, treatment-induced increases in TLS-GEP were associated with improved treatment response, whereas changes in TIL-GEP were not (**Fig. 7F** and **Table S12**). This association persisted after accounting for concurrent changes in TIL-GEP. These findings indicate that dynamic remodeling of immune organization, rather than immune infiltration alone, is associated with therapeutic response across diverse treatment context.

Together, these findings establish immune organization as a dynamic and therapeutically remodelable feature of adaptive antitumor immunity. The preferential induction of immune organization and its association with treatment response suggest that coordinated adaptive immune remodeling, rather than increased immune cell abundance alone, is a hallmark of effective antitumor immunity.

Collectively, these observations support a model in which therapeutic interventions promote immune organization and coordinated humoral and adaptive immune activation, features associated with improved treatment response (**Fig. 8**). These findings position immune organization as both a baseline marker of adaptive immune competence and a dynamic biomarker of therapy-induced adaptive immune remodeling in breast cancer.

**Fig. 8.**
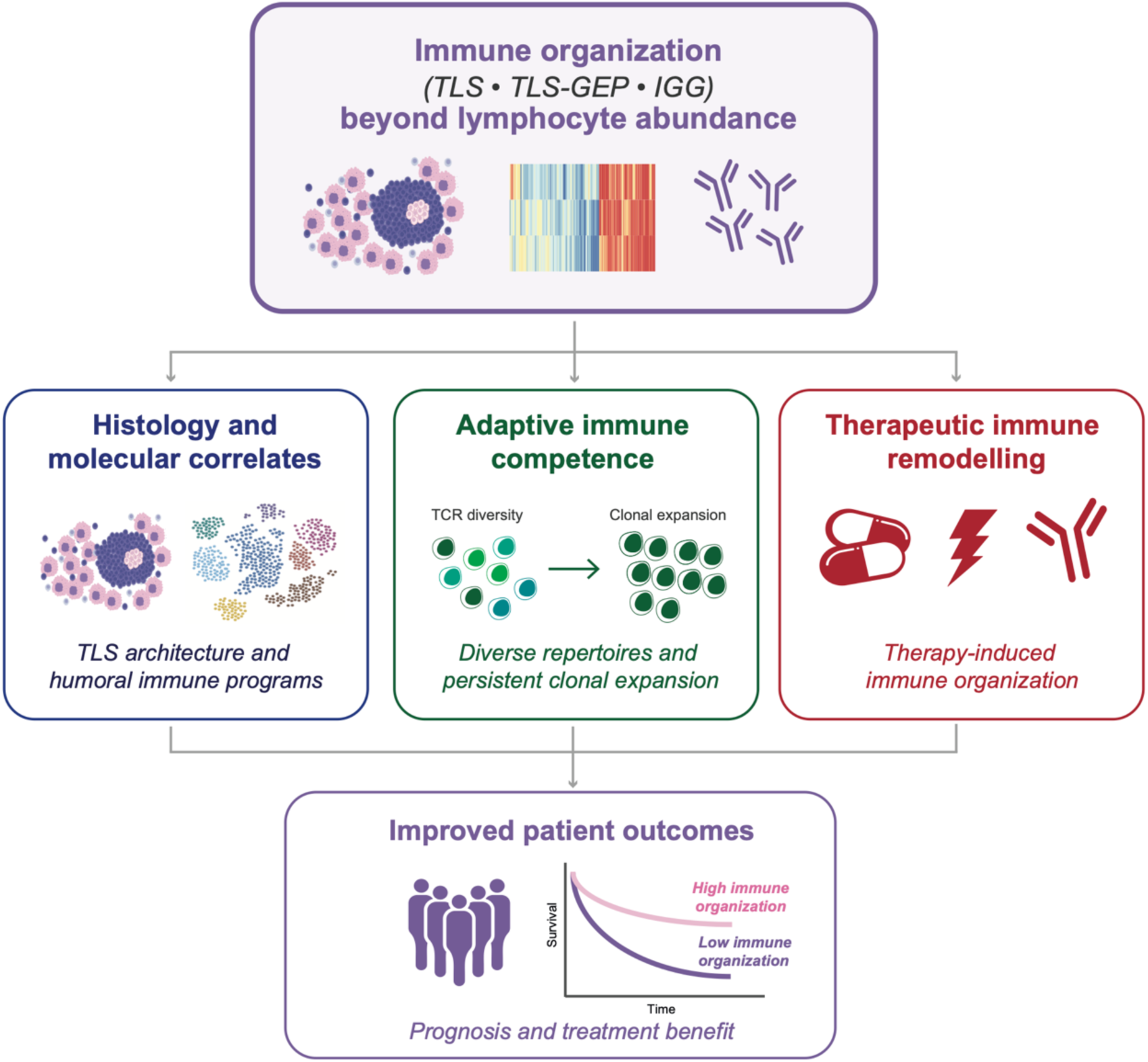
Proposed model of immune organization as a measurable state of adaptive immune competence in breast cancer. Immune organization, reflected by TLS and captured transcriptionally by TLS-GEP and humoral immune programs, represents a biologically distinct dimension of tumor immunity beyond lymphocyte abundance. Organized immune states are characterized by coordinated adaptive immune programs, broader immune repertoires, therapy-induced clonal selection, and preferential expansion of persistent tumor-resident T-cell populations. Dynamic remodeling of immune organization during therapy is associated with improved clinical outcomes, supporting immune organization as a biomarker of adaptive immune competence in breast cancer.

## DISCUSSION

In this study, we identify immune organization as a distinct and clinically relevant dimension of the breast cancer immune microenvironment that is not captured by lymphocyte abundance alone. By integrating histologic TLS assessment with transcriptomic, spatial and TCR repertoire analyses, we show that organized immune states reflect coordinated adaptive immune responses with measurable functional and clinical effects. Across multiple cohorts and orthogonal biological modalities, immune organization consistently provided complementary information beyond lymphocyte abundance and was more closely linked to adaptive immune competence, therapeutic remodeling, and clinical outcome.

A central finding of this work is that immune infiltration and immune organization are related but non-equivalent. Consistent with prior studies, increasing lymphocyte abundance was associated with TLS detection and immune-related transcriptional programs(*2, 4–6, 32–34*). However, this relationship was incomplete. Tumors with high TIL levels frequently lacked organized immune structures, whereas TLS and humoral immune programs were often observed in tumors with low or intermediate infiltration. These discordances indicate that lymphocyte abundance primarily reflects the magnitude of the immune response, whereas immune organization captures its spatial coordination and functional integration into effective adaptive immune responses.

At the transcriptional level, immune organization was characterized by coordinated activation of B-cell, plasma-cell, germinal center, and T-cell programs. Consistent with prior observations(*35, 36*), transcriptional signatures of lymphocyte abundance included genes related to both immune activation and cell proliferation, reflecting the broader biology of inflamed tumors rather than immune organization per se. Humoral immune programs(*23, 27, 28*) emerged as transcriptomic surrogates of this state. Although correlated with TIL levels, IGG showed stronger and more specific associations with TLS presence and maturation, particularly in tumors with low lymphocyte infiltration. Notably, IGG also captured dynamic immune responses under therapy, including clonal selection and expansion. These findings position humoral immune activation as a molecular manifestation of immune organization that extends beyond immune cell density(*14, 16, 24, 25, 28*). Together, these observations indicate that immune organization can be captured across complementary modalities, including histologic TLS assessment, humoral immune programs, and composite models such as TLS-GEP.

Spatial single-cell transcriptomic analyses further showed that organized immune programs extend beyond morphologically defined TLS. While TLS represent highly structured immune niches(*7, 8, 13, 14, 16, 22*), a subset of non-TLS regions displayed coordinated humoral and adaptive immune signatures, particularly in tumors with high IGG expression. These findings support a continuum model of immune organization, with TLS representing a prominent but not exclusive manifestation of spatially coordinated immune activity. Accordingly, morphologically defined TLS should be viewed as one manifestation of a broader organizational state rather than as the sole determinant of immune organization. This framework also explains why transcriptomic measures of immune organization retained prognostic value even in tumors lacking morphologically recognizable TLS.

Consistent with this framework, immune organization was associated with both static and dynamic features of adaptive immunity. At baseline, tumors with organized immune states exhibited more diverse and polyclonal TCR repertoires, consistent with broader adaptive immune engagement. Under therapeutic pressure, immune organization was associated with selective clonal dynamics, including reduced diversity consistent with clonal selection, expansion of specific clonotypes, and preferential expansion of pre-existing tumor-resident clones. Notably, IGG expression, but not TIL levels, predicted these dynamic changes, further supporting immune organization as a marker of effective adaptive immune response. These findings suggest that immune organization reflects not only the presence of immune cells but also their capacity to initiate, sustain, and remodel adaptive immune responses under therapeutic pressure.

The clinical relevance of immune organization was supported across large, independent cohorts. While both lymphocyte abundance- and TLS-related gene expression programs were associated with favorable outcomes in univariable analyses, only immune organization–related signatures retained consistent independent prognostic value in joint models. Importantly, among HR+/HER2-tumors, immune organization retained prognostic value after adjustment for PAM50 Risk of Recurrence (ROR-P), indicating that organized adaptive immune states capture clinically relevant information beyond intrinsic molecular risk. Notably, high lymphocyte abundance in the absence of immune organization did not confer comparable benefit. These findings suggest that the prognostic value attributed to TILs may partly reflect underlying immune organization rather than lymphocyte abundance per se. Collectively, our results indicate that the clinical impact of immune infiltration depends not only on the quantity of immune cells present, but also on their spatial organization and functional coordination. This concept is further supported by recent data from the phase III APHINITY trial, where spatially defined immune aggregates outperformed percentage-based TIL metrics for prognostic discrimination(*37*).

These findings have implications across breast cancer subtypes. In HER2-positive and triple-negative disease, assessment of immune organization may refine patient stratification beyond TILs alone(*2, 5, 6, 25, 26, 30, 33, 34, 38, 39*). In HR+/HER2- tumors, our results identify organized immune responses in a subset of cases, suggesting that immune organization represents an orthogonal biological dimension not captured by existing genomic assays. The observation that immune organization retained prognostic significance after adjustment for PAM50 ROR-P suggests that incorporation of immune organization into existing molecular classifiers could further refine risk stratification. As immunotherapy and other immune-modulating therapies expand across subtypes, immune organization may help identify tumors capable of mounting coordinated and durable immune responses.

From a translational perspective, immune organization can be assessed using complementary histologic and transcriptomic approaches. The H&E-based TLS framework provides a practical and scalable method for routine pathology workflows, consistent with recent efforts toward standardization(*19, 20*). Integration with transcriptomic measures such as the IGG signature may further enhance sensitivity and enable cross-platform implementation. Beyond prognostic stratification, immune organization may serve as a dynamic biomarker of adaptive immune competence and therapeutic responsiveness. Across multiple therapeutic contexts, treatment-induced changes in immune organization were generally more pronounced than changes in immune infiltration and were associated with treatment response, supporting the concept that effective antitumor therapies may preferentially enhance coordinated adaptive immune responses rather than simply increasing lymphocyte abundance. These findings raise the possibility that longitudinal assessment of immune organization could provide a clinically useful framework for monitoring treatment-induced immune remodeling and identifying patients mounting effective antitumor immune responses.

Several limitations should be acknowledged. TLS assessment was based on single sections and may underestimate spatial heterogeneity. Germinal center identification relied on morphology and may introduce variability. TCR analyses support functional engagement but do not establish causal relationships between immune organization, humoral activation, clonal dynamics, and clinical outcome. The derivation of transcriptomic proxies from bulk RNA data may not fully capture spatial heterogeneity of immune organization. Finally, prognostic analyses relied on transcriptomic proxies of immune infiltration and organization rather than direct histologic assessment of TILs and TLS across large cohorts, which may influence the relative performance and comparability of these measures. Future studies integrating longitudinal sampling and functional assays will be needed to further define these relationships.

In summary, immune organization represents a distinct and clinically relevant dimension of tumor immunity in breast cancer. Whereas lymphocyte abundance primarily reflects the extent of immune infiltration, immune organization captures the spatial coordination and adaptive immune competence of antitumor responses, with implications for repertoire diversity, therapeutic immune remodeling, treatment response, and survival. Taken together, our findings support a conceptual framework in which immune organization represents a measurable state of adaptive immune competence that complements both lymphocyte abundance and intrinsic tumor biology. By integrating histologic, transcriptomic, spatial, and immune repertoire analyses, this work links immune organization to coordinated adaptive immune function, therapeutic remodeling, and improved clinical outcomes. These findings provide a foundation for incorporating immune organization into future biomarker strategies aimed at prognosis, treatment monitoring, and therapeutic development in breast cancer.

## MATERIALS AND METHODS

### Study design

This study was designed to evaluate immune organization as a distinct dimension of the tumor immune microenvironment in breast cancer by integrating histopathologic, transcriptomic, spatial, and TCR repertoire analyses. A large discovery cohort with matched histopathologic and gene expression data was used to characterize associations between TLS, TILs, and humoral immune activation. Transcriptomic signatures associated with TILs and TLS were derived and subsequently validated in independent external cohorts with available clinical outcomes. Spatial transcriptomic profiling was performed to assess the spatial distribution of immune programs, and TCR sequencing was used to evaluate functional features of adaptive immune responses and clonal dynamics under therapy. Additional paired pre- and post-treatment datasets were analyzed to evaluate therapy-induced changes in immune organization, immune infiltration, and humoral immune activation.

### Discovery cohort and ethics

We evaluated a cohort of 1,169 breast cancer samples with available histopathologic tumor microenvironment assessment and matched gene expression data generated at the Translational Genomics and Targeted Therapeutics in Solid Tumors Lab (FRCB-IDIBAPS, Barcelona, Spain). Samples were derived from multiple prospective clinical trials conducted by the SOLTI Breast Cancer Research Group—SOLTI-1904 ACROPOLI (n=116)(*40*), SOLTI-1502 ARIANNA (n=35)(*41*), SOLTI-1718 NEREA (n=34)(*42*), SOLTI-1716 TATEN (n=52)(*43*), SOLTI-1503 PROMETEO (n=94)(*44*), and SOLTI-1710 PROMETEO-II (n=63)(*45*)—as well as from an institutional cohort of patients treated at Hospital Clínic Barcelona (n=471) and a real-world cohort of patients who underwent HER2DX testing as part of routine clinical care (Reveal Genomics; n=304)(*46*).

Tumor samples from SOLTI trials and the Hospital Clínic Barcelona cohort were obtained from a shared registered HCB/IDIBAPS biocollection (C.0004038; Spanish National Biobank Registry, ISCIII), whereas samples from Reveal Genomics were obtained from a separate registered biocollection (C.0007805). All patients provided written informed consent for the research use of their tumor samples and for future correlative studies. No identifiable individual-level clinical data were accessed or analyzed. Each contributing cohort was conducted under approval from its corresponding institutional ethics committee. The present study was approved by the Hospital Clínic Barcelona Institutional Ethics Committee (HCB/2025/1142) and conducted in accordance with the Declaration of Helsinki and International Conference on Harmonization Good Clinical Practice guidelines. Material transfer agreements and existing ethical approvals covered the use of all samples included in this study.

### Histopathologic assessment of TILs and TLS

TILs and TLS were retrospectively evaluated on 4-μm formalin-fixed, paraffin-embedded (FFPE) H&E-stained sections by a breast pathologist (E.Sa.) and independently reviewed by a trained postdoctoral researcher (A.M.-R.), with all assessments performed centrally.

TILs were quantified as a continuous variable according to International TILs Working Group recommendations(*47*). TLS were defined as organized, non-encapsulated lymphoid aggregates within the stromal compartment of invasive breast carcinoma. Aggregates were classified as TLS if they contained at least 50 immune cells and were located within 1 mm of the invasive tumor border(*48*). Aggregates outside this region or associated with ductal carcinoma *in situ*, normal mammary epithelium, necrosis, ulceration, or artifacts were excluded.

Germinal centers were identified morphologically on H&E as areas of reduced lymphoid density with organized architecture, reflecting the presence of larger B cells with abundant cytoplasm and a relative decrease in small lymphocytes. Occasional macrophages and vascular structures consistent with high endothelial venules (HEVs) may be observed.

TLS were assessed in both surgical specimens and core biopsies, with identification performed at low magnification (4-10x) and confirmation of germinal centers at higher magnification (20-40×). Detailed guidelines for TLS identification and assessment are provided in **Supplementary Methods.**

### Gene expression analysis

RNA was extracted from the same FFPE tumor blocks used for histopathologic evaluation using the High Pure FFPET RNA isolation kit (Roche). Between one and eight 10-μm sections were used per sample depending on tumor cellularity, and macrodissection was performed when necessary to minimize normal tissue contamination.

A minimum of 100 ng of RNA was analyzed using a 192-gene custom panel including PAM50, HER2DX(*27*), TNBCDX(*49*), and additional immune-related and housekeeping genes(*27*). For each sample, the HER2DX IGG signature score was calculated as previously described(*27*), based on the expression of immunoglobulin-related genes reflecting humoral immune activation, including *CD27, CD79A, HLA-C, IGJ, IGKC, IGL, IGLV3-25, IL2RG, CXCL8, LAX1, NTN3, PIM2, POU2AF1,* and *TNFRSF17*(*23*).

### Derivation of transcriptomic signatures

Cases with available histopathologic annotations were used to derive gene expression signatures associated with high TIL levels (≥30%) and mature TLS (TLS2a/2b). Differential gene expression analysis was performed using Significance Analysis of Microarrays (SAM) with two-class unpaired comparisons, and genes were selected based on fold change greater than 1 and false discovery rate q <0.01.

Signature derivation was performed using least absolute shrinkage and selection operator (LASSO) regression to enable variable selection and shrinkage. Final signature scores were calculated as weighted sums of gene expression values using LASSO-derived coefficients. The TLS-associated gene expression signature included 11 genes (i.e., *CD79A, ITK, IL2RG, MUCL1, BOC, MYC, IRF1, EAF2, CD84, KLRD1, CXCL8*) and the TIL-associated signature included 30 genes (i.e., *ANLN, BOC, CCNB2, CCNE1, CD19, CD3D, CD7, CD79A, CDC6, CTLA4, CXCL13, EOMES, GPNMB, GRB7, GSDMB, IGL, IGLV3.25, IL18R1, IL2RG, MMP1, MRAS, MYC, PTTG1, RAD51, S100A9, SF3A1, STAT1, TMEM45B, TOP2A, TRPV6*).

### Independent validation and on-treatment cohorts

Validation analyses were conducted using TCGA-BRCA(*50*), METABRIC(*51*), and SCAN-B(*52, 53*) datasets. Gene expression, clinical subtype, PAM50 Risk of Recurrence (ROR-P), and overall survival data were available for 1,035 TCGA samples, 1,985 METABRIC samples, and 6,655 SCAN-B samples. In TCGA-BRCA, histopathologic annotations of TILs and TLS derived from H&E evaluation were obtained from previously published data(*12*). Overall survival (OS) was defined as the time from diagnosis to death from any cause or last follow-up.

To evaluate therapy-induced immune remodeling, paired pre- and post-treatment tumor samples from multiple window-of-opportunity and neoadjuvant studies were analyzed (n denotes paired cases), including SOLTI-1503 PROMETEO (T-VEC plus atezolizumab, n=18)(*44*), SOLTI-1710 PROMETEO II (CDK4/6 inhibitor plus endocrine therapy, n=20)(*45*), YOUNGSTER (radiotherapy boost, n=13)(*54*), BIONHER (trastuzumab plus pertuzumab, n=63; NCT05912062), SOLTI-1905 ELIPSE (elacestrant, n=22)(*55*), SOLTI-1402 CORALLEEN (CDK4/6 inhibitor plus endocrine therapy, n=29, and chemotherapy, n=35)(*56*), and a short (2-12 weeks) preoperative endocrine therapy HCB cohort (n=93)(*57*). Histologic TLS and TIL assessments on H&E-stained sections were available for PROMETEO, PROMETEO-II, and the preoperative endocrine therapy HCB cohort, whereas paired gene expression data were available across all cohorts.

Gene expression data were generated using RNA sequencing, microarray, or nCounter-based platforms and were normalized prior to analysis. TLS-GEP, TIL-GEP, and HER2DX IGG scores were calculated using predefined coefficients, standardized within each dataset, and analyzed as continuous variables. For paired analyses, changes from baseline (Δ) were calculated using standardized signature scores.

### Spatial transcriptomics

Spatial transcriptomic profiling was performed in eight breast tumor samples from newly diagnosed patients, obtained from core needle biopsies, using the 10x Genomics Xenium platform with the Xenium Prime 5k panel supplemented by a custom gene panel. Tissue sections were processed according to manufacturer guidelines, followed by H&E staining and image alignment using Xenium Explorer 4. Regions of interest were defined, and gene expression was quantified within each region. Seventeen gene modules were defined per ROI based on the mean expression of predefined marker genes (**Table S9**), including immune, stromal, tumor, TLS, germinal center, plasma cell, and IGG-related programs.

### TCR and BCR repertoire analysis

To assess functional features of adaptive immunity, TCR and B-cell receptor (BCR) repertoire sequencing was performed in paired tumor samples obtained before and after neoadjuvant therapy. RNA was extracted from tumor samples and peripheral blood mononuclear cells, and TCRβ and immunoglobulin heavy chain (IGH) sequencing were performed using Illumina-based platforms. Complementarity-determining region 3 sequences were analyzed to quantify repertoire features.

For TCR analyses, repertoire metrics included the number of productive clonotypes, Shannon entropy, the proportion of hyperexpanded clones, and the presence of persistent clonotypes detected both pre- and post-treatment. Clonal expansion was defined using fold-change thresholds and contracting and newly detected clonotypes were identified. Newly detected clonotypes were defined as those not observed in baseline tumor samples but present in post-treatment samples. To explore the origin of these clonotypes, TCR repertoires from serial peripheral blood mononuclear cell samples were analyzed in a subset of patients and overlap between tumor and blood clonotypes was assessed. Alluvial plots were generated to visualize clonal dynamics.

For BCR analyses, analogous metrics were computed, including clonotype number, diversity, and clonal dominance based on IGH sequences. These analyses were used to characterize humoral immune repertoire diversity and its relationship with TLS presence and IGG signature expression.

### Statistical analysis

Associations between clinicopathologic variables, TILs, TLS categories, and the HER2DX IGG signature were evaluated using descriptive statistics and two-sided tests. Multivariable logistic regression models were used to assess associations between TLS presence and maturation and the HER2DX IGG signature, TIL levels treated as a continuous variable, and tumor site coded as primary versus metastatic. An interaction term between IGG and TIL levels was included to assess effect modification, and results are reported as odds ratios with 95% confidence intervals. Receiver operating characteristic analyses were performed to evaluate predictive performance, and areas under the curve were calculated. To account for multiple testing, p-values were adjusted using the Benjamini-Hochberg FDR method where applicable. Interobserver reproducibility of TLS classification was assessed using percentage agreement.

In validation cohorts, correlations were assessed using Spearman coefficients, and associations between gene expression signatures and histopathologic features were evaluated using two-sided Wilcoxon rank-sum tests. Survival analyses were performed using Cox proportional hazards models with overall survival as the endpoint. Multivariable models were adjusted for clinical subtype and, when indicated, PAM50 Risk of Recurrence (ROR-P). Hazard ratios were reported per standard deviation increase in signature scores. Kaplan-Meier curves were generated using median cutoffs and compared using the log-rank test.

For TCR and BCR repertoire analyses, non-parametric methods were used due to non-normal data distributions. Comparisons were performed using Wilcoxon or Kruskal–Wallis tests, and correlations using Spearman coefficients. To assess whether associations between immune organization and TCR repertoire features were independent of lymphocyte abundance, multivariable linear models were used with TCR diversity metrics, including Shannon entropy, as dependent variables and TLS status as the main predictor, adjusting for TIL levels as a continuous variable. Predefined thresholds were used for categorical analyses, including TIL-high defined as ≥30% stromal TILs and clonal expansion defined as a >2-fold increase in clonotype frequency between pre- and post-treatment samples.

For paired on-treatment analyses, changes from baseline (Δ) in TLS-GEP, TIL-GEP, and HER2DX IGG scores were calculated using standardized signature scores. Within each study, changes from baseline and paired comparisons between ΔTLS-GEP and ΔTIL-GEP were evaluated using Wilcoxon signed-rank tests. Associations between treatment-induced changes in immune signatures were assessed using Spearman correlation. Logistic regression models adjusted for study were used to evaluate associations between treatment-induced changes in immune signatures and treatment response. Additional models simultaneously included ΔTLS-GEP and ΔTIL-GEP to assess their independent associations with treatment response.

All statistical tests were two-sided, and P<0.05 was considered statistically significant. Analyses were performed using R software (version 4.2.3). Cases with missing data for specific variables were excluded from the corresponding analyses. No statistical methods were used to predetermine sample size.

## Data Availability

The data generated in this study, including pathological annotations and gene expression data, are available from the corresponding authors upon reasonable request, subject to institutional approval and ethical regulations governing patient data. Publicly available datasets analyzed in this study are accessible through their respective repositories.

## Supplemental information

- Document S1: Figures: S1-S12
- Document S2: Tables S1-S12
- Document S3: Supplementary Methods

## Acknowledgments

We thank all patients who contributed samples to this study and the clinical teams involved in patient care and sample collection. We also acknowledge the SOLTI Breast Cancer Research Group and Reveal Genomics for providing access to clinical trial and real-world cohorts included in this study. We gratefully acknowledge the IRB Barcelona Advanced Genomic Technologies Core Facility for their expert technical assistance with the Xenium spatial transcriptomics experiments. Preliminary results of this study were presented at the ESMO Breast Cancer Congress 2025 as a poster communication(*58*).

## Funding

Fundación científica AECC Ayudas Investigador AECC 2021 (INVES21943BRAS) (FBM) Fundación CRIS contra el cáncer PR_EX_2021-14 and CRIS Dame5mas (AP)

Agència de Gestó d’Ajuts Universitaris i de Recerca 2021 SGR 01156 (AP) Fundación Fero BECA ONCOXXI21 (AP)

Instituto de Salud Carlos III (ISCIII) PI22/01017, co-funded by the European Union (AP) Asociación Cáncer de Mama Metastásico IV Premios M. Chiara Giorgetti (AP)

Breast Cancer Research Foundation BCRF-22-198, BCRF-23-198 and BCRF-24-198 (AP)

European Union’s Horizon 2020 Research and Innovation Programme (RESCUER; grant 847912) (AP)

Fundación Científica Asociación Española Contra el Cáncer AECC-IDIBAPS Excellence Program EPAEC246711CLIN (AP)

SEOM Research Fellowship 2024 (ESe)

CRIS Out-Back Fellowship Programme 2024 (ESe)

Fundación Científica de la Asociación Española Contra el Cáncer (AECC Clínico Junior Grant) (VA)

No project-specific funding was received for this work.

## Author contributions

Conceptualization: ESa, ESe, AMR, VA, FBM, AP

Methodology: ESa, ESe, AMR, VA, FBM, AP

Investigation: ESa, ESe, AMR, VA, TP, MM, OMS, RGB, IGF, ARH, BW, PG, LA, CRP, CS, MO, ECi, LM, SP, AGW, SMT, AMS, JMFC, ECa, JT, FBM, AP

Data curation: ESa, ESe, AMR, VA, LP, JSP, PV

Formal analysis: ESa, ESe, AMR, VA, LP, JSP, PV, FBM, AP

Visualization: ESa, ESe, AMR, VA

Funding acquisition: FBM, ESe, VA, AP

Project administration: AP

Supervision: FBM, AP

Writing – original draft: ESa, ESe, AMR, VA, FBM, AP

Writing – review & editing: All authors

## Resource availability

### Lead contact

Further information and requests for resources should be directed to and will be fulfilled by the corresponding authors.

## Materials availability

This study did not generate new unique reagents or materials.

## Data and code availability

The data generated in this study, including pathological annotations and gene expression data, are available from the corresponding authors upon reasonable request and subject to institutional and ethical regulations governing patient data. All other data supporting the findings of this study are available within the article and its supplementary materials. No custom code was generated in this study.

## References

1. T. C. Bruno, New predictors for immunotherapy responses sharpen our view of the tumour microenvironment. Nature 577, 474–476 (2020).

2. S. Loi, Tumor-infiltrating lymphocytes, breast cancer subtypes and therapeutic efficacy. OncoImmunology 2, e24720 (2013).

3. R. Salgado, C. Denkert, S. Demaria, N. Sirtaine, F. Klauschen, G. Pruneri, S. Wienert, G. Van den Eynden, F. L. Baehner, F. Penault-Llorca, E. A. Perez, E. A. Thompson, W. F. Symmans, A. L. Richardson, J. Brock, C. Criscitiello, H. Bailey, M. Ignatiadis, G. Floris, J. Sparano, Z. Kos, T. Nielsen, D. L. Rimm, K. H. Allison, J. S. Reis-Filho, S. Loibl, C. Sotiriou, G. Viale, S. Badve, S. Adams, K. Willard-Gallo, S. Loi, The evaluation of tumor-infiltrating lymphocytes (TILs) in breast cancer: recommendations by an International TILs Working Group 2014. Annals of Oncology 26, 259–271 (2015).

4. M. V. Dieci, N. Radosevic-Robin, S. Fineberg, G. van den Eynden, N. Ternes, F. Penault-Llorca, G. Pruneri, T. M. D’Alfonso, S. Demaria, C. Castaneda, J. Sanchez, S. Badve, S. Michiels, V. Bossuyt, F. Rojo, B. Singh, T. Nielsen, G. Viale, S.-R. Kim, S. Hewitt, S. Wienert, S. Loibl, D. Rimm, F. Symmans, C. Denkert, S. Adams, S. Loi, R. Salgado, Update on tumor-infiltrating lymphocytes (TILs) in breast cancer, including recommendations to assess TILs in residual disease after neoadjuvant therapy and in carcinoma in situ: A report of the International Immuno-Oncology Biomarker Working Group on Breast Cancer. Seminars in Cancer Biology 52, 16–25 (2018).

5. S. Loi, R. Salgado, S. Adams, G. Pruneri, P. A. Francis, M. Lacroix-Triki, H. Joensuu, M. V. Dieci, S. Badve, S. Demaria, R. Gray, E. Munzone, D. Drubay, J. Lemonnier, C. Sotiriou, P. L. Kellokumpu-Lehtinen, A. Vingiani, K. Gray, F. André, C. Denkert, M. Piccart, E. Roblin, S. Michiels, Tumor infiltrating lymphocyte stratification of prognostic staging of early-stage triple negative breast cancer. npj Breast Cancer 8, 3 (2022).

6. S. Loi, D. Drubay, S. Adams, G. Pruneri, P. A. Francis, M. Lacroix-Triki, H. Joensuu, M. V. Dieci, S. Badve, S. Demaria, R. Gray, E. Munzone, J. Lemonnier, C. Sotiriou, M. J. Piccart, P.-L. Kellokumpu-Lehtinen, A. Vingiani, K. Gray, F. Andre, C. Denkert, R. Salgado, S. Michiels, Tumor-Infiltrating Lymphocytes and Prognosis: A Pooled Individual Patient Analysis of Early-Stage Triple-Negative Breast Cancers. Journal of Clinical Oncology 37, 559–569 (2019).

7. J.-L. Teillaud, A. Houel, M. Panouillot, C. Riffard, M.-C. Dieu-Nosjean, Tertiary lymphoid structures in anticancer immunity. Nature Reviews Cancer 24, 629–646 (2024).

8. W. H. Fridman, M. Meylan, G. Pupier, A. Calvez, I. Hernandez, C. Sautès-Fridman, Tertiary lymphoid structures and B cells: An intratumoral immunity cycle. Immunity 56, 2254–2269 (2023).

9. B. Goeppert, L. Frauenschuh, M. Zucknick, A. Stenzinger, M. Andrulis, F. Klauschen, K. Joehrens, A. Warth, M. Renner, A. Mehrabi, M. Hafezi, A. Thelen, P. Schirmacher, W. Weichert, Prognostic impact of tumour-infiltrating immune cells on biliary tract cancer. British Journal of Cancer 109, 2665–2674 (2013).

10. M. Garnelo, A. Tan, Z. Her, J. Yeong, C. J. Lim, J. Chen, K. H. Lim, A. Weber, P. Chow, A. Chung, L. L. P. Ooi, H. C. Toh, M. Heikenwalder, I. O. L. Ng, A. Nardin, Q. Chen, J.- P. Abastado, V. Chew, Interaction between tumour-infiltrating B cells and T cells controls the progression of hepatocellular carcinoma. Gut 66, 342–351 (2017).

11. K. Garg, M. Maurer, J. Griss, M.-C. Brüggen, I. H. Wolf, C. Wagner, N. Willi, K. D. Mertz, S. N. Wagner, Tumor-associated B cells in cutaneous primary melanoma and improved clinical outcome. Human Pathology 54, 157–164 (2016).

12. Y. J. Cha, C. E. O’Connell, B. C. Calhoun, B. M. Felsheim, A. Fernandez-Martinez, C. Fan, C. Brueffer, C. Larsson, Å. Borg, L. H. Saal, C. M. Perou, Genomic Characteristics Related to Histology-Based Immune Features in Breast Cancer. Modern Pathology 38, (2025).

13. R. Cabrita, M. Lauss, A. Sanna, M. Donia, M. Skaarup Larsen, S. Mitra, I. Johansson, B. Phung, K. Harbst, J. Vallon-Christersson, A. van Schoiack, K. Lövgren, S. Warren, K. Jirström, H. Olsson, K. Pietras, C. Ingvar, K. Isaksson, D. Schadendorf, H. Schmidt, L. Bastholt, A. Carneiro, J. A. Wargo, I. M. Svane, G. Jönsson, Tertiary lymphoid structures improve immunotherapy and survival in melanoma. Nature 577, 561–565 (2020).

14. F. Petitprez, A. de Reyniès, E. Z. Keung, T. W.-W. Chen, C.-M. Sun, J. Calderaro, Y.-M. Jeng, L.-P. Hsiao, L. Lacroix, A. Bougoüin, M. Moreira, G. Lacroix, I. Natario, J. Adam, C. Lucchesi, Y. h. Laizet, M. Toulmonde, M. A. Burgess, V. Bolejack, D. Reinke, K. M. Wani, W.-L. Wang, A. J. Lazar, C. L. Roland, J. A. Wargo, A. Italiano, C. Sautès-Fridman, H. A. Tawbi, W. H. Fridman, B cells are associated with survival and immunotherapy response in sarcoma. Nature 577, 556–560 (2020).

15. L. Vanhersecke, M. Brunet, J.-P. Guégan, C. Rey, A. Bougouin, S. Cousin, S. Le Moulec, B. Besse, Y. Loriot, M. Larroquette, I. Soubeyran, M. Toulmonde, G. Roubaud, S. Pernot, M. Cabart, F. Chomy, C. Lefevre, K. Bourcier, M. Kind, I. Giglioli, C. Sautès-Fridman, V. Velasco, F. Courgeon, E. Oflazoglu, A. Savina, A. Marabelle, J.-C. Soria, C. Bellera, C. Sofeu, A. Bessede, W. H. Fridman, F. Le Loarer, A. Italiano, Mature tertiary lymphoid structures predict immune checkpoint inhibitor efficacy in solid tumors independently of PD-L1 expression. Nature Cancer 2, 794–802 (2021).

16. B. A. Helmink, S. M. Reddy, J. Gao, S. Zhang, R. Basar, R. Thakur, K. Yizhak, M. Sade-Feldman, J. Blando, G. Han, V. Gopalakrishnan, Y. Xi, H. Zhao, R. N. Amaria, H. A. Tawbi, A. P. Cogdill, W. Liu, V. S. LeBleu, F. G. Kugeratski, S. Patel, M. A. Davies, P. Hwu, J. E. Lee, J. E. Gershenwald, A. Lucci, R. Arora, S. Woodman, E. Z. Keung, P.-O. Gaudreau, A. Reuben, C. N. Spencer, E. M. Burton, L. E. Haydu, A. J. Lazar, R. Zapassodi, C. W. Hudgens, D. A. Ledesma, S. Ong, M. Bailey, S. Warren, D. Rao, O. Krijgsman, E. A. Rozeman, D. Peeper, C. U. Blank, T. N. Schumacher, L. H. Butterfield, M. A. Zelazowska, K. M. McBride, R. Kalluri, J. Allison, F. Petitprez, W. H. Fridman, C. Sautès-Fridman, N. Hacohen, K. Rezvani, P. Sharma, M. T. Tetzlaff, L. Wang, J. A. Wargo, B cells and tertiary lymphoid structures promote immunotherapy response. Nature 577, 549–555 (2020).

17. P. Tsou, H. Katayama, E. J. Ostrin, S. M. Hanash, The Emerging Role of B Cells in Tumor Immunity. Cancer Research 76, 5597–5601 (2016).

18. B. Wang, J. Liu, Y. Han, Y. Deng, J. Li, Y. Jiang, The Presence of Tertiary Lymphoid Structures Provides New Insight Into the Clinicopathological Features and Prognosis of Patients With Breast Cancer. Frontiers in Immunology Volume 13 - 2022, (2022).

19. L. Buisseret, C. Desmedt, S. Garaud, M. Fornili, X. Wang, G. Van den Eyden, A. de Wind, S. Duquenne, A. Boisson, C. Naveaux, F. Rothé, S. Rorive, C. Decaestecker, D. Larsimont, M. Piccart-Gebhart, E. Biganzoli, C. Sotiriou, K. Willard-Gallo, Reliability of tumor-infiltrating lymphocyte and tertiary lymphoid structure assessment in human breast cancer. Modern Pathology 30, 1204–1212 (2017).

20. L. Vanhersecke, A. Bougouin, A. Crombé, M. Brunet, C. Sofeu, M. Parrens, H. Pierron, B. Bonhomme, N. Lembege, C. Rey, V. Velasco, I. Soubeyran, H. Begueret, A. Bessede, C. Bellera, J.-Y. Scoazec, A. Italiano, C. S. Fridman, W. H. Fridman, F. Le Loarer, Standardized Pathology Screening of Mature Tertiary Lymphoid Structures in Cancers. Laboratory Investigation 103, (2023).

21. Y. Zhang, H. Chen, H. Mo, X. Hu, R. Gao, Y. Zhao, B. Liu, L. Niu, X. Sun, X. Yu, Y. Wang, Q. Chang, T. Gong, X. Guan, T. Hu, T. Qian, B. Xu, F. Ma, Z. Zhang, Z. Liu, Single-cell analyses reveal key immune cell subsets associated with response to PD-L1 blockade in triple-negative breast cancer. Cancer Cell 39, 1578–1593.e1578 (2021).

22. D. Narvaez, J. Nadal, A. Nervo, M. V. Costanzo, C. Paletta, F. E. Petracci, S. Rivero, A. Ostinelli, B. Freile, D. Enrico, M. T. Pombo, M. Amat, E. D. Aguirre, M. Chacon, F. Waisberg, The Emerging Role of Tertiary Lymphoid Structures in Breast Cancer: A Narrative Review. Cancers 16, 396 (2024).

23. C. Fan, A. Prat, J. S. Parker, Y. Liu, L. A. Carey, M. A. Troester, C. M. Perou, Building prognostic models for breast cancer patients using clinical variables and hundreds of gene expression signatures. BMC Medical Genomics 4, 3 (2011).

24. M. D. Iglesia, B. G. Vincent, J. S. Parker, K. A. Hoadley, L. A. Carey, C. M. Perou, J. S. Serody, Prognostic B-cell Signatures Using mRNA-Seq in Patients with Subtype-Specific Breast and Ovarian Cancer. Clinical Cancer Research 20, 3818–3829 (2014).

25. A. Fernandez-Martinez, T. Pascual, B. Singh, P. Nuciforo, N. U. Rashid, K. V. Ballman, J. D. Campbell, K. A. Hoadley, P. A. Spears, L. Pare, F. Brasó-Maristany, N. Chic, I. Krop, A. Partridge, J. Cortés, A. Llombart-Cussac, A. Prat, C. M. Perou, L. A. Carey, Prognostic and Predictive Value of Immune-Related Gene Expression Signatures vs Tumor-Infiltrating Lymphocytes in Early-Stage ERBB2/HER2-Positive Breast Cancer: A Correlative Analysis of the CALGB 40601 and PAMELA Trials. JAMA Oncology 9, 490–499 (2023).

26. A. Fernandez-Martinez, I. E. Krop, D. W. Hillman, M.-Y. Polley, J. S. Parker, L. Huebner, K. A. Hoadley, J. Shepherd, S. Tolaney, N. L. Henry, C. Dang, L. Harris, D. Berry, O. Hahn, C. Hudis, E. Winer, A. Partridge, C. M. Perou, L. A. Carey, Survival, Pathologic Response, and Genomics in CALGB 40601 (Alliance), a Neoadjuvant Phase III Trial of Paclitaxel-Trastuzumab With or Without Lapatinib in HER2-Positive Breast Cancer. Journal of Clinical Oncology 38, 4184–4193 (2020).

27. A. Prat, V. Guarneri, T. Pascual, F. Brasó-Maristany, E. Sanfeliu, L. Paré, F. Schettini, D. Martínez, P. Jares, G. Griguolo, M. V. Dieci, J. Cortés, A. Llombart-Cussac, B. Conte, M. Marín-Aguilera, N. Chic, J. A. Puig-Butillé, A. Martínez, P. Galván, Y.-H. Tsai, B. González-Farré, A. Mira, A. Vivancos, P. Villagrasa, J. S. Parker, P. Conte, C. M. Perou, Development and validation of the new HER2DX assay for predicting pathological response and survival outcome in early-stage HER2-positive breast cancer. eBioMedicine 75, (2022).

28. L. Angelats, L. Paré, C. Rubio-Perez, E. Sanfeliu, A. González, E. Seguí, G. Villacampa, M. Marín-Aguilera, S. Pernas, B. Conte, V. Albarrán-Fernández, O. Martínez-Sáez, Á. Aguirre, P. Galván, A. Fernandez-Martinez, S. Cobo, M. Rey, A. Martínez-Romero, B. Walbaum, F. Schettini, M. Vidal, W. Buckingham, M. Muñoz, B. Adamo, Y. Agrawal, S. Guedan, T. Pascual, J. Agudo, M. Grzelak, N. Borcherding, H. Heyn, A. Vivancos, J. S. Parker, P. Villagrasa, C. M. Perou, A. Prat, F. Brasó-Maristany, Linking tumor immune infiltration to enhanced longevity in recurrence-free breast cancer. ESMO Open 10, (2025).

29. G. Villacampa, T. Pascual, P. Tarantino, J. Cortés, J. Perez-García, A. Llombart-Cussac, P. Conte, M. Mancino, V. Guarneri, M. V. Dieci, A. G. Waks, F. Schettini, F. Brasó-Maristany, G. Griguolo, B. A. de Castro, C. Reboredo, S. Antolín, C. Bueno-Muiño, I. Echavarría, S. López-Tarruella, T. Massarrah, M. del Monte-Millán, M. Martín, W. Buckingham, J. S. Parker, A. Vivancos, K. Polyak, O. M. Filho, A. C. Wolff, A. DeMichele, N. M. Tung, C. M. Perou, L. Paré, P. Villagrasa, A. Prat, S. M. Tolaney, HER2DX and survival outcomes in early-stage HER2-positive breast cancer: an individual patient-level meta-analysis. The Lancet Oncology 26, 1100–1112 (2025).

30. K. Nozawa, M. Sawaki, Y. Uemura, M. Tsuneizumi, T. Takano, N. Gondo, F. Hara, M. Harao, T. Toyama, N. Taira, A. Vivancos, C. M. Perou, E. Sanfeliu, F. Brasó-Maristany, J. S. Parker, W. Buckingham, L. Paré, G. Villacampa, M. Marín-Aguilera, P. Villagrasa, A. Prat, H. Iwata, HER2DX in older patients with HER2-positive early breast cancer: extended follow-up from the RESPECT trial of trastuzumab ± chemotherapy. Nature Communications 16, 9585 (2025).

31. G. Villacampa, N. M. Tung, S. Pernas, L. Paré, C. Bueno-Muiño, I. Echavarría, S. López-Tarruella, M. Roche-Molina, M. del Monte-Millán, M. Marín-Aguilera, F. Brasó-Maristany, A. G. Waks, T. Pascual, O. Martínez-Sáez, A. Vivancos, P. F. Conte, V. Guarneri, M. Vittoria Dieci, G. Griguolo, J. Cortés, A. Llombart-Cussac, M. Muñoz, M. Vidal, B. Adamo, A. C. Wolff, A. DeMichele, P. Villagrasa, J. S. Parker, C. M. Perou, A. Fernandez-Martinez, L. A. Carey, E. A. Mittendorf, M. Martín, A. Prat, S. M. Tolaney, Association of HER2DX with pathological complete response and survival outcomes in HER2-positive breast cancer. Annals of Oncology 34, 783–795 (2023).

32. A. Prat, V. Guarneri, L. Paré, G. Griguolo, T. Pascual, M. V. Dieci, N. Chic, B. González-Farré, A. Frassoldati, E. Sanfeliu, J. M. Cejalvo, M. Muñoz, G. Bisagni, F. Brasó-Maristany, L. Urso, M. Vidal, A. A. Brandes, B. Adamo, A. Musolino, F. Miglietta, B. Conte, M. Oliveira, C. Saura, S. Pernas, J. Alarcón, A. Llombart-Cussac, J. Cortés, L. Manso, R. López, E. Ciruelos, F. Schettini, P. Villagrasa, L. A. Carey, C. M. Perou, F. Piacentini, R. D’Amico, E. Tagliafico, J. S. Parker, P. Conte, A multivariable prognostic score to guide systemic therapy in early-stage HER2-positive breast cancer: a retrospective study with an external evaluation. The Lancet Oncology 21, 1455–1464 (2020).

33. P. Nuciforo, T. Pascual, J. Cortés, A. Llombart-Cussac, R. Fasani, L. Paré, M. Oliveira, P. Galvan, N. Martínez, B. Bermejo, M. Vidal, S. Pernas, R. López, M. Muñoz, I. Garau, L. Manso, J. Alarcón, E. Martínez, V. Rodrik-Outmezguine, J. C. Brase, P. Villagrasa, A. Prat, E. Holgado, A predictive model of pathologic response based on tumor cellularity and tumor-infiltrating lymphocytes (CelTIL) in HER2-positive breast cancer treated with chemo-free dual HER2 blockade. Annals of Oncology 29, 170–177 (2018).

34. N. Chic, S. J. Luen, P. Nuciforo, R. Salgado, D. Fumagalli, F. Hilbers, Y. Wang, E. de Azambuja, I. Láng, S. Di Cosimo, C. Saura, J. Huober, A. Prat, S. Loi, Tumor Cellularity and Infiltrating Lymphocytes as a Survival Surrogate in HER2-Positive Breast Cancer. JNCI: Journal of the National Cancer Institute 114, 467–470 (2021).

35. H. Locy, S. Verhulst, W. Cools, W. Waelput, S. Brock, L. Cras, A. Schiettecatte, J. Jonckheere, L. A. van Grunsven, M. Vanhoeij, K. Thielemans, K. Breckpot, Assessing Tumor-Infiltrating Lymphocytes in Breast Cancer: A Proposal for Combining Immunohistochemistry and Gene Expression Analysis to Refine Scoring. Front Immunol 13, 794175 (2022).

36. M. Kochi, T. Iwamoto, N. Niikura, G. Bianchini, S. Masuda, T. Mizoo, T. Nogami, T. Shien, T. Motoki, N. Taira, Y. Tokuda, H. Doihara, J. Matsuoka, T. Fujiwara, Tumour-infiltrating lymphocytes (TILs)-related genomic signature predicts chemotherapy response in breast cancer. Breast Cancer Research and Treatment 167, 39–47 (2018).

37. R. Salgado, L. E. Lara Gonzalez, F. Giudici, F. Rojo, L. Comerma, S. Wienert, J. Palacios, Z. Kos, S. L. De Haas, A. Rodriguez Lescure, paper presented at the San Antonio Breast Cancer Symposium 2025, San Antonio, TX, USA, 2025.

38. I. Schlam, S. Loi, R. Salgado, S. M. Swain, Tumor-infiltrating lymphocytes in HER2-positive breast cancer: potential impact and challenges. ESMO Open 10, (2025).

39. S. M. Tolaney, P. Tarantino, N. Graham, N. Tayob, L. Parè, G. Villacampa, C. T. Dang, D. A. Yardley, B. Moy, P. K. Marcom, K. S. Albain, H. S. Rugo, M. J. Ellis, I. Shapira, A. C. Wolff, L. A. Carey, R. Barroso-Sousa, P. Villagrasa, M. DeMeo, M. DiLullo, J. G. T. Zanudo, J. Weiss, N. Wagle, A. H. Partridge, A. G. Waks, C. A. Hudis, I. E. Krop, H. J. Burstein, A. Prat, E. P. Winer, Adjuvant paclitaxel and trastuzumab for node-negative, HER2-positive breast cancer: final 10-year analysis of the open-label, single-arm, phase 2 APT trial. The Lancet Oncology 24, 273–285 (2023).

40. E. Seguí, F. Brasó-Maristany, T. Pascual, E. Sanfeliu, I. Victoria, C. Saura, C. Hierro, A. López-González, Y. Izarzugaza, E. Ciruelos, J. Gavilá, F. Racca, J. M. Cejalvo, K. Amillano, L. Paz-Ares, M. Juan, E. Felip, E. Garralda, B. González, A. Arance, J. Martín-Liberal, B. Conte, B. Walbaum, A. Indacochea, O. Castillo, P. Blasco, F. Pardo, Á. Aguirre, V. Sirenko, X. González, P. Galván, A. Vivancos, J. M. Ferrero-Cafiero, A. Mulero-Sánchez, F. Salvador, G. Villacampa, R. Mesía, A. Cervantes, A. Prat, J. Tabernero, Efficacy of anti-PD1 therapy in PD1-high mRNA tumors across multiple cancer types: results from cohort 1 and cohort 2 of the phase II SOLTI-1904 ACROPOLI trial. ESMO Open 10, 105838 (2025).

41. M. Oliveira, T. Pascual, S. Pernas, M. Margelí, S. Blanch, B. Adamo, F. J. S. Bofill, X. Gonzalez-Farré, S. Chillara, P. Galván, E. Sanfeliu, P. Blasco, V. Sirenko, A. Espinosa, C. M. Perou, A. Prat, L. Manso, Abstract PO3-05-06: Targeting PAM50 HER2-Enriched intrinsic subtype with enzalutamide in hormone receptor-positive/HER2-negative (HR+/HER2-) advanced breast cancer: results of the SOLTI-1502 ARIANNA trial. Cancer Research 84, PO3–05–06–PO03–05–06 (2024).

42. E. Ciruelos, C. Saura, X. Gonzalez-Farré, F. J. S. Bofill, M. Vidal, I. Blancas, E. López-Miranda, M. Iglesias, M. Arumí, M. Margelí, C. Pulido, S. M. Murillo, F. Henao, P. Sanchez, S. Alves, A. G. Ortiz, J. P. Coelho, S. Escrivá-de-Romaní, A. Espinosa, T. Pascual, A. Prat, Abstract PO3-05-07: SOLTI-1718 NEREA Trial: Neratinib in hormone receptor (HR)-positive/HER2-negative HER2-enriched (HER2-E) advanced breast cancer (BC). Cancer Research 84, PO3–05–07–PO03–05–07 (2024).

43. B. Conte, F. Brasó-Maristany, T. Pascual, C. Hernando, S. Vázquez, S. Blanch, M. Oliveira, J. A. Virizuela, M. Muñoz, E. Seguí, A. Rodriguez Hernández, M. J. Vidal Losada, P. Galván, O. Castillo, P. Blasco, M. Alva, N. Chic, E. Sanfeliu, S. Cano-Crespo, F. Salvador, G. Villacampa, L. Villanueva, J. M. Ferrero-Cafiero, A. Vivancos, A. Prat, E. Ciruelos, Pembrolizumab and Paclitaxel in HR+/HER2- Breast Cancer with HER2-Enriched or Basal-Like Subtypes. Clin Cancer Res, (2026).

44. T. Pascual, M. Vidal, J. M. Cejalvo, E. Vega, E. Sanfeliu, G. Villacampa, S. Ganau, A. M. J. Parreño, E. Zamora, I. Miranda, A. Delgado, B. Bermejo, E. Seguí, F. Brasó-Maristanty, L. de la Cruz-Merino, M. Juan, P. Galván, X. Gonzàlez-Farré, S. Chillara, P. Villagrasa, A. D. Pfefferle, C. E. O’Connell, J. M. Ferrero-Cafiero, M. Oliveira, C. M. Perou, A. Prat, Talimogene laherparepvec and atezolizumab in HER2-negative breast cancer following neoadjuvant chemotherapy: a window-of-opportunity phase II trial (SOLTI-1503 PROMETEO). Nat Commun 17, (2026).

45. S. Pernas, E. Sanfeliu, G. Villacampa, J. Salvador, A. Perelló, X. González, B. Jiménez, M. Merino, P. Palacios, T. Pascual, E. Alba, L. Villanueva, S. Chillara, J. M. Ferrero-Cafiero, P. Galvan, A. Prat, E. Ciruelos, Palbociclib and letrozole for hormone receptor-positive HER2-negative breast cancer with residual disease after neoadjuvant chemotherapy. npj Breast Cancer 10, 101 (2024).

46. E. Sanfeliu, A. Martínez-Romero, M. Marín-Aguilera, S. Cobo, B. González-Farré, E. Hernandez-Illan, P. Jares, J. A. Puig-Butillé, M. Muñoz, R. Gómez-Bravo, M. Tapia, C. Tebar, C. Saura, S. Escrivà-de-Romaní, J. Soberino, J. Cortés, S. Morales, K. Amillano, L. Paré, P. Villagrasa, W. Buckingham, F. Pardo, J. S. Parker, F. Brasó-Maristany, E. Ciruelos, R. Sánchez-Bayona, O. Martinez-Sáez, J. M. Cejalvo, A. Prat, Associations of the HER2DX Genomic Test with Biological and Pathologic Features in HER2-Positive Breast Cancer. Clinical Cancer Research 32, 570–580 (2026).

47. R. Salgado, C. Denkert, S. Demaria, N. Sirtaine, F. Klauschen, G. Pruneri, S. Wienert, G. Van den Eynden, F. L. Baehner, F. Penault-Llorca, E. A. Perez, E. A. Thompson, W. F. Symmans, A. L. Richardson, J. Brock, C. Criscitiello, H. Bailey, M. Ignatiadis, G. Floris, J. Sparano, Z. Kos, T. Nielsen, D. L. Rimm, K. H. Allison, J. S. Reis-Filho, S. Loibl, C. Sotiriou, G. Viale, S. Badve, S. Adams, K. Willard-Gallo, S. Loi, The evaluation of tumor-infiltrating lymphocytes (TILs) in breast cancer: recommendations by an International TILs Working Group 2014. Ann Oncol 26, 259–271 (2015).

48. L. Vanhersecke, A. Bougouin, A. Crombé, M. Brunet, C. Sofeu, M. Parrens, H. Pierron, A. Bonhomme, N. Lembege, C. Rey, V. Velasco, I. Soubeyran, H. Begueret, A. Bessede, A. Bellera, J. Y. Scoazec, A. Italiano, C. S. Fridman, W. H. Fridman, F. Le Loarer, Standardized Pathology Screening of Mature Tertiary Lymphoid Structures in Cancers. Lab Invest 103, 100063 (2023).

49. M. Martín, S. R. Stecklein, O. Gluz, G. Villacampa, M. Monte-Millán, U. Nitz, S. Cobo, M. Christgen, F. Brasó-Maristany, E. L. Álvarez, I. Echavarría, B. Conte, S. Kuemmel, C. Bueno-Muiño, Y. Jerez, R. Kates, M. Cebollero, C. Kolberg-Liedtke, O. Bueno, J. Á. García-Saenz, F. Moreno, E. M. Grischke, H. Forstbauer, M. Braun, M. Warm, J. Hackmann, C. Uleer, B. Aktas, C. Schumacher, R. Wuerstleins, M. Graeser, C. zu Eulenburg, H. H. Kreipe, H. Gómez, T. Massarrah, B. Herrero, L. Paré, U. Bohn, S. López-Tarruella, A. Vivancos, E. Sanfeliu, J. S. Parker, C. M. Perou, P. Villagrasa, A. Prat, P. Sharma, N. Harbeck, TNBC-DX genomic test in early-stage triple-negative breast cancer treated with neoadjuvant taxane-based therapy. Annals of Oncology 36, 158–171 (2025).

50. D. C. Koboldt, R. S. Fulton, M. D. McLellan, H. Schmidt, J. Kalicki-Veizer, J. F. McMichael, L. L. Fulton, D. J. Dooling, L. Ding, E. R. Mardis, R. K. Wilson, A. Ally, M. Balasundaram, Y. S. N. Butterfield, R. Carlsen, C. Carter, A. Chu, E. Chuah, H.-J. E. Chun, R. J. N. Coope, N. Dhalla, R. Guin, C. Hirst, M. Hirst, R. A. Holt, D. Lee, H. I. Li, M. Mayo, R. A. Moore, A. J. Mungall, E. Pleasance, A. Gordon Robertson, J. E. Schein, A. Shafiei, P. Sipahimalani, J. R. Slobodan, D. Stoll, A. Tam, N. Thiessen, R. J. Varhol, N. Wye, T. Zeng, Y. Zhao, I. Birol, S. J. M. Jones, M. A. Marra, A. D. Cherniack, G. Saksena, R. C. Onofrio, N. H. Pho, S. L. Carter, S. E. Schumacher, B. Tabak, B. Hernandez, J. Gentry, H. Nguyen, A. Crenshaw, K. Ardlie, R. Beroukhim, W. Winckler, G. Getz, S. B. Gabriel, M. Meyerson, L. Chin, P. J. Park, R. Kucherlapati, K. A. Hoadley, J. Todd Auman, C. Fan, Y. J. Turman, Y. Shi, L. Li, M. D. Topal, X. He, H.-H. Chao, A. Prat, G. O. Silva, M. D. Iglesia, W. Zhao, J. Usary, J. S. Berg, M. Adams, J. Booker, J. Wu, A. Gulabani, T. Bodenheimer, A. P. Hoyle, J. V. Simons, M. G. Soloway, L. E. Mose, S. R. Jefferys, S. Balu, J. S. Parker, D. Neil Hayes, C. M. Perou, S. Malik, S. Mahurkar, H. Shen, D. J. Weisenberger, T. Triche Jr, P. H. Lai, M. S. Bootwalla, D. T. Maglinte, B. P. Berman, D. J. Van Den Berg, S. B. Baylin, P. W. Laird, C. J. Creighton, L. A. Donehower, G. Getz, M. Noble, D. Voet, G. Saksena, N. Gehlenborg, D. DiCara, J. Zhang, H. Zhang, C.-J. Wu, S. Yingchun Liu, M. S. Lawrence, L. Zou, A. Sivachenko, P. Lin, P. Stojanov, R. Jing, J. Cho, R. Sinha, R. W. Park, M.-D. Nazaire, J. Robinson, H. Thorvaldsdottir, J. Mesirov, P. J. Park, L. Chin, S. Reynolds, R. B. Kreisberg, B. Bernard, R. Bressler, T. Erkkila, J. Lin, V. Thorsson, W. Zhang, I. Shmulevich, G. Ciriello, N. Weinhold, N. Schultz, J. Gao, E. Cerami, B. Gross, A. Jacobsen, R. Sinha, B. Arman Aksoy, Y. Antipin, B. Reva, R. Shen, B. S. Taylor, M. Ladanyi, C. Sander, P. Anur, P. T. Spellman, Y. Lu, W. Liu, R. R. G. Verhaak, G. B. Mills, R. Akbani, N. Zhang, B. M. Broom, T. D. Casasent, C. Wakefield, A. K. Unruh, K. Baggerly, K. Coombes, J. N. Weinstein, D. Haussler, C. C. Benz, J. M. Stuart, S. C. Benz, J. Zhu, C. C. Szeto, G. K. Scott, C. Yau, E. O. Paull, D. Carlin, C. Wong, A. Sokolov, J. Thusberg, S. Mooney, S. Ng, T. C. Goldstein, K. Ellrott, M. Grifford, C. Wilks, S. Ma, B. Craft, C. Yan, Y. Hu, D. Meerzaman, J. M. Gastier-Foster, J. Bowen, N. C. Ramirez, A. D. Black, R. E. Pyatt, P. White, E. J. Zmuda, J. Frick, T. M. Lichtenberg, R. Brookens, M. M. George, M. A. Gerken, H. A. Harper, K. M. Leraas, L. J. Wise, T. R. Tabler, C. McAllister, T. Barr, M. Hart-Kothari, K. Tarvin, C. Saller, G. Sandusky, C. Mitchell, M. V. Iacocca, J. Brown, B. Rabeno, C. Czerwinski, N. Petrelli, O. Dolzhansky, M. Abramov, O. Voronina, O. Potapova, J. R. Marks, W. M. Suchorska, D. Murawa, W. Kycler, M. Ibbs, K. Korski, A. Spychała, P. Murawa, J. J. Brzeziński, H. Perz, R. Łaźniak, M. Teresiak, H. Tatka, E. Leporowska, M. Bogusz-Czerniewicz, J. Malicki, A. Mackiewicz, M. Wiznerowicz, X. Van Le, B. Kohl, N. Viet Tien, R. Thorp, N. Van Bang, H. Sussman, B. Duc Phu, R. Hajek, N. Phi Hung, T. Viet The Phuong, H. Quyet Thang, K. Zaki Khan, R. Penny, D. Mallery, E. Curley, C. Shelton, P. Yena, J. N. Ingle, F. J. Couch, W. L. Lingle, T. A. King, A. Maria Gonzalez-Angulo, G. B. Mills, M. D. Dyer, S. Liu, X. Meng, M. Patangan, N. The Cancer Genome Atlas, L. Genome sequencing centres: Washington University in St, B. C. C. A. Genome characterization centres, I. Broad, Brigham, H. Women’s, S. Harvard Medical, C. H. University of North Carolina, H. University of Southern California/Johns, M. Genome data analysis: Baylor College of, B. Institute for Systems, C. Memorial Sloan-Kettering Cancer, H. Oregon, U. Science, M. D. A. C. C. The University of Texas, S. C. B. I. University of California, Nci, R. Biospecimen core resource: Nationwide Children’s Hospital Biospecimen Core, A.-I. Tissue source sites, Christiana, Cureline, C. Duke University Medical, C. The Greater Poland Cancer, Ilsbio, C. International Genomics, C. Mayo, Mskcc, M. D. A. C. Center, Comprehensive molecular portraits of human breast tumours. Nature 490, 61–70 (2012).

51. C. Curtis, S. P. Shah, S.-F. Chin, G. Turashvili, O. M. Rueda, M. J. Dunning, D. Speed, A. G. Lynch, S. Samarajiwa, Y. Yuan, S. Gräf, G. Ha, G. Haffari, A. Bashashati, R. Russell, S. McKinney, C. Caldas, S. Aparicio, C. Curtis†, S. P. Shah, C. Caldas, S. Aparicio, J. D. Brenton, I. Ellis, D. Huntsman, S. Pinder, A. Purushotham, L. Murphy, C. Caldas, S. Aparicio, C. Caldas, H. Bardwell, S.-F. Chin, C. Curtis, Z. Ding, S. Gräf, L. Jones, B. Liu, A. G. Lynch, I. Papatheodorou, S. J. Sammut, G. Wishart, S. Aparicio, S. Chia, K. Gelmon, D. Huntsman, S. McKinney, C. Speers, G. Turashvili, P. Watson, I. Ellis, R. Blamey, A. Green, D. Macmillan, E. Rakha, A. Purushotham, C. Gillett, A. Grigoriadis, S. Pinder, E. de Rinaldis, A. Tutt, L. Murphy, M. Parisien, S. Troup, C. Caldas, S.-F. Chin, D. Chan, C. Fielding, A.-T. Maia, S. McGuire, M. Osborne, S. M. Sayalero, I. Spiteri, J. Hadfield, S. Aparicio, G. Turashvili, L. Bell, K. Chow, N. Gale, D. Huntsman, M. Kovalik, Y. Ng, L. Prentice, C. Caldas, S. Tavaré, C. Curtis, M. J. Dunning, S. Gräf, A. G. Lynch, O. M. Rueda, R. Russell, S. Samarajiwa, D. Speed, F. Markowetz, Y. Yuan, J. D. Brenton, S. Aparicio, S. P. Shah, A. Bashashati, G. Ha, G. Haffari, S. McKinney, A. Langerød, A. Green, E. Provenzano, G. Wishart, S. Pinder, P. Watson, F. Markowetz, L. Murphy, I. Ellis, A. Purushotham, A.-L. Børresen-Dale, J. D. Brenton, S. Tavaré, C. Caldas, S. Aparicio, M. Group, c. Co, c. Writing, c. Steering, Tissue, s. clinical data source, U. K. C. R. I. University of Cambridge/Cancer Research, A. British Columbia Cancer, N. University of, L. King’s College, B. Manitoba Institute of Cell, c. Cancer genome/transcriptome characterization, s. Data analysis, The genomic and transcriptomic architecture of 2,000 breast tumours reveals novel subgroups. Nature 486, 346–352 (2012).

52. L. H. Saal, J. Vallon-Christersson, J. Häkkinen, C. Hegardt, D. Grabau, C. Winter, C. Brueffer, M.-H. E. Tang, C. Reuterswärd, R. Schulz, A. Karlsson, A. Ehinger, J. Malina, J. Manjer, M. Malmberg, C. Larsson, L. Rydén, N. Loman, Å. Borg, The Sweden Cancerome Analysis Network - Breast (SCAN-B) Initiative: a large-scale multicenter infrastructure towards implementation of breast cancer genomic analyses in the clinical routine. Genome Medicine 7, 20 (2015).

53. C. Brueffer, S. Gladchuk, C. Winter, J. Vallon-Christersson, C. Hegardt, J. Häkkinen, A. M. George, Y. Chen, A. Ehinger, C. Larsson, N. Loman, M. Malmberg, L. Rydén, Å. Borg, L. H. Saal, The mutational landscape of the SCAN-B real-world primary breast cancer transcriptome. EMBO Molecular Medicine 12, EMMM202012118 (2020).

54. C. Gomà, M. Mollà, G. Oses, E. Sanfeliu, A. Rodríguez-Hernández, B. González-Farré, P. Galván, V. Sirenko, O. Castillo, P. Blasco, A. Llop-Guevara, V. Serra, S. Ganau, B. Úbeda, X. Bargalló, C. Cases, A. Latorre-Musoll, O. Martínez-Sáez, I. Garcia-Fructuoso, R. Gómez-Bravo, F. Schettini, B. Adamo, T. Pascual, M. Vidal, M. Muñoz, M. Laplana, I. Cebrecos, E. Mension, A. Prat, F. Brasó-Maristany, Preoperative Radiation Therapy-Induced Molecular and Immune Modulation in Early-Stage Breast Cancer: Results From the YOUNGSTER trial. Int J Radiat Oncol Biol Phys 124, 122–132 (2026).

55. M. Vidal, C. Falato, T. Pascual, R. Sanchez-Bayona, M. Muñoz-Mateu, I. Cebrecos, X. Gonzalez-Farré, T. Cortadellas, M. Margelí Vila, M. A. Luna, C. Siso, K. Amillano, P. Galván, M. A. Bergamino, J. M. Ferrero-Cafiero, F. Salvador, A. Espinosa Guerrero, L. Pare, E. Sanfeliu, A. Prat, M. Bellet, Elacestrant in Women with Estrogen Receptor-Positive and HER2-Negative Early Breast Cancer: Results from the Preoperative Window-of-Opportunity ELIPSE Trial. Clin Cancer Res 31, 1223–1232 (2025).

56. A. Prat, C. Saura, T. Pascual, C. Hernando, M. Muñoz, L. Paré, B. González Farré, P. L. Fernández, P. Galván, N. Chic, X. González Farré, M. Oliveira, M. Gil-Gil, M. Arumi, N. Ferrer, A. Montaño, Y. Izarzugaza, A. Llombart-Cussac, R. Bratos, S. González Santiago, E. Martínez, S. Hoyos, B. Rojas, J. A. Virizuela, V. Ortega, R. López, P. Céliz, E. Ciruelos, P. Villagrasa, J. Gavilá, Ribociclib plus letrozole versus chemotherapy for postmenopausal women with hormone receptor-positive, HER2-negative, luminal B breast cancer (CORALLEEN): an open-label, multicentre, randomised, phase 2 trial. Lancet Oncol 21, 33–43 (2020).

57. R. Gómez-Bravo, B. Walbaum, M. Bergamino, O. Martínez-Sáez, F. Schettini, E. Seguí, I. García-Fructuoso, T. Pascual, N. Chic, M. González, A. Rodríguez, M. Rey, P. Giménez-Xavier, P. Blasco, O. Castillo, P. Galván, E. Sanfeliu, B. González-Farré, M. Vidal, B. Adamo, F. Brasó-Maristany, A. Prat, M. Muñoz, Ki67 dynamic predicts endocrine sensitivity in estrogen receptor-positive/HER2-negative breast cancer patients undergoing preoperative endocrine therapy. ESMO Open 10, 105845 (2025).

58. E. S. Solis, A. Martínez-Romero, F. Brasó-Maristany, T. Pascual, M. Marin, R. G. Bravo, I. G. Fructuoso, A. R. Hernandez, O. M. Saez, P. Galván, C. S. Manich, E. M. Ciruelos, A. Waks, S. M. Tolaney, M. Oliveira, L. P. Brunet, J. M. Ferrero-Cafiero, A. Prat, E. S. Torres, 13P A novel H&amp;E-based classification of tertiary lymphoid structures in breast cancer. ESMO Open 10, (2025).

