## Supplementary Figures for "Immune organization defines adaptive immune competence and clinical outcome in breast cancer"

### Supplementary Figure S1. Representative histologic and immunohistochemical features across TLS categories.

Representative examples of tumors without TLS and low or high lymphocyte infiltration, as well as tumors with TLS lacking or containing germinal centers (GCs). Hematoxylin and eosin (H&E) staining and immunohistochemistry (IHC) for Ki67, CD20, and CD23 are shown. Within TLS, CD20 highlights B-cell aggregates, CD23 identifies follicular dendritic cell networks characteristic of GC-containing TLS, and Ki67 highlights the proliferative activity within GCs.

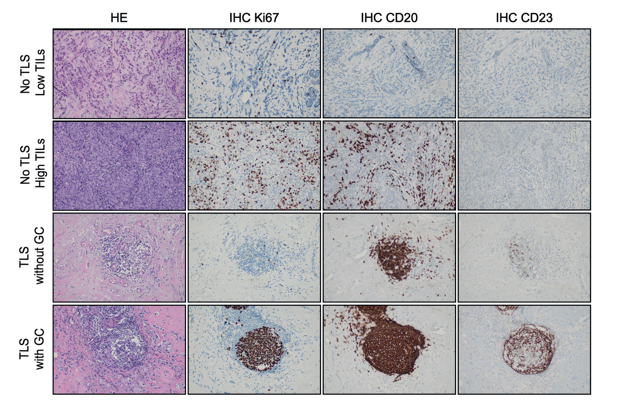

**Abbreviations:** GC, germinal center; H&E, hematoxylin and eosin; IHC, immunohistochemistry; TLS, tertiary lymphoid structure.

### Supplementary Figure S2. Distribution of TLS categories assessed by two independent pathologists, showing high concordance.

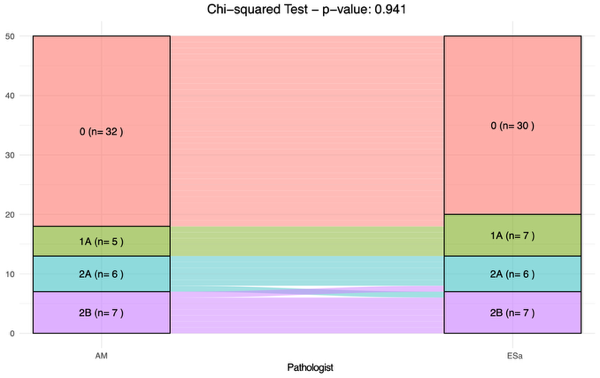

Supplementary Figure S3. Distribution of histologic TLS categories according to disease setting and breast cancer subtype**.**

**(A)** Distribution of TLS categories (0, 1a, 1b, 2a, 2b) according to tumor site (primary versus metastatic), showing a higher proportion of TLS-negative tumors and reduced prevalence of mature TLS in metastatic samples. P value was calculated using a chi-square test. **(B)** Distribution of TLS categories across breast cancer subtypes (HR+/HER2-, HER2+, and TNBC), illustrating differences in TLS prevalence and maturation. Unadjusted P values are shown (chi-square test); significance was not retained after correction for multiple testing. **(C)** TLS distribution stratified by breast cancer subtype (HR+/HER2-, HER2+, and TNBC), shown separately for primary and metastatic tumors.

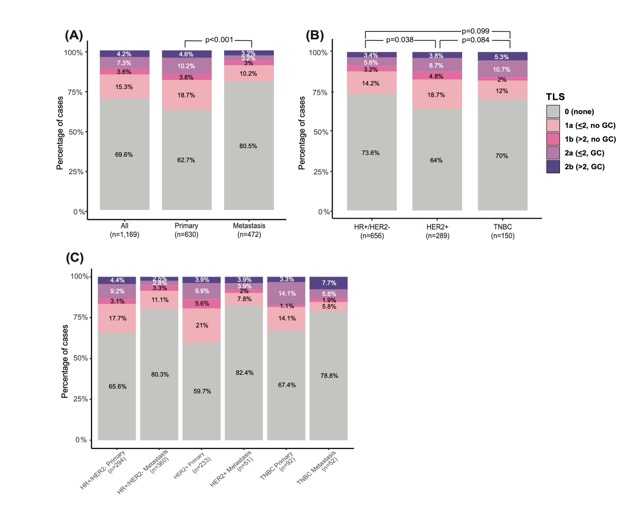

**Abbreviations**: TLS, tertiary lymphoid structures; HR, hormone receptor; HER2, human epidermal growth factor receptor 2; TNBC, triple-negative breast cancer.

### Supplemental Figure S4. TLS category distribution by metastatic site.

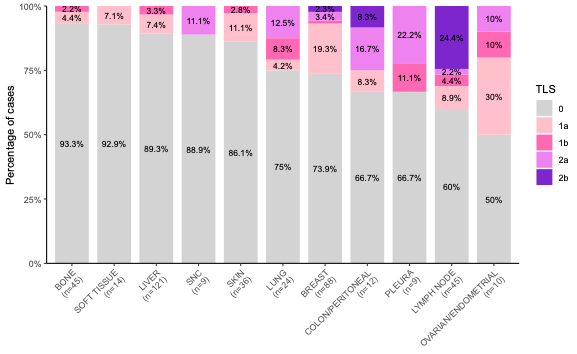

**Abbreviations:** TLS, tertiary lymphoid structures; CNS, central nervous system.

Supplementary Figure S5. Cross-prediction performance of TLS-GEP and TIL-GEP in the TCGA cohort (n=1,035).

Left panel: TLS-GEP scores stratified by histologic TIL levels (<30% vs ≥30%), showing higher scores in TIL-high tumors. Right panel: TIL-GEP scores stratified by histologic TLS status (mature TLS present vs absent), showing higher scores in tumors with mature TLS. In both cases, cross-prediction performance (AUC ≈0.72) was lower than that observed for each signature with its corresponding histologic feature, indicating partial overlap but non-equivalence between immune infiltration and organization.

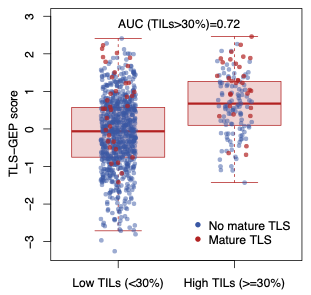

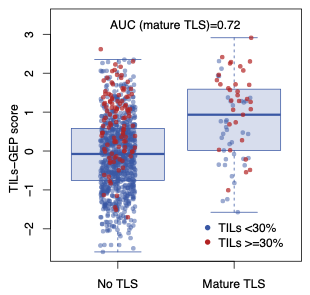

**Abbreviations:** TLS, tertiary lymphoid structures; TILs, tumor-infiltrating lymphocytes; GEP, gene expression profile; AUC, area under the curve.

Supplementary Figure S6. Correlation between TIL-GEP and TLS-GEP scores in the METABRIC (n=1,985; left) and SCAN-B (n=6,655; right) cohorts, assessed using Spearman correlation.

Spearman correlation coefficients (ρ) and corresponding p-values are shown.

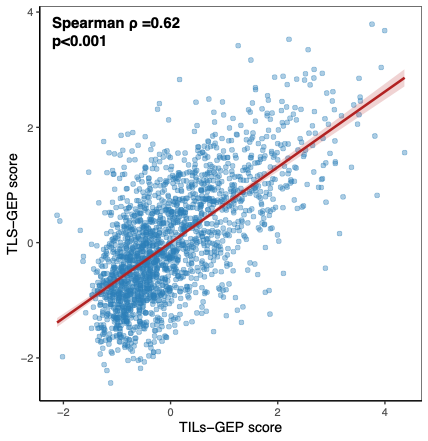

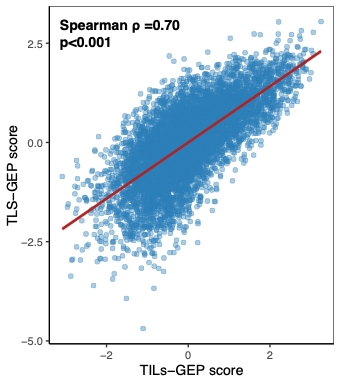

**Abbreviations:** TILs, tumor-infiltrating lymphocytes; TLS, tertiary lymphoid structures; GEP, gene expression profile; ρ, Spearman correlation coefficient.

Supplementary Figure S7. Kaplan–Meier curves for overall survival in the TCGA (n=1,035; left) and METABRIC (n=1,985; right) cohorts, stratified by combined TLS-GEP and TIL-GEP groups using median-based dichotomization.

Tumors were classified into four groups according to high or low TLS-GEP and TIL-GEP scores. Statistical significance was assessed using the log-rank test.

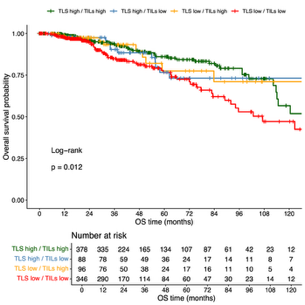

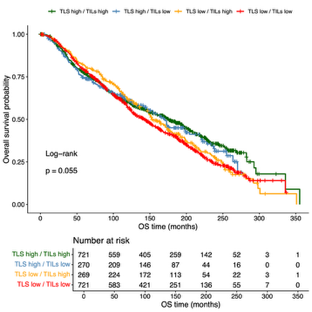

Supplementary Figure S8. Prognostic impact of immune organization across intrinsic molecular risk groups in the SCAN-B HR+HER2- breast cancer cohort (n=5,036).

(A) Kaplan–Meier analyses of overall survival according to high versus low TLS-GEP within PAM50 ROR-low, ROR-intermediate, and ROR-high tumors. (B) Kaplan–Meier analysis of overall survival according to combined intrinsic molecular risk (ROR) and immune organization states defined by TLS-GEP.

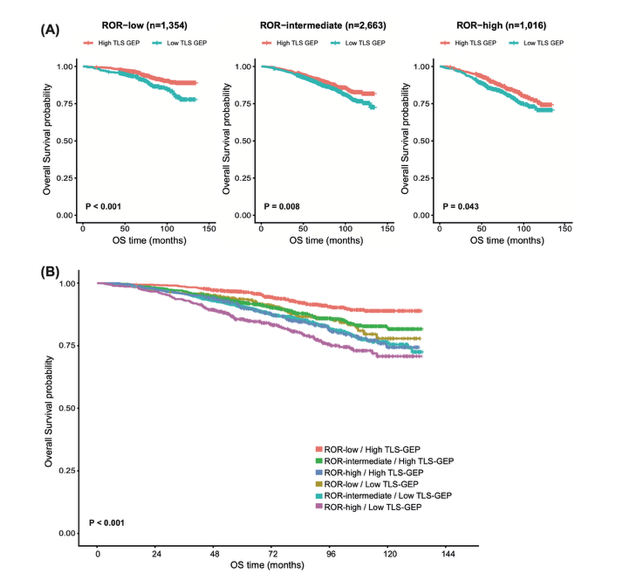

Supplementary Figure S9. Baseline TCR/BCR repertoire characteristics according to immune organization.

Tumors with higher IGG expression show increased TCR clonotype counts (**A**) and higher clonal diversity (**B**), consistent with a more abundant and polyclonal repertoire. BCR diversity is also higher in tumors with immune organization, both by TLS (**C**) and IGG signature (**D**). TLS-negative tumors display a more oligoclonal baseline T-cell repertoire, with more clones accounting for >1% of the repertoire (**E**) and a higher fraction accounted for by the top five (**F**). Forest plot of odds ratios (OR) and 95% confidence intervals from logistic regression models: TCR diversity (Shannon entropy) remained significantly associated with TLS in multivariate analysis, whereas TIL percentage did not (**G**). TIL abundance and diversity were more strongly correlated with IGG expression (**H, I**) than TIL percentage (**J, K**).

**C**

**B**

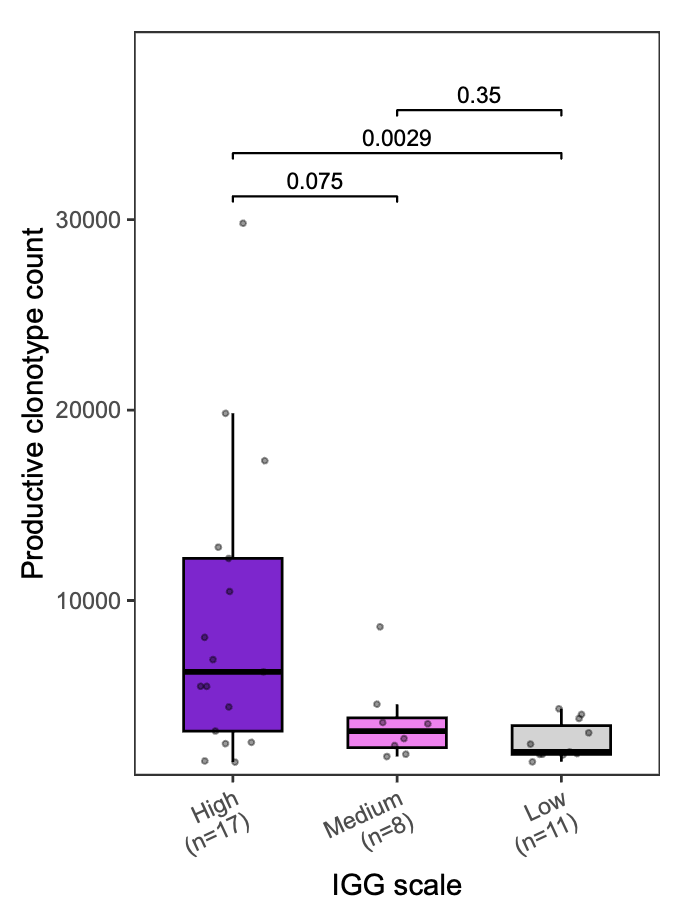

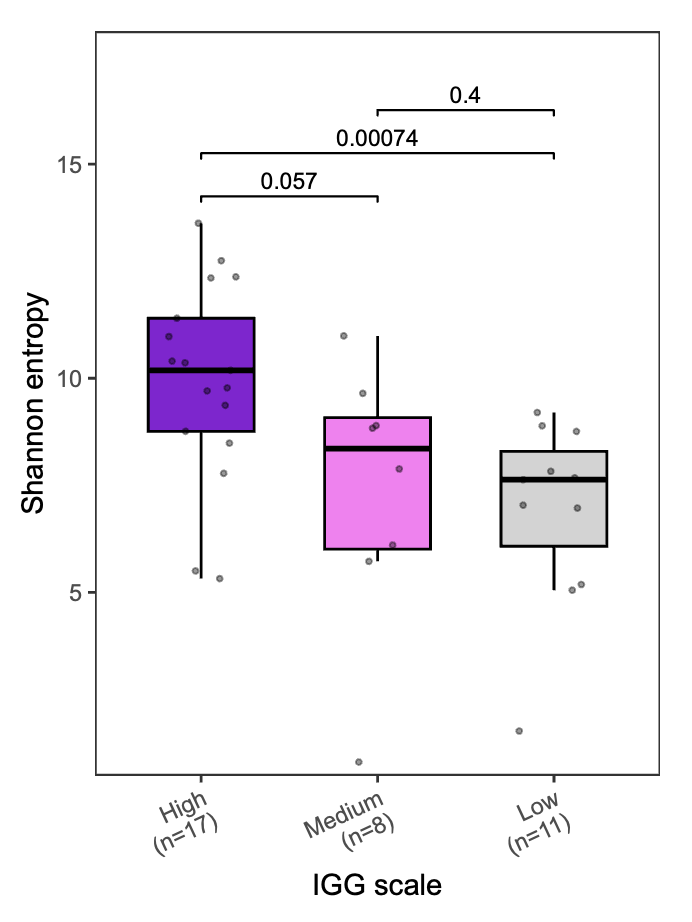

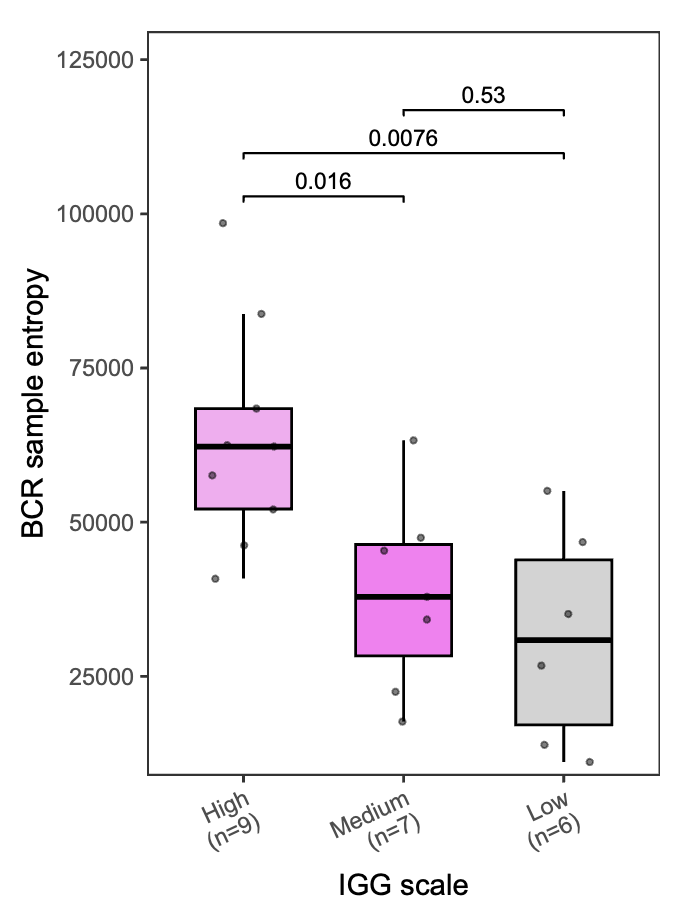

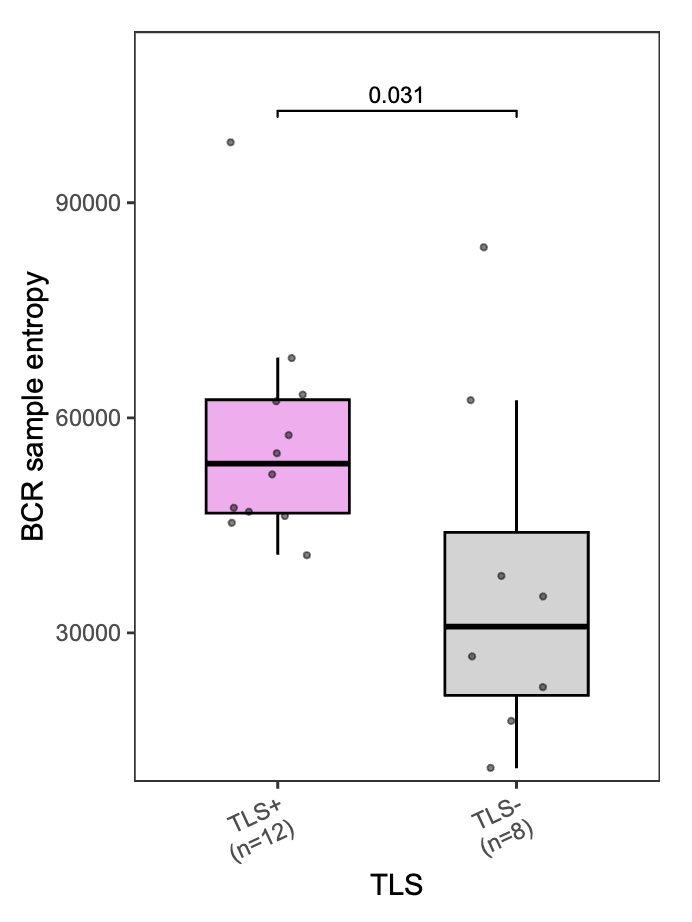

**D**

**A**

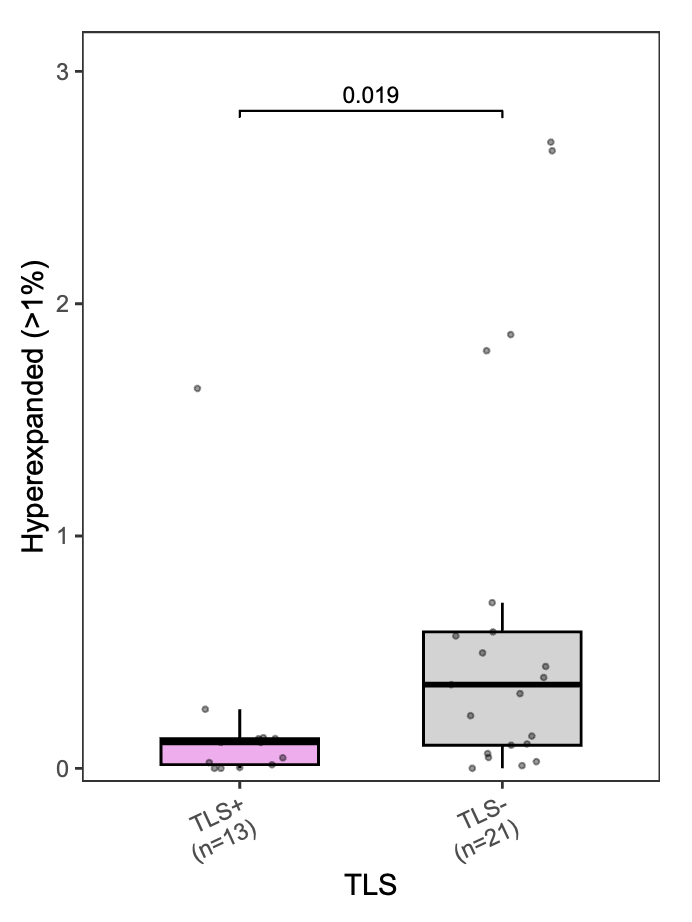

**ED**

**F**

**G**

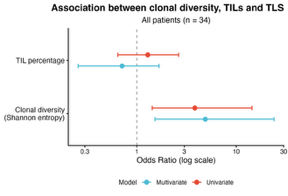

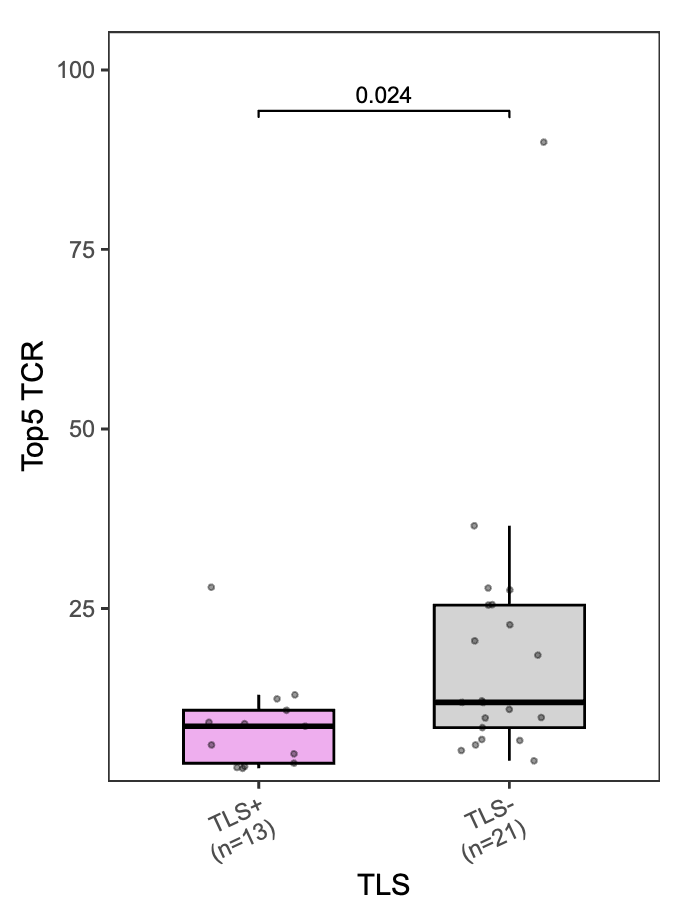

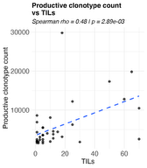

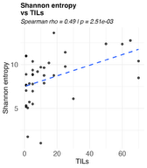

**K**

**J**

**I**

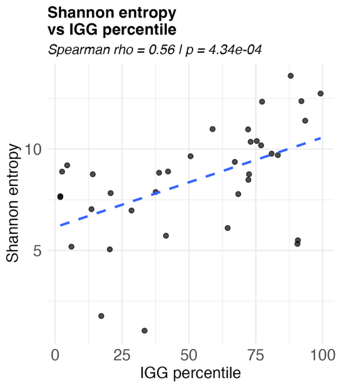

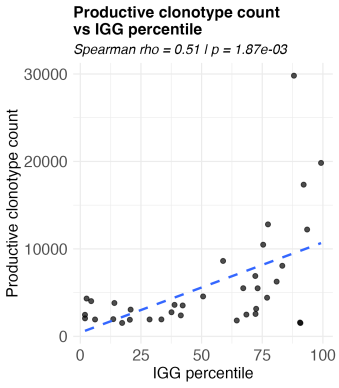

**H**

Supplementary Figure S10. Association between TCR dynamics and clinical response.

Complete responders exhibited more polyclonal baseline T-cell repertoires (**A**), followed by a decline in TCR diversity driven by the selective expansion of tumor-reactive clones (**B**). Consistent with IGG-high and TLS-positive tumors, pCR cases showed a substantial expansion of pre-existing tumor-derived clonotypes, in contrast to poor responders (**C**). The absolute number of clones that at least doubled their clonal fraction was significantly higher in the pCR group (**D**). Newly gained –non-tumor-derived– TCRs were more frequent in non-pCR (**E**), IGG-low (**F**), and TLS-negative tumors (**G**).

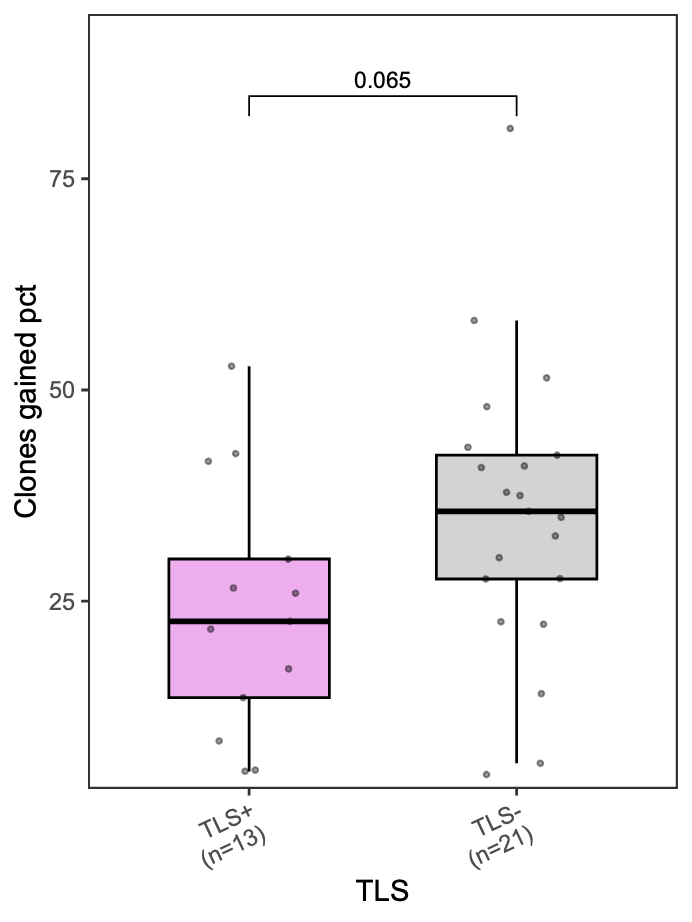

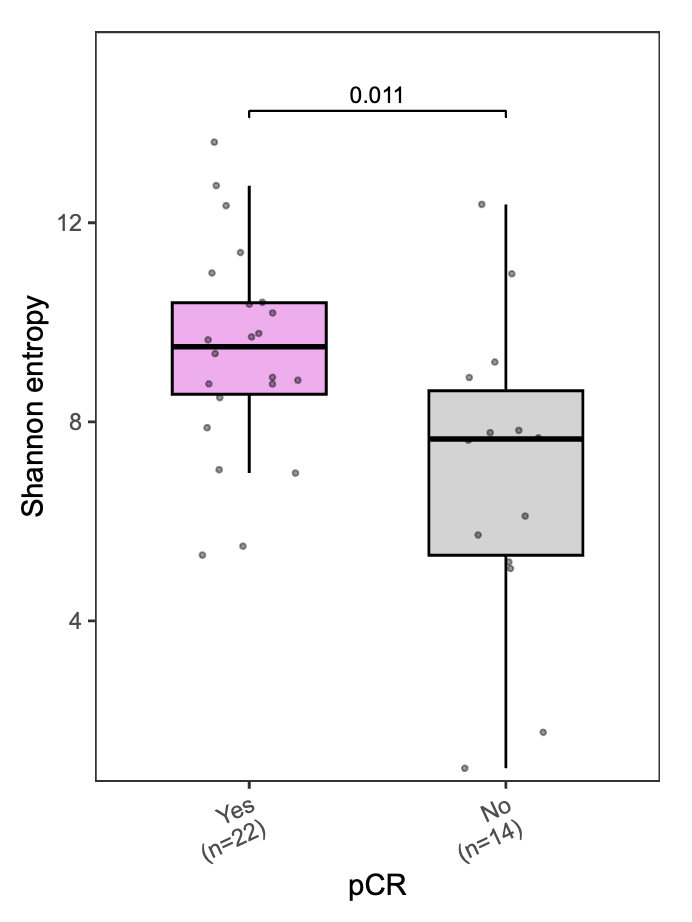

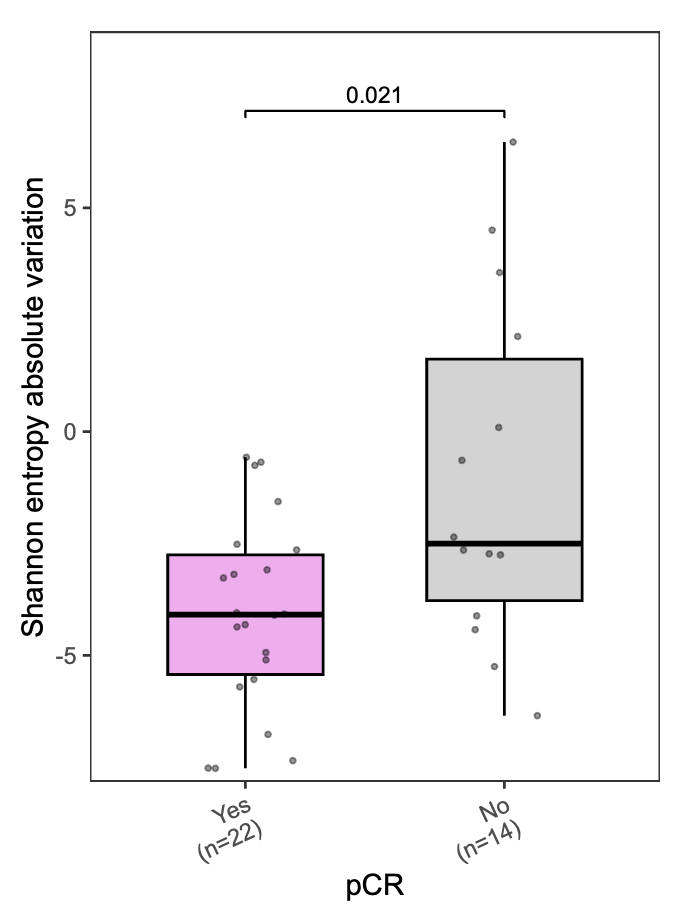

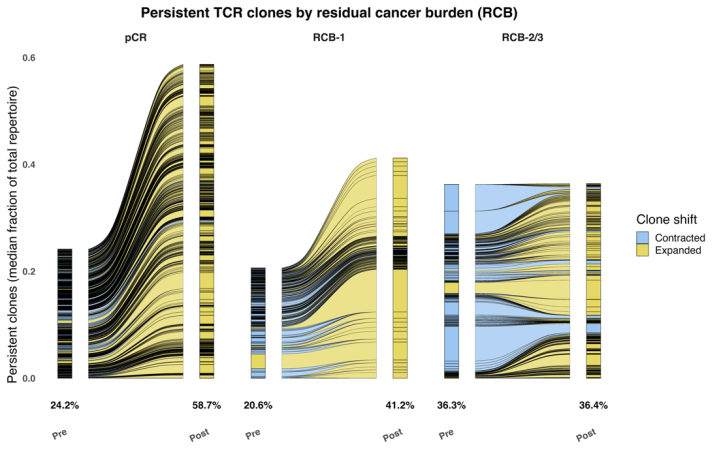

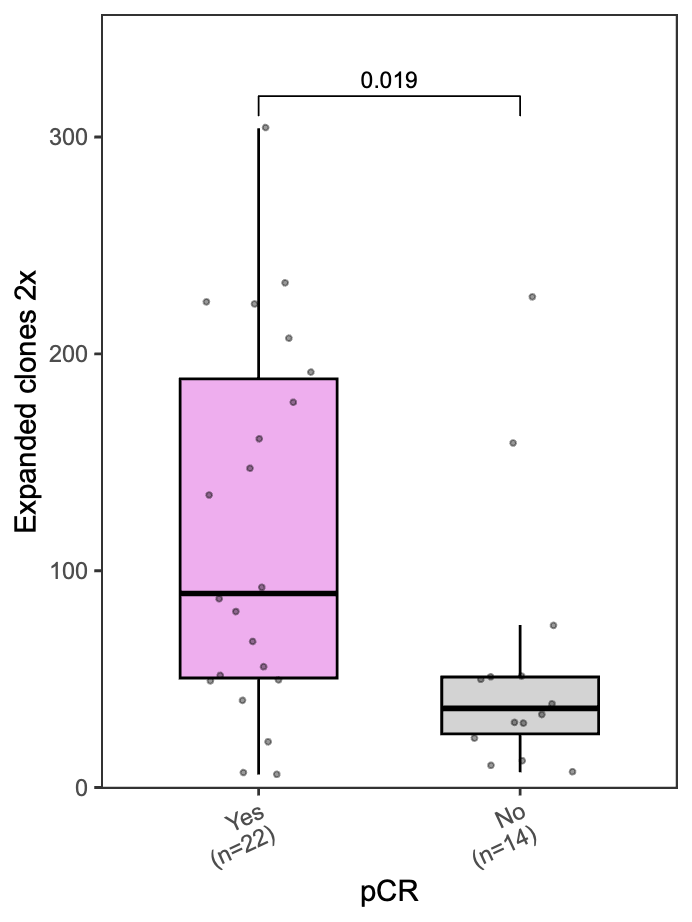

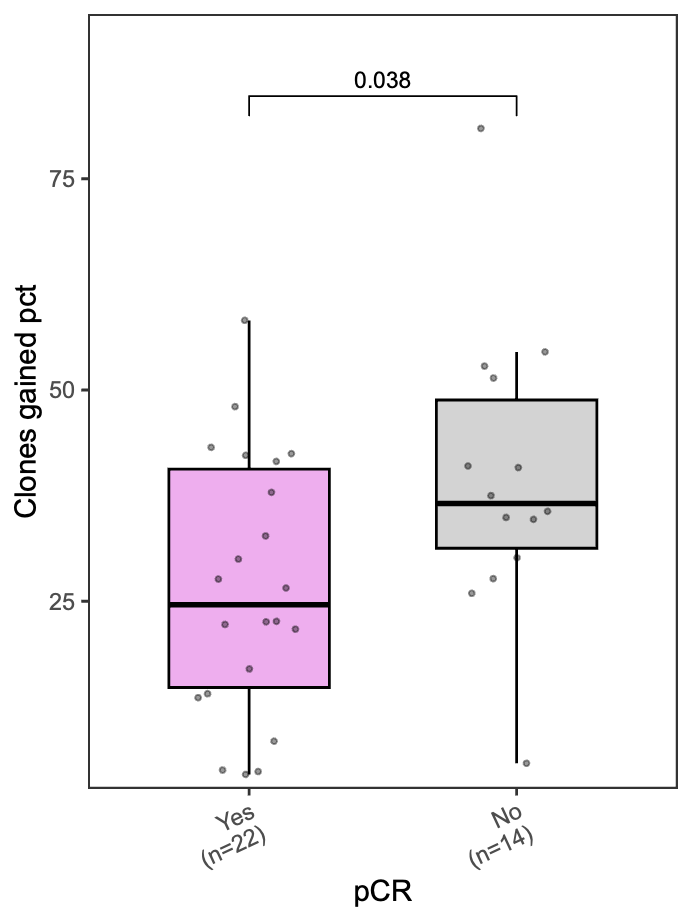

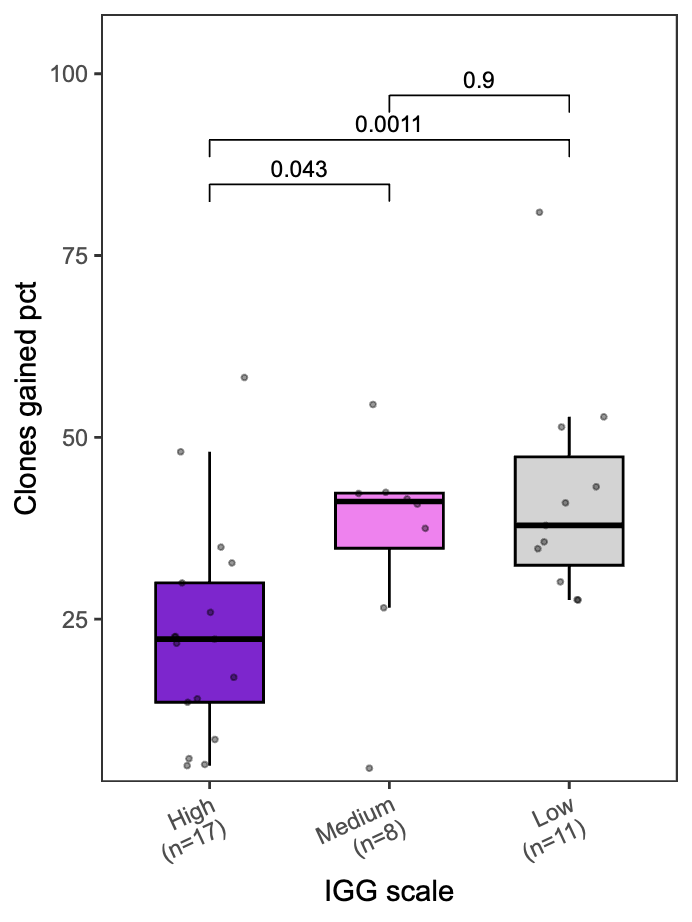

**DE**

**E**

**F**

**G**

**A**

**B**

**C**

### ****Supplementary Fig. S11. Changes in stromal TIL categories following treatment across paired breast cancer cohorts.****

Changes in stromal TIL categories (<10%, 10–30%, and >30%) across paired tumor samples from PROMETEO-I (T-VEC plus atezolizumab), PROMETEO-II (CDK4/6 inhibitor plus endocrine therapy), and preoperative endocrine therapy cohorts. Alluvial plots show transitions between TIL categories from baseline to on-treatment and/or surgery samples.

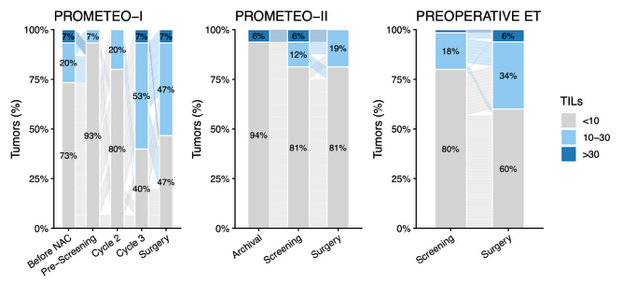

### ****Supplementary Fig. S12. Correlation between treatment-induced changes in TLS-GEP and HER2DX IGG signature.****

Correlation between treatment-induced changes (Δ) in TLS-GEP and the HER2DX IGG signature across paired tumor samples from all studies included in the pooled analysis. Each dot represents one patient and colors indicate study cohort. The solid line represents the linear regression fit with the shaded area indicating the 95% confidence interval. Correlation was assessed using Spearman's rank correlation coefficient.

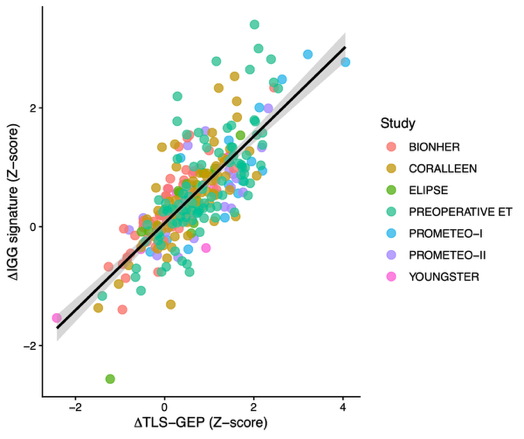
