## Supplementary Tables for "Immune organization defines adaptive immune competence and clinical outcome in breast cancer"

| | |
|---|---|
| **1** | TLS classification association with lymphocyte abundance and humoral immune activation overall and by breast cancer subtype. |
| **2** | Differentially expressed genes between TIL-high (≥30%) and TIL-low (<30%) tumors identified by SAM unpaired analysis. |
| **3** | Differentially expressed genes between tumors with mature TLS (TLS 2a/2b) and tumors with no mature TLS (TLS 0/1a/1b) identified by SAM unpaired analysis. |
| **4** | Factors associated with the presence of TLS (TLS≠0 vs 0) and mature TLS (TLS 2a/2b) |
| **5** | Multivariable logistic regression models evaluating factors associated with mature TLS (TLS 2a/2b). |
| **6** | Gene composition and overlap of the TLS-GEP and TILs-GEP signatures. |
| **7** | Multivariable Cox regression analyses of immune organization signatures adjusted for PAM50 ROR-P across the TCGA, METABRIC, and SCAN-B |
| **8** | Main clinicopathological characteristics of patients with available TCR-seq data stratified by tertiary lymphoid structure (TLS) status. |
| **9** | Characteristics of the spatial transcriptomics cohort (n = 8 samples) |
| **10** | Predefined gene modules and marker genes used to calculate mean expression per region of interest (ROI). |
| **11** | Treatment-induced changes in TLS-GEP and TIL-GEP across studies. |
| **12** | Association between treatment-induced changes in TLS-GEP and TIL-GEP and treatment response. |

**Supplementary Table S1. Association of TLS categories with lymphocyte abundance (TILs) and HER2DX IgG status overall and by breast cancer subtype.**

| Overall population | | | |
|---|---|---|---|
| **TLS category** | **Cases, n (%)** | **Median TILs, % (IQR)** | **HER2DX IGG-high, %** |
| TLS 0 | 814 (69.6%) | 3% (1-8) | 20.9% |
| TLS 1a (≤2, no GC) | 179 (15.3%) | 8% (4-18) | 43.6% |
| TLS 1b (>2, no GC) | 42 (3.6%) | 12% (7-25) | 50.0% |
| TLS 2a (≤2, GC) | 85 (7.3%) | 12% (5-25) | 55.3% |
| TLS 2b (>2, GC) | 49 (4.2%) | 30% (15-43) | 77.6% |
| **HR+/HER2-** | | | |
| **TLS category** | **Cases, n (%)** | **Median TILs, % (IQR)** | **HER2DX IGG-high, %** |
| TLS 0 | 483 (73.6%) | 2% (1-5) | 16.1% |
| TLS 1a (≤2, no GC) | 93 (14.2%) | 5% (4-12) | 39.8% |
| TLS 1b (>2, no GC) | 21 (3.2%) | 11% (5.25-15) | 28.6% |
| TLS 2a (≤2, GC) | 37 (5.6%) | 5% (4-20) | 48.6% |
| TLS 2b (>2, GC) | 22 (3.4%) | 18% (12-40) | 72.3% |
| **HER2+** | | | |
| **TLS category** | **Cases, n (%)** | **Median TILs, % (IQR)** | **HER2DX IGG-high, %** |
| TLS 0 | 185 (64.0%) | 5% (2-12) | 27.6% |
| TLS 1a (≤2, no GC) | 54 (18.7%) | 12% (5-25) | 48.1% |
| TLS 1b (>2, no GC) | 14 (4.8%) | 13.5% (5.75-25) | 64.3% |
| TLS 2a (≤2, GC) | 25 (8.7%) | 18% (7-40) | 48.0% |
| TLS 2b (>2, GC) | 11 (3.8%) | 40% (15-50) | 90.9% |
| **TNBC** | | | |
| **TLS category** | **Cases, n (%)** | **Median TILs, % (IQR)** | **HER2DX IGG-high, %** |
| TLS 0 | 105 (70.0%) | 3% (1-10) | 26.7% |
| TLS 1a (≤2, no GC) | 18 (12.0%) | 15% (5-25) | 44.4% |
| TLS 1b (>2, no GC) | 3 (2.0%) | 35% (25-37.5) | 100.0% |
| TLS 2a (≤2, GC) | 16 (10.7%) | 7% (5-20) | 68.8% |
| TLS 2b (>2, GC) | 8 (5.3%) | 35% (20-40) | 75.0% |

**Supplementary Table S2. Differentially expressed genes between TIL-high (≥30%) and TIL-low (<30%) tumors identified by SAM analysis.**

| Gene ID | Score(d) | Fold Change | FDR (q-value) |
|---|---|---|---|
| *CD79A* | 9.456478034 | 8.587330866 | 0 |
| *IGLV3.25* | 8.661791408 | 8.773287529 | 0 |
| *CD19* | 8.541950882 | 4.740985614 | 0 |
| *CD7* | 8.462197539 | 4.126664872 | 0 |
| *CXCL9* | 8.330307727 | 7.015408765 | 0 |
| *POU2AF1* | 8.295167176 | 5.458531599 | 0 |
| *CD27* | 8.069461847 | 3.608216827 | 0 |
| *CD8A* | 7.958807706 | 3.997698705 | 0 |
| *CTLA4* | 7.940848092 | 3.992718008 | 0 |
| *IGL* | 7.935496711 | 8.301325945 | 0 |
| *CXCL13* | 7.863071289 | 4.931027536 | 0 |
| *IL2RG* | 7.832055335 | 3.827084951 | 0 |
| *ITK* | 7.804599317 | 3.922923139 | 0 |
| *PIM2* | 7.768129791 | 3.46321112 | 0 |
| *SLAMF1* | 7.707402173 | 3.815236322 | 0 |
| *IRF4* | 7.620612342 | 4.743192069 | 0 |
| *LAX1* | 7.595672059 | 3.813818511 | 0 |
| *CD2* | 7.576977659 | 3.550858575 | 0 |
| *CD3G* | 7.546775045 | 3.361877766 | 0 |
| *LY9* | 7.483318007 | 3.747646322 | 0 |
| *PDCD1* | 7.425919092 | 3.424809359 | 0 |
| *SH2D1A* | 7.369360472 | 3.113762392 | 0 |
| *TNFRSF17* | 7.335685037 | 4.409933972 | 0 |
| *GZMA* | 7.047458145 | 3.470698386 | 0 |
| *CD3D* | 7.032539187 | 5.025522048 | 0 |
| *EOMES* | 7.016679234 | 3.887652968 | 0 |
| *IGKC* | 6.796884058 | 6.81659767 | 0 |
| *GNLY* | 6.507525554 | 3.624807398 | 0 |
| *GZMB* | 6.455488225 | 4.00380036 | 0 |
| *IRF8* | 6.096500595 | 2.403945827 | 0 |
| *CD274* | 6.031050263 | 2.412554393 | 0 |
| *STAT4* | 5.992560596 | 2.445896828 | 0 |
| *CXCR6* | 5.972695918 | 3.042555107 | 0 |
| *STAT1* | 5.948706998 | 2.286095776 | 0 |
| *IGJ* | 5.617123299 | 4.289735889 | 0 |
| *IRF1* | 5.383174761 | 2.305583602 | 0 |
| *CD40* | 5.307931292 | 2.048375872 | 0 |
| *ISG20* | 5.225101002 | 2.299381557 | 0 |
| *KLRB1* | 4.790170281 | 2.285035325 | 0 |
| *CD4* | 4.748675693 | 1.883563908 | 0 |
| *EAF2* | 4.386930172 | 1.952994898 | 0 |
| *KYNU* | 4.156655239 | 2.049569609 | 0 |
| *KLRD1* | 3.889043267 | 1.970517755 | 0 |
| *HLA.C* | 3.828153163 | 1.899560763 | 0 |
| *C2orf54* | 3.673029633 | 2.581217529 | 0 |
| *LGALS9* | 3.614873565 | 1.663763644 | 0 |
| *CD84* | 3.608018193 | 1.668722093 | 0 |
| *CD86* | 3.60762905 | 1.609705177 | 0 |
| *MUCL1* | 3.585513053 | 3.622387536 | 0 |

| | | | |
|---|---|---|---|
| *S100A9* | 3.57093994 | 2.504690306 | 0 |
| *KNTC2* | 3.530722783 | 2.38615733 | 0 |
| *IL18R1* | 3.356426398 | 1.697254591 | 0 |
| *MMP1* | 3.255648367 | 2.232896954 | 0 |
| *PNMT* | 3.170833366 | 1.86252891 | 0 |
| *CD68* | 3.061562732 | 1.550760824 | 0 |
| *STARD3* | 2.968899114 | 1.65561188 | 0 |
| *TCAP* | 2.835284435 | 1.762935421 | 0 |
| *TRPV6* | 2.738080185 | 1.881934312 | 0 |
| *GRB7* | 2.539654021 | 1.71727436 | 0 |
| *IL23A* | 2.495769635 | 1.55800099 | 0 |
| *ERBB2* | 2.416874023 | 1.677542676 | 0 |
| *CX3CL1* | 2.293007372 | 1.505161104 | 0 |
| *IL34* | 1.903449385 | 1.345597645 | 0 |
| *CDC20* | 1.840049985 | 1.324024018 | 0 |
| *BRCA2* | 1.799143712 | 1.266761733 | 0 |
| *CNTNAP2* | 1.76264343 | 1.327495029 | 0 |
| *BUB1* | 1.725631312 | 1.276921853 | 0 |
| *MPHOSPH6* | 1.671561265 | 1.259333852 | 0 |
| *GSDMB* | 1.666433681 | 1.352498884 | 0 |
| *MFSD2A* | 1.664820363 | 1.325826852 | 0 |
| *TTK* | 1.64432023 | 1.284729799 | 0 |
| *MKI67* | 1.587795216 | 1.286457255 | 0 |
| *RRM2* | 1.543924701 | 1.293850587 | 0.29466332 |
| *PSMD3* | 1.509571727 | 1.266354257 | 0.29466332 |
| *CDCA5* | 1.505550252 | 1.257291427 | 0.29466332 |
| *CDH3* | 1.497696391 | 1.329935409 | 0.29466332 |
| *MYBL2* | 1.468958795 | 1.309235198 | 0.29466332 |
| *ORC6L* | 1.275744614 | 1.205273248 | 0.832990539 |
| *FGFR4* | 1.264713849 | 1.316082662 | 0.832990539 |
| *MID1* | 1.227472106 | 1.210977689 | 0.832990539 |
| *MYC* | 1.198731608 | 1.209187098 | 0.832990539 |
| *ORMDL3* | 1.18776889 | 1.217346839 | 0.832990539 |
| *EXO1* | 1.088026851 | 1.178613588 | 1.560919208 |
| *ABCC11* | 1.072314522 | 1.340406722 | 1.560919208 |
| *KIF2C* | 1.070907854 | 1.195672572 | 1.560919208 |
| *CDCA8* | 1.052081819 | 1.163964647 | 1.560919208 |
| *CEP55* | 1.013698547 | 1.158698705 | 1.560919208 |
| *CDKN3* | 0.998264335 | 1.156296707 | 1.560919208 |
| *GPNMB* | 0.986603261 | 1.160769479 | 1.560919208 |
| *MELK* | 0.971231023 | 1.163855248 | 1.560919208 |
| *CDC6* | 0.929253398 | 1.154823189 | 2.625182304 |
| *CENPA* | 0.887697288 | 1.14838903 | 2.625182304 |
| *ANLN* | 0.881322305 | 1.153374427 | 2.625182304 |
| *MND1* | 0.865524503 | 1.121091278 | 2.625182304 |
| *CDCA1* | 0.822817055 | 1.126298672 | 2.625182304 |
| *E2F1* | 0.774125313 | 1.122294473 | 4.060828877 |
| *CCNE1* | 0.741298266 | 1.111388466 | 4.060828877 |
| *ASPM* | 0.729760301 | 1.118237288 | 4.060828877 |
| *CXCL8* | 0.710119905 | 1.145408703 | 4.060828877 |
| *CCNB2* | 0.690409289 | 1.103818301 | 4.060828877 |
| *CENPF* | 0.666540413 | 1.105979495 | 4.060828877 |
| *BIRC5* | 0.643181022 | 1.118337816 | 4.060828877 |

| | | | |
|---|---|---|---|
| *NFIB* | 0.637264079 | 1.104532517 | 4.060828877 |
| *MIA* | 0.605069228 | 1.137459075 | 4.060828877 |
| *PTTG1* | 0.565056819 | 1.081899971 | 4.060828877 |
| *MMP11* | 0.50743366 | 1.113632231 | 4.060828877 |
| *XBP1* | 0.402387419 | 1.070652748 | 4.060828877 |
| *UBE2T* | 0.351007507 | 1.051121471 | 4.060828877 |
| *EGFR* | 0.243148437 | 1.044075529 | 4.060828877 |
| *GABRP* | 0.200741705 | 1.057833139 | 4.060828877 |
| *TMEM45B* | 0.194779114 | 1.034184644 | 4.060828877 |
| *FA2H* | 0.186358716 | 1.033727168 | 4.060828877 |
| *SF3A1* | 0.178464983 | 1.017726366 | 4.060828877 |
| *PHGDH* | 0.161465729 | 1.028900286 | 4.060828877 |
| *MSLN* | 0.158744082 | 1.036933732 | 4.060828877 |
| *MRAS* | 0.143758089 | 1.020793148 | 4.060828877 |
| *CCNB1* | 0.142444169 | 1.020004059 | 4.060828877 |
| *RAD51* | 0.115779144 | 1.016525433 | 4.060828877 |
| *BOC* | 0.086120236 | 1.015741711 | 4.060828877 |
| *NEK2* | 0.074146911 | 1.011282987 | 4.060828877 |
| *TOP2A* | 0.063748036 | 1.011041028 | 4.060828877 |
| *KLK5* | 0.037995357 | 1.010045698 | 4.060828877 |
| *SFRP1* | 0.01796086 | 1.004253758 | 4.060828877 |
| *AGR3* | -4.70416674 | 0.227844358 | 0 |
| *ESR1* | -4.370183029 | 0.300144947 | 0 |
| *SLC39A6* | -4.272757192 | 0.425753414 | 0 |
| *THSD4* | -3.969132231 | 0.489351644 | 0 |
| *FGFR2* | -3.495712951 | 0.527539315 | 0 |
| *MAPT* | -3.421174523 | 0.469288435 | 0 |
| *GATA3* | -3.164812732 | 0.486520584 | 0 |
| *PGR* | -3.119679895 | 0.452449463 | 0 |
| *CCND1* | -3.088840166 | 0.580631339 | 0 |
| *ERBB4* | -2.977742536 | 0.476460669 | 0 |
| *BAG1* | -2.71951687 | 0.684447722 | 0 |
| *AFF3* | -2.603930436 | 0.550512658 | 0 |
| *ZNF552* | -2.525054165 | 0.648488441 | 0 |
| *DNAJC12* | -2.401843239 | 0.593090297 | 0 |
| *ACTR3B* | -2.385862262 | 0.746126737 | 0 |
| *F12* | -2.349839867 | 0.664988425 | 0 |
| *MAGED2* | -2.33335199 | 0.698194432 | 0 |
| *MLPH* | -2.284355099 | 0.657426945 | 0 |
| *NAT1* | -2.184361356 | 0.615876422 | 0 |
| *ID4* | -2.047178293 | 0.671668691 | 0.29466332 |
| *TSPAN13* | -2.031383666 | 0.72100562 | 0.29466332 |
| *CXXC5* | -1.931939287 | 0.755015736 | 0.29466332 |
| *FOXA1* | -1.679455658 | 0.649784317 | 1.560919208 |
| *AGR2* | -1.645837036 | 0.640734726 | 1.560919208 |
| *CLUAP1* | -1.63126136 | 0.801596999 | 1.560919208 |
| *SIAH2* | -1.559499425 | 0.790947509 | 2.625182304 |
| *TROP2* | -1.554735917 | 0.766678081 | 2.625182304 |
| *TFCP2L1* | -1.3992131 | 0.771943951 | 4.060828877 |
| *BCL2* | -1.387873216 | 0.794535699 | 4.060828877 |
| *GPR160* | -1.365767644 | 0.790997783 | 4.060828877 |
| *FGFR1* | -1.306329385 | 0.799020129 | 4.060828877 |
| *RRAGA* | -1.292867121 | 0.871341785 | 4.060828877 |

| | | | |
|---|---|---|---|
| *MRPL19* | -1.251729108 | 0.85169401 | 4.253202338 |
| *RB1* | -1.142242261 | 0.86889319 | 4.956500918 |
| *CRYAB* | -1.125730717 | 0.793087195 | 4.956500918 |
| *KRT14* | -1.055922097 | 0.728665779 | 4.956500918 |
| *CREB3L4* | -1.052194544 | 0.845034774 | 4.956500918 |
| *KRT18* | -1.036470811 | 0.802520613 | 4.956500918 |
| *NECTIN4* | -0.922292836 | 0.865666407 | 6.600458365 |
| *SERPINB5* | -0.910833515 | 0.814746505 | 6.600458365 |
| *ERBB3* | -0.903070836 | 0.82743038 | 6.600458365 |
| *TYMS* | -0.885756746 | 0.875790757 | 6.600458365 |
| *KIF23* | -0.858281833 | 0.887207846 | 6.600458365 |
| *NDRG2* | -0.809083345 | 0.877768088 | 6.600458365 |
| *ACTG2* | -0.794322521 | 0.844339013 | 6.600458365 |
| *NQO1* | -0.730741556 | 0.8689043 | 7.422610529 |
| *NTN3* | -0.656786536 | 0.90507164 | 8.279421114 |
| *DGKD* | -0.632989146 | 0.917419868 | 8.279421114 |
| *AR* | -0.613786949 | 0.881690213 | 9.098234562 |
| *KCTD9* | -0.584549518 | 0.932533006 | 9.098234562 |
| *SPDEF* | -0.508913962 | 0.894718085 | 10.4114373 |
| *DNALI1* | -0.478713044 | 0.923114264 | 10.97326203 |
| *GARS* | -0.468496372 | 0.946525652 | 10.97326203 |
| *MDM2* | -0.465365376 | 0.937939872 | 10.97326203 |
| *BRCA1* | -0.298628818 | 0.953883978 | 10.97326203 |
| *AURKA* | -0.276546672 | 0.965647184 | 10.97326203 |
| *ETFA* | -0.275152086 | 0.963393829 | 10.97326203 |
| *KRT6B* | -0.238626563 | 0.939174787 | 10.97326203 |
| *KRT5* | -0.181988763 | 0.945261144 | 10.97326203 |
| *KRT17* | -0.136693473 | 0.95993688 | 10.97326203 |
| *UBE2C* | -0.12090754 | 0.983788103 | 10.97326203 |
| *BLVRA* | -0.091207743 | 0.988615186 | 10.97326203 |
| *FHOD1* | -0.079144818 | 0.989271262 | 10.97326203 |
| *FOXC1* | -0.074575962 | 0.987202383 | 10.97326203 |

**Supplementary Table S3. Differentially expressed genes between tumors with mature TLS (TLS 2a/2b) and tumors with no mature TLS (TLS 0/1a/1**

| Gene ID | Score(d) | Fold Change | FDR (q-value) |
|---|---|---|---|
| *CD79A* | 8.77596667 | 5.16712928 | 0 |
| *POU2AF1* | 7.22037499 | 3.39017368 | 0 |
| *IGL* | 6.93924308 | 4.58631559 | 0 |
| *IGKC* | 6.78084668 | 4.78433528 | 0 |
| *IGLV3.25* | 6.59945615 | 3.96525389 | 0 |
| *PIM2* | 6.33496462 | 2.3166367 | 0 |
| *CD27* | 6.32656836 | 2.30965712 | 0 |
| *IGJ* | 6.23824919 | 3.72613422 | 0 |
| *TNFRSF17* | 6.11678363 | 2.7794767 | 0 |
| *LY9* | 6.09441423 | 2.43385339 | 0 |
| *CD19* | 6.06940343 | 2.53306602 | 0 |
| *IRF4* | 5.98360848 | 2.75036537 | 0 |
| *SLAMF1* | 5.89685447 | 2.34155778 | 0 |
| *ITK* | 5.87590601 | 2.35338371 | 0 |
| *IL2RG* | 5.78320529 | 2.28098703 | 0 |
| *CTLA4* | 5.66762488 | 2.27889951 | 0 |
| *LAX1* | 5.66188096 | 2.29144664 | 0 |
| *CD8A* | 5.65665097 | 2.27361597 | 0 |
| *CD7* | 5.50725893 | 2.17047663 | 0 |
| *CXCL9* | 5.4483446 | 2.90362805 | 0 |
| *GZMA* | 5.11818025 | 2.11622455 | 0 |
| *SH2D1A* | 5.01355933 | 1.9088202 | 0 |
| *CD2* | 4.83768044 | 1.96806126 | 0 |
| *CXCL13* | 4.78674653 | 2.25510458 | 0 |
| *IRF8* | 4.75203916 | 1.75952834 | 0 |
| *KLRB1* | 4.74463077 | 1.95347834 | 0 |
| *CD3G* | 4.73906808 | 1.89181759 | 0 |
| *PDCD1* | 4.61902925 | 1.89759101 | 0 |
| *GZMB* | 4.50828409 | 2.22379488 | 0 |
| *STAT4* | 4.31470208 | 1.70593472 | 0 |
| *EOMES* | 4.18372454 | 1.9611449 | 0 |
| *CXCR6* | 4.12149409 | 1.88545994 | 0 |
| *MUCL1* | 3.86489807 | 3.09914546 | 0 |
| *GNLY* | 3.55820821 | 1.7959121 | 0 |
| *CD274* | 3.53729298 | 1.53776797 | 0 |
| *ISG20* | 3.48875913 | 1.58295539 | 0 |
| *CD4* | 3.41309746 | 1.45592487 | 0 |
| *CD40* | 3.40697712 | 1.46513837 | 0 |
| *IL18R1* | 3.35168075 | 1.54075224 | 0 |
| *SFRP1* | 3.23111352 | 1.85530093 | 0 |
| *BOC* | 3.20767682 | 1.60240439 | 0 |
| *CD3D* | 2.99456376 | 1.77821231 | 0 |
| *MYC* | 2.98266295 | 1.46793239 | 0 |
| *KRT17* | 2.80742714 | 1.97778231 | 0 |
| *HLA.C* | 2.76590996 | 1.46198693 | 0 |
| *CX3CL1* | 2.60252945 | 1.46016461 | 0 |

| | | | |
|---|---|---|---|
| *KRT5* | 2.58829001 | 1.91800618 | 0 |
| *STAT1* | 2.50053562 | 1.33729671 | 0 |
| *IL34* | 2.46196924 | 1.36812847 | 0 |
| *MID1* | 2.45983636 | 1.36750006 | 0 |
| *IRF1* | 2.35157588 | 1.35485856 | 0 |
| *NFIB* | 2.29794565 | 1.33841719 | 0 |
| *EAF2* | 2.29285316 | 1.33543589 | 0 |
| *EGFR* | 2.26155458 | 1.38572288 | 0 |
| *KYNU* | 2.11245374 | 1.35045632 | 0 |
| *GPNMB* | 2.04683858 | 1.28628324 | 0 |
| *MIA* | 2.02581713 | 1.41984573 | 0 |
| *CRYAB* | 2.01728627 | 1.40189208 | 0 |
| *KRT14* | 1.97672628 | 1.62033954 | 0 |
| *CD86* | 1.93730238 | 1.23444874 | 0 |
| *CD84* | 1.86195549 | 1.24384041 | 0 |
| *CDH3* | 1.85049052 | 1.3340273 | 0 |
| *LGALS9* | 1.78998633 | 1.23075562 | 0 |
| *KLRD1* | 1.77983928 | 1.29221142 | 0 |
| *ACTG2* | 1.75009562 | 1.35494221 | 0 |
| *TRPV6* | 1.73741833 | 1.38952319 | 0 |
| *S100A9* | 1.72924824 | 1.44361133 | 0 |
| *CXCL8* | 1.67776661 | 1.29844204 | 0 |
| *GABRP* | 1.66154131 | 1.46025772 | 0 |
| *KLK5* | 1.56920669 | 1.3995775 | 0.25235835 |
| *FOXC1* | 1.54625349 | 1.24494704 | 0.25235835 |
| *ID4* | 1.44844855 | 1.25928314 | 0.41592395 |
| *KRT6B* | 1.42679733 | 1.35782806 | 0.41592395 |
| *SERPINB5* | 1.38593786 | 1.28976798 | 0.41592395 |
| *C2orf54* | 1.38438952 | 1.34077587 | 0.41592395 |
| *CNTNAP2* | 1.3807733 | 1.20061332 | 0.41592395 |
| *MRAS* | 1.3088917 | 1.16550087 | 0.57589469 |
| *MPHOSPH6* | 1.10388914 | 1.133078 | 1.24777184 |
| *STARD3* | 1.06783668 | 1.16017098 | 1.24777184 |
| *CD68* | 0.97947198 | 1.12247664 | 1.90045249 |
| *IL23A* | 0.94413022 | 1.1481664 | 1.90045249 |
| *KNTC2* | 0.85394182 | 1.18879628 | 2.95524909 |
| *PNMT* | 0.84163073 | 1.14542705 | 2.95524909 |
| *MSLN* | 0.79206414 | 1.15885192 | 2.95524909 |
| *NDRG2* | 0.75201669 | 1.10430359 | 4.55480349 |
| *MMP1* | 0.727672 | 1.15869777 | 4.55480349 |
| *KCTD9* | 0.70669067 | 1.07149696 | 4.55480349 |
| *GSDMB* | 0.68376613 | 1.10638562 | 5.79846912 |
| *SF3A1* | 0.67183286 | 1.0557056 | 5.79846912 |
| *TCAP* | 0.36932514 | 1.06237222 | 12.5911522 |
| *ERBB2* | 0.3616169 | 1.06532374 | 12.5911522 |
| *RB1* | 0.24231667 | 1.02475275 | 15.8387685 |
| *PHGDH* | 0.21350711 | 1.03122364 | 15.8387685 |
| *XBP1* | 0.19458229 | 1.02728293 | 15.8387685 |
| *CCNE1* | 0.02096741 | 1.00244962 | 15.8387685 |

| | | | |
|---|---|---|---|
| *F12* | -3.325402 | 0.62422596 | 0 |
| *SLC39A6* | -3.0017479 | 0.61160716 | 0 |
| *AGR3* | -2.9574234 | 0.46536648 | 0 |
| *TSPAN13* | -2.8569901 | 0.68684077 | 0 |
| *GATA3* | -2.7927294 | 0.595263 | 0 |
| *ESR1* | -2.7815437 | 0.5333891 | 0 |
| *AGR2* | -2.716664 | 0.55020291 | 0 |
| *THSD4* | -2.6691211 | 0.67402091 | 0 |
| *MLPH* | -2.6027495 | 0.67724735 | 0 |
| *KIF23* | -2.538546 | 0.75007761 | 0 |
| *ZNF552* | -2.4981189 | 0.70506043 | 0 |
| *ERBB4* | -2.4742092 | 0.60488902 | 0 |
| *MAPT* | -2.4511093 | 0.64156126 | 0 |
| *UBE2C* | -2.4257378 | 0.76627317 | 0 |
| *NAT1* | -2.3205926 | 0.65737086 | 0 |
| *TOP2A* | -2.3123947 | 0.72344507 | 0 |
| *BIRC5* | -2.3016996 | 0.72238054 | 0 |
| *CCND1* | -2.2504236 | 0.72170678 | 0 |
| *TYMS* | -2.036236 | 0.78031523 | 0 |
| *AURKA* | -2.035959 | 0.81097849 | 0 |
| *NEK2* | -1.9887893 | 0.78245142 | 0.41592395 |
| *CENPA* | -1.9215229 | 0.78350174 | 0.41592395 |
| *CCNB1* | -1.9013772 | 0.80639945 | 0.41592395 |
| *RAD51* | -1.8434027 | 0.80841719 | 0.41592395 |
| *CDKN3* | -1.8384679 | 0.80424827 | 0.41592395 |
| *SPDEF* | -1.8068675 | 0.72516582 | 0.41592395 |
| *FOXA1* | -1.8038917 | 0.68580214 | 0.41592395 |
| *GPR160* | -1.7927627 | 0.7782 | 0.41592395 |
| *E2F1* | -1.7818592 | 0.80536483 | 0.41592395 |
| *FGFR2* | -1.7808227 | 0.76508627 | 0.41592395 |
| *ANLN* | -1.7803172 | 0.79100857 | 0.41592395 |
| *UBE2T* | -1.776568 | 0.81403105 | 0.41592395 |
| *DNAJC12* | -1.6860402 | 0.74092338 | 0.41592395 |
| *SIAH2* | -1.685301 | 0.81283262 | 0.41592395 |
| *MAGED2* | -1.6621763 | 0.81096517 | 0.57589469 |
| *ASPM* | -1.6494145 | 0.81396932 | 0.57589469 |
| *RRM2* | -1.6465221 | 0.79929498 | 0.57589469 |
| *CREB3L4* | -1.6102314 | 0.8103503 | 0.57589469 |
| *BRCA1* | -1.5607935 | 0.81768694 | 0.57589469 |
| *CDCA8* | -1.560589 | 0.8319929 | 0.57589469 |
| *BAG1* | -1.5354258 | 0.83869013 | 0.57589469 |
| *CDC6* | -1.4927925 | 0.82823617 | 0.57589469 |
| *CDCA5* | -1.4505938 | 0.83531746 | 1.24777184 |
| *ERBB3* | -1.4378372 | 0.78197925 | 1.24777184 |
| *ORC6L* | -1.3921007 | 0.84668532 | 1.24777184 |
| *AFF3* | -1.3908631 | 0.77023954 | 1.24777184 |
| *MELK* | -1.3794268 | 0.83887216 | 1.24777184 |
| *CEP55* | -1.378844 | 0.84903528 | 1.24777184 |
| *CXXC5* | -1.3071545 | 0.85612168 | 1.90045249 |

| | | | |
|---|---|---|---|
| *EXO1* | -1.3047355 | 0.85150521 | 1.90045249 |
| *MRPL19* | -1.1780302 | 0.88396826 | 2.95524909 |
| *PTTG1* | -1.1178097 | 0.88076998 | 2.95524909 |
| *CDCA1* | -1.0998285 | 0.87815056 | 2.95524909 |
| *KRT18* | -1.082395 | 0.82927916 | 4.55480349 |
| *TTK* | -1.063679 | 0.87594101 | 4.55480349 |
| *CCNB2* | -1.0322092 | 0.88616183 | 4.55480349 |
| *FA2H* | -0.9965608 | 0.86541097 | 4.55480349 |
| *DGKD* | -0.9727982 | 0.89739064 | 4.55480349 |
| *PGR* | -0.9509549 | 0.82046383 | 4.55480349 |
| *FGFR4* | -0.9384325 | 0.84636883 | 4.55480349 |
| *MYBL2* | -0.9383806 | 0.86910594 | 4.55480349 |
| *ACTR3B* | -0.9057827 | 0.91264656 | 5.79846912 |
| *CDC20* | -0.8492226 | 0.8995247 | 5.79846912 |
| *NECTIN4* | -0.8350409 | 0.89871604 | 5.79846912 |
| *MKI67* | -0.8037498 | 0.90102231 | 5.79846912 |
| *BUB1* | -0.7577497 | 0.91586874 | 6.23080904 |
| *TMEM45B* | -0.7191192 | 0.90371869 | 6.23080904 |
| *CENPF* | -0.6976576 | 0.91753277 | 7.06285945 |
| *NQO1* | -0.6695306 | 0.90035271 | 7.06285945 |
| *BRCA2* | -0.653652 | 0.93187849 | 7.06285945 |
| *BLVRA* | -0.550854 | 0.94511834 | 12.5911522 |
| *TROP2* | -0.5313824 | 0.92807743 | 12.5911522 |
| *GARS* | -0.5144933 | 0.95186154 | 12.5911522 |
| *AR* | -0.4669655 | 0.92481534 | 12.5911522 |
| *MND1* | -0.4523817 | 0.95223985 | 12.5911522 |
| *RRAGA* | -0.4456025 | 0.96186166 | 12.5911522 |
| *DNALI1* | -0.4252358 | 0.94362231 | 12.5911522 |
| *KIF2C* | -0.3830631 | 0.94916993 | 13.3563269 |
| *PSMD3* | -0.3614822 | 0.95487259 | 13.3563269 |
| *FHOD1* | -0.3455083 | 0.96222005 | 15.8387685 |
| *MMP11* | -0.3157812 | 0.94668937 | 15.8387685 |
| *CLUAP1* | -0.2755089 | 0.96986376 | 16.2352941 |
| *FGFR1* | -0.2742819 | 0.96192612 | 16.2352941 |
| *TFCP2L1* | -0.228793 | 0.96595185 | 16.7201426 |
| *GRB7* | -0.2129516 | 0.96354007 | 16.7201426 |
| *ORMDL3* | -0.2128272 | 0.97165326 | 16.7201426 |
| *NTN3* | -0.2086342 | 0.97430255 | 16.7201426 |
| *MDM2* | -0.2007445 | 0.97767122 | 16.7201426 |
| *MFSD2A* | -0.1493685 | 0.97946852 | 17.4278952 |
| *ABCC11* | -0.1476671 | 0.96755299 | 17.4278952 |
| *BCL2* | -0.1317961 | 0.9822949 | 17.4278952 |
| *ETFA* | -0.0230762 | 0.99744274 | 17.4278952 |

**Supplementary Table S4. Factors associated with the presence of TLS (TLS≠0 vs 0) and mature TLS (TLS 2a/2b)**

| **Factors associated with the presence of TLS (TLS≠0 vs 0)** | | | | | |
|---|---|---|---|---|---|
| | **N (%)** | **Univariable OR (95% CI)** | **p-value** | **Adjusted OR (95% CI)** | **p-value** |
| **IHC Subtype** | | | | | |
| HR+/HER2- | 656 (60%) | 1 [Reference] | | 1 [Reference] | |
| HER2+ | 289 (26%) | 1.57 (1.17-2.11) | **0.003** | 0.88 (0.61-1.25) | 0.470 |
| TNBC | 150 (14%) | 1.19 (0.80-1.76) | 0.367 | 0.71 (0.45-1.12) | 0.710 |
| **Tumor Site** | | | | | |
| Primary | 630 (57%) | 1 [Reference] | | 1 [Reference] | |
| Metastasis | 472 (43%) | 0.41 (0.31-0.54) | **<0.001** | 0.56 (0.40-0.78) | **<0.001** |
| **IGG signature (continuous)** | - | 1.94 (1.71-2.16) | **<0.001** | 1.57 (1.39-1.79) | **<0.001** |
| **TILs % (continuous)** | - | 1.06 (1.05-1.07) | **<0.001** | 1.04 (1.02-1.05) | **<0.001** |
| **Interaction:** IGG × TILs interaction, $p = 2.3 \times 10^{-6}$ | | | | | |
| **Factors associated with the presence of mature TLS (TLS 2a/2b)** | | | | | |
| | **N (%)** | **Univariable OR (95% CI)** | **p-value** | **Adjusted OR (95% CI)** | **p-value** |
| **IHC Subtype** | | | | | |
| HR+/HER2- | 656 (60%) | 1 [Reference] | | 1 [Reference] | |
| HER2+ | 289 (26%) | 1.44 (0.92-2.22) | 0.104 | 0.73 (0.43-1.21) | 0.225 |
| TNBC | 150 (14%) | 1.93 (1.14-3.18) | **0.012** | 1.29 (0.71-2.27) | 0.394 |
| **Tumor Site** | | | | | |
| Primary | 630 (57%) | 1 [Reference] | | 1 [Reference] | |
| Metastasis | 472 (43%) | 0.39 (0.25-0.60) | **<0.001** | 0.57 (0.34-0.92) | **0.024** |
| **IGG signature (continuous)** | - | 2.11 (1.82-2.48) | **<0.001** | 1.63 (1.36-1.96) | **<0.001** |
| **TILs % (continuous)** | - | 1.05 (1.04-1.07) | **<0.001** | 1.03 (1.02-1.05) | **<0.001** |
| **Interaction:** IGG × TILs interaction, p = 0.003 | | | | | |

**Supplementary Table S5. Multivariable logistic regression models evaluating factors associated with mature TLS (TLS 2a/2b).**

| Model | Variable | OR | 95% CI | p-value | AIC |
|---|---|---|---|---|---|
| TLS abundance + TILs | TLS abundance | 1.92 | 1.14-3.25 | 0.014 | 437.56 |
| | TILs | 1.03 | 1.02-1.04 | <0.001 | |
| TLS abundance + IGG | TLS abundance | 2.07 | 1.25-3.41 | 0.004 | 448.84 |
| | IGG signature | 1.45 | 1.19-1.76 | <0.001 | |
| TLS abundance + IGG + TILs | TLS abundance | 1.86 | 1.10-3.16 | 0.020 | 434.87 |
| | IGG signature | 1.27 | 1.02-1.59 | 0.033 | |
| | TILs | 1.02 | 1.01-1.05 | 0.009 | |

**Supplementary Table S6. Gene composition and overlap of the TLS-GEP and TILs-GEP signatures.**

| TLS-GEP genes | TILs-GEP genes |
|---|---|
| ***CD79A*** | *ANLN* |
| *ITK* | ***BOC*** |
| ***IL2RG*** | *CCNB2* |
| *MUCL1* | *CCNE1* |
| ***BOC*** | *CD19* |
| ***MYC*** | *CD3D* |
| *IRF1* | *CD7* |
| *EAF2* | ***CD79A*** |
| *CD84* | *CDC6* |
| *KLRD1* | *CTLA4* |
| *CXCL8* | *CXCL13* |
| | *EOMES* |
| | *GPNMB* |
| | *GRB7* |
| | *GSDMB* |
| | *IGL* |
| | *IGLV3.25* |
| | *IL18R1* |
| | ***IL2RG*** |
| | *MMP1* |
| | *MRAS* |
| | ***MYC*** |
| | *PTTG1* |
| | *RAD51* |
| | *S100A9* |
| | *SF3A1* |
| | *STAT1* |
| | *TMEM45B* |
| | *TOP2A* |
| | *TRPV6* |

**(*) Overlap genes in bold**

**Supplementary Table S7. Multivariable Cox regression analyses of immune organization signatures adjusted for PAM50 ROR-P across the TCGA, METABRIC, and SCAN-B HR+/HER2- breast cancer cohorts.**

| Cohort | Model | Variable | HR (95% CI) | P value |
|---|---|---|---|---|
| TCGA | ROR-P + TLS_GEP | ROR-P | 1.28 (1.03–1.61) | 0.028 |
| TCGA | ROR-P + TLS_GEP | TLS GEP | 0.77 (0.62–0.95) | 0.014 |
| TCGA | ROR-P + IGG | ROR-P | 1.26 (1.01–1.56) | 0.038 |
| TCGA | ROR-P + IGG | IGG | 0.80 (0.66–0.97) | 0.023 |
| TCGA | ROR-P + TIL_GEP | ROR-P | 1.34 (1.08-1.66) | 0.009 |
| TCGA | ROR-P + TIL_GEP | TIL GEP | 0.76 (0.62-0.93) | 0.008 |
| TCGA | ROR-P + TLS_GEP + TIL_GEP | ROR-P | 1.34 (1.05-1.70) | 0.019 |
| TCGA | ROR-P + TLS_GEP + TIL_GEP | TLS-GEP | 0.86 (0.57-1.30) | 0.474 |
| TCGA | ROR-P + TLS_GEP + TIL_GEP | TIL-GEP | 0.83 (0.53-1.30) | 0.409 |
| METABRIC | ROR-P + TLS_GEP | ROR-P | 1.32 (1.23–1.43) | <0.001 |
| METABRIC | ROR-P + TLS_GEP | TLS GEP | 0.88 (0.82–0.95) | 0.001 |
| METABRIC | ROR-P + IGG | ROR-P | 1.33 (1.23–1.43) | <0.001 |
| METABRIC | ROR-P + IGG | IGG | 0.89 (0.82–0.96) | 0.002 |
| METABRIC | ROR-P + TIL_GEP | ROR-P | 1.35 (1.26–1.46) | <0.001 |
| METABRIC | ROR-P + TIL_GEP | TIL GEP | 0.94 (0.87–1.01) | 0.08 |
| METABRIC | ROR-P + TLS_GEP + TIL_GEP | ROR-P | 1.32 (1.22-1.42) | <0.001 |
| METABRIC | ROR-P + TLS_GEP + TIL_GEP | TLS-GEP | 0.87 )0.79-0.96) | 0.006 |
| METABRIC | ROR-P + TLS_GEP + TIL_GEP | TIL-GEP | 1.02 (0.93--1.12) | 0.685 |
| SCAN-B | ROR-P + TLS_GEP | ROR-P | 1.26 (1.16–1.36) | <0.001 |
| SCAN-B | ROR-P + TLS_GEP | TLS GEP | 0.81 (0.75–0.87) | <0.001 |
| SCAN-B | ROR-P + IGG | ROR-P | 1.31 (1.21–1.41) | <0.001 |
| SCAN-B | ROR-P + IGG | IGG | 0.83 (0.78–0.89) | <0.001 |
| SCAN-B | ROR-P + TIL_GEP | ROR-P | 1.35 (1.25–1.46) | <0.001 |
| SCAN-B | ROR-P + TIL_GEP | TIL GEP | 0.86 (0.80–0.93) | <0.001 |
| SCAN-B | ROR-P + TLS_GEP + TIL_GEP | ROR-P | 1.25 (1.15-1.36) | <0.001 |
| SCAN-B | ROR-P + TLS_GEP + TIL_GEP | TLS-GEP | 0.80 (0.72-0.89) | <0.001 |
| SCAN-B | ROR-P + TLS_GEP + TIL_GEP | TIL-GEP | 1.02 (0.91-1.24) | 0.743 |

Hazard ratios are shown per standard deviation increase of each continuous variable. Multivariable Cox regression models were adjusted for PAM50 Risk of Recurrence (ROR-P).

**Supplementary Table S8.Main clinicopathological characteristics of patients with available TCR-seq data stratified by tertiary lymphoid structure (TLS) status.**

| **Clinicopathological variables** | | **All** (n: 34) | **TLS+** (n: 13) | **TLS-** (n: 21) |
|---|---|---|---|---|
| **Subtype** | HER2-positive | 14 (41.2%) | 9 (69.2%) | 5 (23.8%) |
| | Triple negative | 20 (58.8%) | 4 (30.8%) | 16 (76.2%) |
| **Stage at diagnosis** | I | 4 (11.8%) | 2 (15.4%) | 2 (9.5%) |
| | IIA | 15 (44.1%) | 7 (53.8%) | 8 (38.1%) |
| | IIB | 7 (20.6%) | 2 (15.4%) | 5 (23.8%) |
| | IIIA | 1 (2.9%) | 0 | 1 (4.8%) |
| | IIIB | 1 (2.9%) | 0 | 1 (4.8%) |
| | IIIC | 6 (17.6%) | 2 (15.4%) | 4 (19%) |
| **Grade** | 1 | 3 (9.1%) | 2 (15.4%) | 1 (5%) |
| | 2 | 10 (30.3%) | 5 (38.5%) | 5 (25%) |
| | 3 | 20 (60.6%) | 6 (46.2%) | 14 (70%) |
| **TILs (%)** [median, range] | | 5 [1-70] | 12 [1-65] | 5 [1-70] |
| **TLS** | 0 | 21 (61.8%) | NA | 21 (100%) |
| | 1a | 5 (14.7%) | 5 (38.5%) | NA |
| | 1b | 2 (5.9%) | 2 (15.4%) | NA |
| | 2a | 4 (11.8%) | 4 (30.8%) | NA |
| | | 2 (5.9%) | 2 (15.4%) | |
| **Neoadjuvant therapy** | Carboplatin and taxane | 3 (8.8%) | NA | 3 (14.3%) |
| | Carboplatin, taxane and anthracyclines | 14 (41.2%) | 3 (23.1%) | 11 (52.4%) |
| | Taxane and anthracyclines | 1 (2.9%) | NA | 1 (4.8%) |
| | Datopotamab-deruxtecan | 1 (2.9%) | 0 | 1 (4.8%) |
| | Olaparib | 1 (2.9%) | 1 (7.7%) | |
| | Carboplatin, taxane and anti-HER2 | 4 (11.8%) | 3 (23.1%) | 1 (4.8%) |
| | Taxane, anthracyclines and anti-HER2 | 4 (11.8%) | 2 (15.4%) | 2 (9.5%) |
| | Taxane and anti-HER2 | 6 (17.6%) | 4 (30.8%) | 2 (9.5%) |
| **Pathologic complete response** (pCR) | Yes | 22 (64.7%) | 11 (84.6%) | 11 (52.4%) |
| | No | 12 (35.3%) | 2 (15.4%) | 10 (47.6%) |

**Supplementary Table S9.Characteristics of the spatial transcriptomics cohort (n = 8 samples)**

| ID | TLS | TILs (%) | IGG | TLS ROIs | Non-TLS TME ROIs | Tumor ROIs |
|---|---|---|---|---|---|---|
| 1 | Immature TLS (1a) | 15 | High | 2 | 4 | 4 |
| 2 | No TLS | 12 | High | 0 | 4 | 4 |
| 3 | No TLS | 1 | Medium | 0 | 4 | 4 |
| 4 | No TLS | 8 | Low | 0 | 3 | 1 |
| 5 | Mature TLS (2a) | 70 | High | 3 | 4 | 4 |
| 6 | Immature TLS (1a) | 6 | Medium | 1 | 4 | 4 |
| 7 | No TLS | 6 | High | 0 | 4 | 4 |
| 8 | No TLS | 1 | Medium | 0 | 4 | 4 |

**Supplementary Table S10.Predefined gene modules and marker genes used to calculate mean expression per region of interest (ROI).**

| Gene | Gene module |
|---|---|
| GZMB | Cytotoxic T cells |
| PRF1 | Cytotoxic T cells |
| GZMA | Cytotoxic T cells |
| KLRK1 | Cytotoxic T cells |
| GZMK | Effector T cells |
| CXCR3 | Effector T cells |
| IFNG | Effector T cells |
| PDCD1 | Exhausted T cells |
| TIGIT | Exhausted T cells |
| LAG3 | Exhausted T cells |
| HAVCR2 | Exhausted T cells |
| CTLA4 | Exhausted T cells |
| TOX | Exhausted T cells |
| FOXP3 | T-regs |
| IL2RA | T-regs |
| IKZF2 | T-regs |
| CTLA4 | T-regs |
| TIGIT | T-regs |
| IL1B | M1 macrophages |
| TNF | M1 macrophages |
| CXCL9 | M1 macrophages |
| CXCL10 | M1 macrophages |
| STAT1 | M1 macrophages |
| IRF1 | M1 macrophages |
| CD163 | M2 macrophages |
| MRC1 | M2 macrophages |
| MSR1 | M2 macrophages |
| CCL18 | M2 macrophages |
| VSIG4 | M2 macrophages |
| STAB1 | M2 macrophages |
| SIGLEC1 | M2 macrophages |
| LAMP3 | Dendritic cells |
| CCR7 | Dendritic cells |
| CD1C | Dendritic cells |
| FCER1A | Dendritic cells |
| CLEC10A | Dendritic cells |
| COL11A1 | Matrix CAF |
| POSTN | Matrix CAF |
| COL10A1 | Matrix CAF |
| COL5A1 | Matrix CAF |
| COL5A2 | Matrix CAF |
| COMP | Matrix CAF |
| FAP | Matrix CAF |
| LRRC15 | Matrix CAF |
| THBS2 | Matrix CAF |
| CXCL12 | Inflammatory CAF |

| | |
|---|---|
| IL6 | Inflammatory CAF |
| CCL19 | Inflammatory CAF |
| VCAM1 | Inflammatory CAF |
| MKI67 | Proliferative tumor |
| TOP2A | Proliferative tumor |
| CCNB1 | Proliferative tumor |
| CCNB2 | Proliferative tumor |
| CDK1 | Proliferative tumor |
| AURKA | Proliferative tumor |
| PLK1 | Proliferative tumor |
| UBE2C | Proliferative tumor |
| CA9 | Hypoxic tumor |
| BNIP3 | Hypoxic tumor |
| NDRG1 | Hypoxic tumor |
| EGLN3 | Hypoxic tumor |
| ERBB2 | HER2-like tumor |
| GSDMB | HER2-like tumor |
| MUC1 | HER2-like tumor |
| VTCN1 | HER2-like tumor |
| PROM2 | HER2-like tumor |
| ANKRD30A | HER2-like tumor |
| STAT1 | IFN response |
| STAT2 | IFN response |
| IRF1 | IFN response |
| IRF7 | IFN response |
| CXCL9 | IFN response |
| CXCL10 | IFN response |
| CXCL11 | IFN response |
| CXCL13 | TLS |
| CCL19 | TLS |
| MS4A1 | TLS |
| CD79A | TLS |
| LAMP3 | TLS |
| MZB1 | TLS |
| CD27 | IGG signature |
| CD79A | IGG signature |
| IL2RG | IGG signature |
| PIM2 | IGG signature |
| POU2AF1 | IGG signature |
| TNFRSF17 | IGG signature |
| MZB1 | Plasma cell |
| PRDM1 | Plasma cell |
| IRF4 | Plasma cell |
| TNFRSF17 | Plasma cell |
| BCL6 | Germinal center |
| AICDA | Germinal center |
| LMO2 | Germinal center |
| CD19 | Germinal center |
| MS4A1 | Germinal center |

| | |
|---|---|
| CXCR5 | Germinal center |
| CXCL13 | Germinal center |
| CR2 | Germinal center |
| PAX5 | Germinal center |

**Supplementary Table S11.Treatment-induced changes in TLS-GEP and TIL-GEP across studies.**

| Study | Therapy | Marker | n_pairs | mean_delta | ci_low | ci_high | p_value | p_TLS_vs_TIL |
|---|---|---|---|---|---|---|---|---|
| PROMETEO-I | T-VEC + atezolizumab | TLS-GEP | 18 | 1.461526316 | 0.892125009 | 2.030927623 | 0.000355446 | 0.011539555 |
| PROMETEO-I | T-VEC + atezolizumab | TIL-GEP | 18 | 0.969089163 | 0.35431333 | 1.583864996 | 0.00897457 | 0.011539555 |
| PROMETEO-II | CDK4/6i + ET | TLS-GEP | 20 | 0.903944865 | 0.495370253 | 1.312519477 | 0.001088441 | 0.001825294 |
| PROMETEO-II | CDK4/6i + ET | TIL-GEP | 20 | 0.281523098 | -0.04386741 | 0.606913604 | 0.070197451 | 0.001825294 |
| YOUNGSTER | RT boost | TLS-GEP | 13 | 0.463319571 | -0.18652072 | 1.113159866 | 0.059172068 | 0.107973209 |
| YOUNGSTER | RT boost | TIL-GEP | 13 | 0.226302565 | -0.093997 | 0.546602135 | 0.184235113 | 0.107973209 |
| BIONHER | Trastuzumab + pertuzumab | TLS-GEP | 63 | 0.11396909 | -0.05268059 | 0.280618768 | 0.216561995 | 0.920925081 |
| BIONHER | Trastuzumab + pertuzumab | TIL-GEP | 63 | 0.103634374 | -0.06596712 | 0.273235863 | 0.28091712 | 0.920925081 |
| ELIPSE | Elacestrant | TLS-GEP | 22 | 0.417675052 | 0.128545163 | 0.706804941 | 0.009397295 | 0.435876278 |
| ELIPSE | Elacestrant | TIL-GEP | 22 | 0.33254953 | 0.017278302 | 0.647820759 | 0.008545581 | 0.435876278 |
| CORALEEN | CDK4/6i + ET | TLS-GEP | 29 | 0.569521977 | 0.26208169 | 0.876962265 | 0.001479828 | 0.000129553 |
| CORALEEN | CDK4/6i + ET | TIL-GEP | 29 | -0.04841637 | -0.4320063 | 0.335173568 | 0.828806902 | 0.000129553 |
| CORALEEN | Chemotherapy | TLS-GEP | 35 | 0.587227394 | 0.355002136 | 0.819452652 | 5.7932E-05 | 3.42578E-06 |
| CORALEEN | Chemotherapy | TIL-GEP | 35 | -0.24931423 | -0.51104838 | 0.012419923 | 0.041429742 | 3.42578E-06 |
| PREOPERATIVE ET | Endocrine therapy | TLS-GEP | 93 | 0.89581072 | 0.725754974 | 1.065866466 | 1.89E-13 | 0.00029363 |
| PREOPERATIVE ET | Endocrine therapy | TIL-GEP | 93 | 0.609734022 | 0.435248322 | 0.784219723 | 1.2152E-08 | 0.00029363 |

**Supplementary Table S11.Association between treatment-induced changes in TLS-GEP and TIL-GEP and treatment response.**

| Model | OR (95% CI) | P value |
|---|---|---|
| ΔTLS-GEP adjusted for study | 1.46 (1.00–2.14) | 0.051 |
| ΔTIL-GEP adjusted for study | 0.92 (0.65–1.31) | 0.655 |
| ΔTLS-GEP adjusted for ΔTIL-GEP and study | 2.28 (1.35–3.98) | 0.003 |
| ΔTIL-GEP adjusted for ΔTLS-GEP and study | 0.54 (0.32–0.89) | 0.018 |
