## Supplementary Methods for "Immune organization defines adaptive immune competence and clinical outcome in breast cancer"

**Supplementary Methods 1. Guidelines for identification and assessment of TLS**

1. A tertiary lymphoid structure (TLS) is an organized, non-encapsulated, ectopic lymphoid aggregate located within the tumor microenvironment.
2. TLS should be reported only when located within the stromal compartment of the invasive tumor.
3. TLS assessment should be restricted to the invasive tumor border, defined as the stromal area within 1 mm of the invasive tumor front.
4. TLS located outside this region, or those associated with ductal carcinoma in situ (DCIS) or normal mammary epithelium, should be excluded.
5. Areas affected by crush artifact, necrosis, regressive hyalinization, or prior biopsy-related changes should be excluded from evaluation.
6. TLS classification is based on two features:
    a. TLS number: ≤ 2 (low) or > 2 (high).
    b. Germinal center (GC) presence: TLS are considered mature when a GC is present and immature when absent.

    Germinal centers are identified morphologically as well-defined regions with organized architecture and relatively reduced lymphoid density, reflecting the presence of larger activated B-cells, fewer small lymphocytes, and occasional macrophages. Features consistent with follicular dendritic cell networks and early vascular structures, such as high endothelial venules (HEVs), may be observed. When morphology is inconclusive, immunohistochemical markers (e.g., Ki-67, CD20, CD23) may assist in confirming GC presence.

7. The combination of TLS number and GC presence defines four categories. In cases of discordance, GC presence takes precedence over TLS number due to its greater biological relevance.

| | **≤ 2 TLS** | **> 2 TLS** |
|---|---|---|
| **NO GC** | 1a | 1b |
| **With GC** | 2a | 2b |

8. TLS identification should be performed on a single 4-µm hematoxylin and eosin (H&E)–stained section. TLS are identified at low magnification (4–10×), and GC presence should be confirmed at higher magnification (20–40×).

9. Full tumor sections are preferred for accurate TLS quantification. Core biopsies may be used but are inherently limited due to restricted tissue sampling and potential underrepresentation of tumor margins, where TLS are more frequently located.

**Abbreviations:** TLS, tertiary lymphoid structure; GC, germinal center; H&E, hematoxylin and eosin; DCIS, ductal carcinoma in situ; HEV, high endothelial venule; IHC, immunohistochemistry.
